# Cross-cohort analysis of expression and splicing quantitative trait loci in TOPMed

**DOI:** 10.1101/2025.02.19.25322561

**Authors:** Peter Orchard, Thomas W. Blackwell, Linda Kachuri, Peter J. Castaldi, Michael H. Cho, Stephanie A. Christenson, Peter Durda, Stacey Gabriel, Craig P. Hersh, Scott Huntsman, Seungyong Hwang, Roby Joehanes, Mari Johnson, Xingnan Li, Honghuang Lin, Ching-Ti Liu, Yongmei Liu, Angel C.Y. Mak, Ani W. Manichaikul, David Paik, Aabida Saferali, Joshua D. Smith, Kent D. Taylor, Russell P. Tracy, Jiongming Wang, Mingqiang Wang, Joshua S. Weinstock, Jeffrey Weiss, Heather E. Wheeler, Ying Zhou, Sebastian Zoellner, Joseph C. Wu, Luisa Mestroni, Sharon Graw, Matthew R.G. Taylor, Victor E. Ortega, Craig W. Johnson, Weiniu Gan, Goncalo Abecasis, Deborah A. Nickerson, Namrata Gupta, Kristin Ardlie, Prescott G. Woodruff, Yinan Zheng, Russell P. Bowler, Deborah A. Meyers, Alex Reiner, Charles Kooperberg, Elad Ziv, Vasan S. Ramachandran, Martin G. Larson, L. Adrienne Cupples, Esteban G. Burchard, Edwin K Silverman, Stephen S. Rich, Nancy Heard-Costa, Hua Tang, Jerome I. Rotter, Albert V. Smith, Daniel Levy, NHLBI TOPMed Consortium Multi-Omics Working Group, NHLBI TOPMed Consortium, François Aguet, Laura Scott, Laura M. Raffield, Stephen C.J. Parker

## Abstract

Most genetic variants associated with complex traits and diseases occur in non-coding genomic regions and are hypothesized to regulate gene expression. To understand the genetics underlying gene expression variability, we characterize 14,324 ancestrally diverse RNA-sequencing samples from the NHLBI Trans-Omics for Precision Medicine (TOPMed) program and integrate whole genome sequencing data to perform *cis* and *trans* expression and splicing quantitative trait locus (*cis*-/trans-e/sQTL) analyses in six tissues and cell types, most notably whole blood (N=6,454) and lung (N=1,291). We show this dataset enables greater detection of secondary cis-e/sQTL signals than was achieved in previous studies, and that secondary cis-eQTL and primary trans-eQTL signal discovery is not saturated even though eGene discovery is. Most TOPMed trans-eQTL signals colocalize with cis-e/sQTL signals, suggesting many trans signals are mediated by cis signals. We fine-map European UK BioBank GWAS signals from 164 traits and colocalize the resulting 34,107 fine-mapped GWAS signals with TOPMed e/sQTL signals, finding that of 10,611 GWAS signals with a colocalization, 7,096 GWAS signals colocalize with at least one secondary e/sQTL signal. These results demonstrate that larger e/sQTL analyses will continue to uncover secondary e/sQTL signals, and that these new signals will benefit GWAS interpretation.

## Introduction

Most genetic variants associated with complex human traits occur in non-coding genomic regions (Hindorff et al., 2009). This complicates the task of interpreting variant - trait associations identified in genome wide association studies (GWAS). While GWAS signals are commonly annotated with the name of the closest gene, without additional data it is often impossible to determine the gene(s) impacted by trait-associated variants as well as the functional effects of a variant on a gene product. Molecular quantitative trait locus (QTL) analyses detect associations between genetic variants and molecular traits such as gene expression (eQTLs) or RNA splicing (sQTLs). Because non-coding GWAS variants are expected to act through the transcriptome, e/sQTL analyses are a critical step in interpreting the molecular cascade of events at GWAS signals.

e/sQTL studies have made important contributions to understanding gene regulation and GWAS signals (Brown et al., 2023; GTEx Consortium, 2020; Liu et al., 2022; Võsa et al., 2021; Yao et al., 2017). Recent notable efforts include the GTEx Consortium (GTEx Consortium, 2020), which performed e/sQTL analyses in 49 tissues and cell types and is the most expansive published resource in terms of tissue and cell type breadth (many genetic variants affect the transcriptome in a cell type-specific manner); the eQTLGen Consortium (Võsa et al., 2021) which published a study including 31,684 whole blood and PBMC samples, representing the largest eQTL study in terms of sample size published to date; and the DIRECT consortium (Brown et al., 2023), which performed QTL analyses of gene expression, proteins, and metabolites in 3,029 blood and plasma samples.

These studies have multiple limitations. The largest sample size in any single GTEx tissue is 706, which limits power to detect relatively weaker genetic effects. 84.6% of GTEx donors and 100% of donors in the DIRECT study are of European ancestry, which limits power to detect e/sQTLs driven by genetic variants absent or at low frequency in European samples. eQTLGen has technical limitations: the study was a meta-analysis of 37 separate studies, many of which profiled gene expression and genotypes using gene expression and genotyping arrays. This heterogeneity in gene expression profiling and genotyping platforms complicates cross-cohort analysis; as a result eQTLGen did not attempt to identify more than one eQTL signal per gene (i.e., no secondary eQTL signals), despite the fact that many genes are expected to have more than one eQTL signal and many of these secondary eQTLs may colocalize with GWAS signals (GTEx Consortium, 2020; Zeng et al., 2019). Furthermore eQTLGen did not attempt to detect sQTLs, as splicing is difficult to quantify with gene expression arrays.

The National Heart, Lung, and Blood Institute’s (NHLBI) Trans-Omics for Precision Medicine (TOPMed) program has performed whole genome sequencing (WGS) on >180k samples from >85 phenotypically and ethnically diverse cohorts, generating a large dataset of genetic variation (Taliun et al., 2021) (https://topmed.nhlbi.nih.gov/). Here we characterize 14,324 TOPMed RNA-sequencing samples and use existing WGS sample genotypes to perform cis- and trans-expression and splicing quantitative trait locus (cis- and trans-eQTL / sQTL) analyses in six tissues and cell types (Fig 1A,B), with the following aims: (1) generate a large, high-quality e/sQTL reference dataset that addresses the above-described shortcomings of previous studies, and (2) explore secondary e/sQTL signal discovery with larger sample sizes and characterize whether this benefits e/sQTL - GWAS signal colocalizations. We find that the large TOPMed sample sizes available for e/sQTL analyses in whole blood (N=6,454) and lung (N=1,291) combined with our mega-analysis approach (directly combining data from all studies, rather than meta-analyzing studies) enables greater detection of secondary cis-e/sQTL signals than was achieved in previous studies. This TOPMed e/sQTL resource complements GTEx, which has e/sQTL for a broader array of tissues but fewer samples per tissue and (in whole blood) less ancestral diversity, as well as eQTLGen, which has a larger sample size but no secondary signal discovery or splicing analyses. We show that even with 6,454 whole blood samples, discovery of secondary cis-eQTL and primary trans-eQTL signals has not reached saturation even though eGene discovery has. We find that most trans-eQTL signals at TOPMed sample sizes colocalize with cis-e/sQTL signals, consistent with the hypothesis that many trans signals are mediated by cis signals. We fine-map 34,107 UK BioBank GWAS signals from 164 traits (Bycroft et al., 2018; Sudlow et al., 2015) (https://pan.ukbb.broadinstitute.org) and compare GWAS signal colocalization between TOPMed and GTEx e/sQTL signals (Fig 1C). Of 2,308 GWAS signals with a TOPMed cis-e/sQTL colocalization and no GTEx cis-e/sQTL colocalization, 1,200 colocalize only with secondary TOPMed cis-e/sQTL signals, demonstrating that larger e/sQTL sample sizes enabling extensive secondary e/sQTL signal discovery substantially improve our ability to identify e/sQTL - GWAS signal colocalizations.

**Figure 1.**
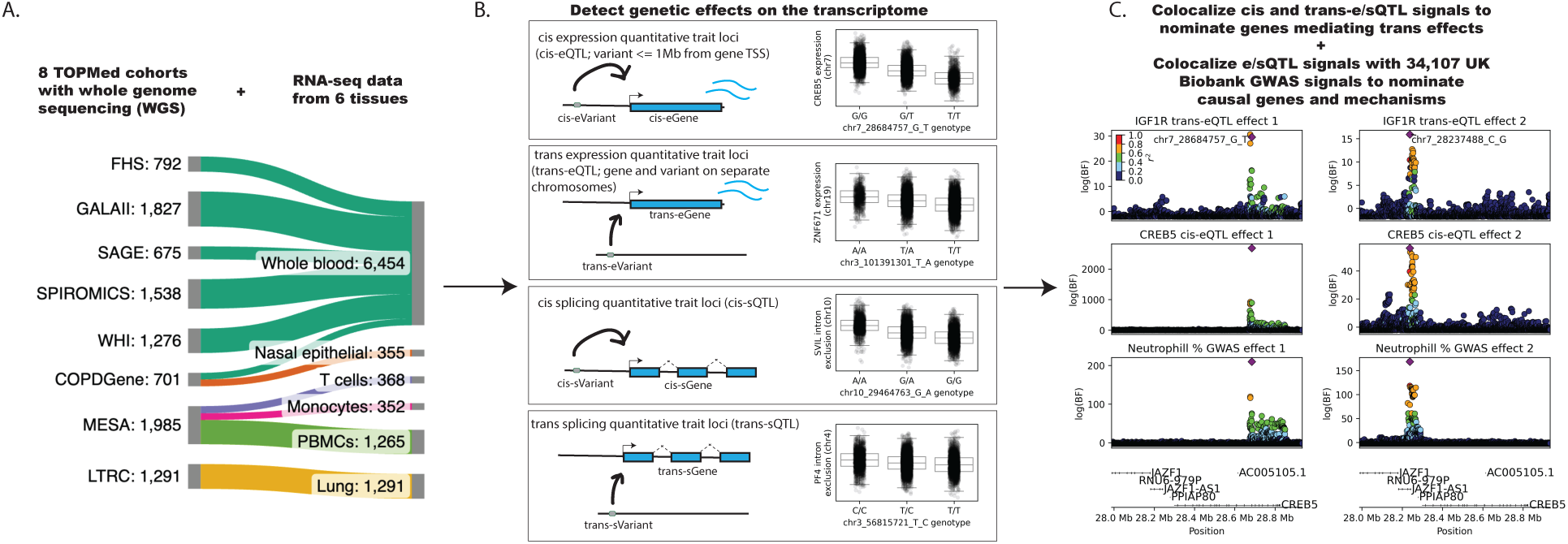
Study design. (A) RNA-seq sample sizes for cis- and trans-e/sQTL scans, by TOPMed study and tissue. The full TOPMed study names and accompanying abbreviations are: Framingham Heart Study (FHS); Gene-Environments and Admixture in Latino Asthmatics (GALA II); Study of African Americans, Asthma, Genes, & Environments (SAGE); Subpopulations and Intermediate Outcome Measures In COPD Study (SPIROMICS); Women’s Health Initiative (WHI); COPDGene Study (COPDGene); Multi-Ethnic Study of Atherosclerosis (MESA); Lung Tissue Research Consortium (LTRC). Diagram generated with SankeyMATIC. (B) cis-e/sQTL scans tested MAF *→* 0.01 variants within 1Mb of gene TSS (for whole blood, a scan with variant MAF *→* 0.001 was also performed). Trans scans tested variant - gene pairs on separate chromosomes (variants with MAF *→* 0.05). Splicing phenotypes (intron excision ratios) were derived using LeafCutter (Li et al., 2018). (C) cis-e/sQTL signals were colocalized with trans-e/sQTL signals to nominate genes mediating trans e!ects, and cis- and trans-e/sQTL signals were colocalized with 34,107 GWAS signals from 164 UK BioBank GWAS to nominate genes and molecular mechanisms underlying GWAS signals.

## Results

### Sample demographics

We performed RNA-seq on 14,324 samples from the TOPMed program, including 12,863 samples from 10,195 donors with WGS-derived genotypes. RNA-seq samples represented six tissue types and eight individual TOPMed cohorts (Fig S1; Supplementary methods). Samples from each tissue were derived from single TOPMed cohorts, with the exception of whole blood which included samples from six cohorts. Sample characteristics such as ancestry, sex, and age varied between tissues (Fig S1, S2).

For each sample we computed the genetically inferred global ancestry, the fraction of the genome assigned to each of seven ancestry groups (Sub-saharan Africa (AFR); Native America (AMR); East Asia (EAS); Europe (EUR); Middle East (MES); and Central and South Asia (SAS)) using a reference panel derived from the Human Genome Diversity Panel (J. Z. Li et al., 2008) (Fig S2). Lung and nasal epithelial samples were heavily European (87.0% of lung samples and 91.1% of nasal epithelial samples were >=90% EUR; 11.4% of lung samples and 8.1% of nasal epithelial samples were admixed, defined here as less than 90% of the genome attributable to a single ancestry). 50.8% of whole blood samples were >= 90% EUR, while 46.7% were admixed (mostly representing EUR, AFR, and AMR ancestry). The three tissues and cell types derived solely from the MESA cohort (PBMCs, monocytes, and T cells) were the most diverse. Most notably, of PBMC samples, 31.7% were >= 90% EUR, 6.5% were >= 90% AFR, 5.2% were > 90% EAS, and 56.5% were admixed.

### Primary cis-e/sQTL signal discovery

To identify genetic variants associated with gene expression or RNA splicing, we performed cis-e/sQTL scans using unrelated subjects and a single time point for each individual (Fig 1A,B), testing variants with minor allele frequency (MAF) >= 0.01. The sample size per tissue ranged from 352 (monocytes) to 6,454 (whole blood) (Fig S3, S4). We identified 9,330 - 19,465 genes with a significant cis-eQTL (cis-eGenes) and 3,290 - 8,795 genes with a significant cis-sQTL (cis-sGenes) (5% FDR; Fig 2A). The number of significant cis-e/sGenes in each tissue correlated strongly with sample size. In whole blood and lung, the tissues profiled in both TOPMed and GTEx, the rate of cis-e/sGene discovery in TOPMed exceeded that of GTEx, reflecting the greater TOPMed sample sizes (Fig S5; e.g., 19,465 / 22,187 tested genes (87.8%) in TOPMed whole blood were eGenes vs. 12,360 / 20,315 (60.8%) in GTEx whole blood). The rate of cis-eGene discovery in TOPMed whole blood was comparable to that in the larger eQTLGen study (16,987 / 19,250 tested genes (88.2%) were eQTLGen eGenes), suggesting that cis-eGene discovery is largely saturated at the TOPMed whole blood sample size (N=6,454). Primary cis-e/sQTL signals in TOPMed showed high direction-of-effect concordance in GTEx and eQTLGen (Fig S6-S11), supporting the robustness of our results.

**Figure 2.**
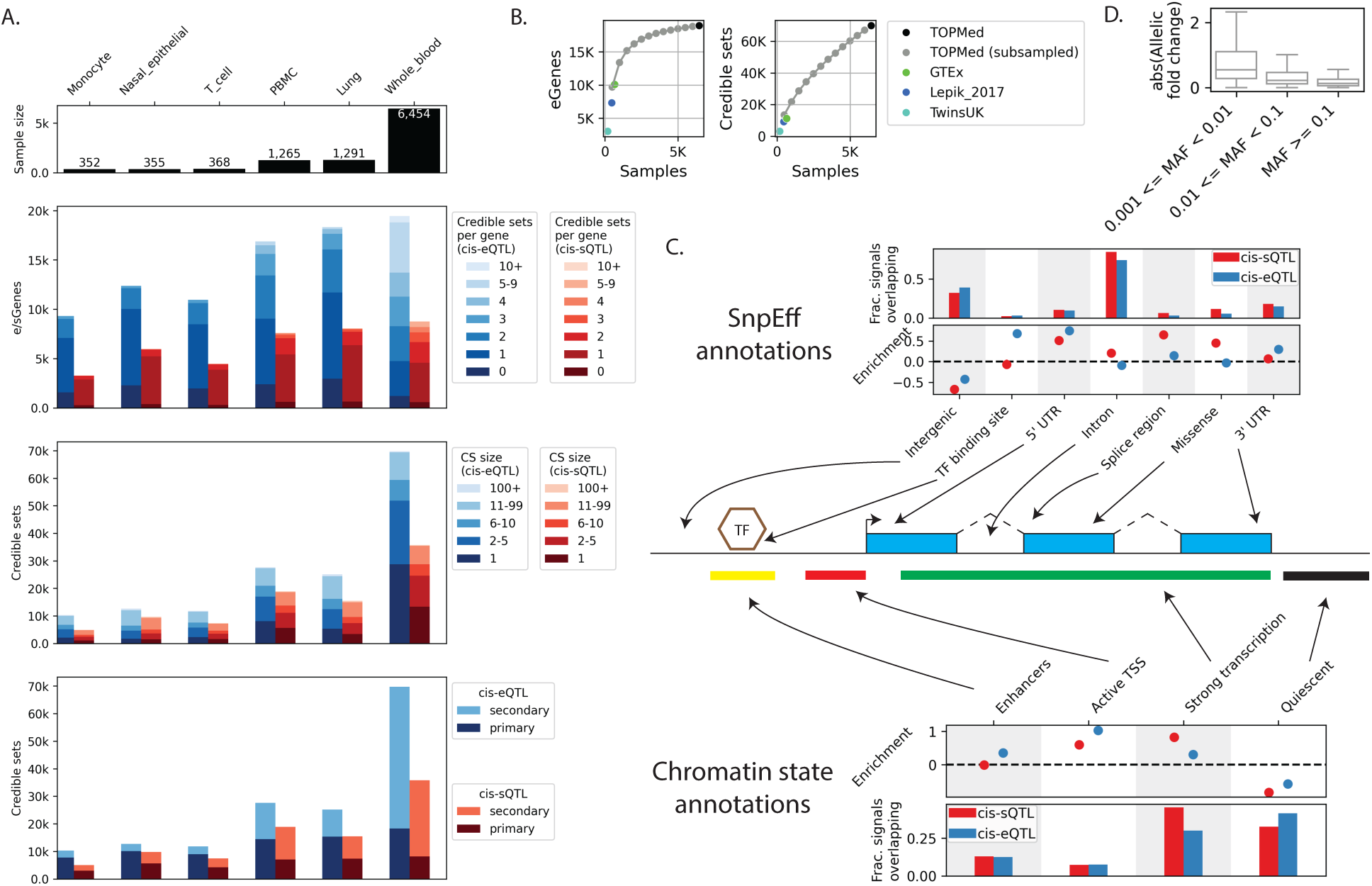
cis-e/sQTL summary. (A) Sample sizes per tissue (top panel), number of genes with a significant cis-e/sQTL (cis-e/sGenes) and number of SuSiE credible sets discovered per cis-e/sGene (second from top), credible set sizes (third from top), and number of primary or secondary (including tertiary, quaternary, etc.) cis-e/sQTL signals per tissue (bottom). (B) Cis-eQTL saturation analysis, and comparison to other published datasets (from eQTL-Catalogue or GTEx). Number of cis-eGenes discovered at each downsampled sample size (left) and total number of cis-eQTL signals discovered (right; 95% credible sets). Results are shown for 1% FDR cis-Genes, as eQTL-Catalogue fine-maps QTL signals for 1% FDR cis-e/sGenes. (C) Functional annotation enrichments for whole blood cis-e/sQTLs. Enrichment calculated relative to control credible sets matched on MAF, LD, and number of genes tested against; error bars represent 95% confidence intervals. (D) E!ect size (absolute allelic fold change) for cis-eQTL in whole blood scan with MAF 0.1% threshold.

### Fine-mapping of cis-e/sQTL signals reveals tens of thousands of secondary signals

Many genes are expected to have multiple independent cis-e/sQTLs, and detecting secondary signals provides a more comprehensive picture of gene expression regulation and may increase the number of colocalizations with other QTL and GWAS signals. We used SuSiE (G. Wang et al., 2020) to fine-map independent cis-e/sQTL signals, generating 95% credible sets for each signal. Throughout this work ‘signal’ denotes a 95% credible set, ‘secondary signal’ denotes any e/sQTL signal that is not the most significant signal for the gene (see Methods), and ‘top PIP variant’ denotes the variant with the greatest posterior inclusion probability (PIP) in the 95% credible set (i.e., the variant statistically judged most likely to be causal). This identified 10,282 - 69,766 and 4,992 - 35,770 total cis-eQTL and cis-sQTL signals per tissue, respectively (Fig 2A). In whole blood and lung in particular, this represents a meaningful increase (518.2% increase vs GTEx whole blood cis-eQTLs; 16.3% increase vs DIRECT whole blood cis-eQTLs; 114.4% increase vs GTEx lung cis-eQTLs) in signals detected relative to previous studies (Fig 2B) (Brown et al., 2023; Buil et al., 2015; GTEx Consortium, 2020; Kerimov et al., 2021; Lepik et al., 2017) (this could not be evaluated in eQTLGen, which reported only primary cis-eQTLs). In whole blood, we discovered a mean of 3.6 cis-eQTL and 1.9 cis-sQTL signals per cis-e/sGene (Fig. 2A). 28715 (41.2%) whole blood cis-eQTL and 13393 (37.4%) whole blood cis-sQTL 95% credible sets contained a single variant (Fig. 2A), representing a substantial increase in resolution relative to previously published results (e.g. 2,112 (18.7%) of GTEx whole blood cis-eQTL credible sets contained a single variant; Fig S12). Ancestry differences between TOPMed and previously published datasets likely contribute, as TOPMed whole blood donors (69% EUR, as defined by the ancestry underlying the greatest fraction of the each donor’s genome) are more diverse than GTEx (∼84.6% EUR), TwinsUK (∼100% EUR), or Lepik_2017 (derived from the Estonian BioBank (Leitsalu et al., 2015)). These results emphasize the benefits of large sample sizes and diverse ancestries in characterizing genetic effects on the transcriptome.

As whole blood cis-eGene discovery seems to be largely saturated at the TOPMed sample size, we asked whether the total number of cis-eQTL signals might also be at or near saturation. To test this we downsampled the whole blood sample set and re-performed cis-eGene discovery and cis-eQTL fine-mapping (Fig. 2B, S13). While cis-eGene discovery began to saturate around 3,000 samples, the number of total cis-eQTL signals did not saturate even at the full whole blood sample size, suggesting that additional cis-eQTL signals will be discovered with larger sample sizes. cis-sGene discovery likewise shows signs of saturation, while the number of total cis-sQTL signals discovered is not saturated (Fig. S14).

### Characterization of cis-eQTL and sQTL by functional annotation, tissue, and ancestry

Cis-eQTLs were enriched in TF binding sites, and cis-e/sQTL signals were enriched in splice regions, though cis-eQTLs not colocalizing with cis-sQTLs showed no splice region enrichment (Fig 2C, S15-S17; Table S1, S2). cis-eQTL signals were enriched in enhancer, active transcription start site (TSS), flanking active TSS and weak/strong transcription chromatin states, while cis-sQTL signals were enriched in active TSS, flanking active TSS, weak/strong transcription, transcription at gene 5’ and 3’, and genic enhancer chromatin states (Fig 2C, S18-19). This is consistent with the fact that cis-eQTL signals are more likely than cis-sQTL signals to be in promoters, and less likely to be in a gene body (Fig S20). Both primary and secondary cis-sQTL signals were enriched in splice regions, but the enrichment was stronger for primary cis-sQTLs (Fig S21, Table S3). Primary and secondary cis-eQTL signals were enriched in active TSS and enhancer chromatin states, but active TSS enrichment was stronger for primary cis-eQTL signals (Fig S22).

In the case that a TF tends to activate rather than repress gene expression, eVariant alleles that disrupt that TF’s motifs should associate with decreases in gene expression, while alleles that create a binding site should associate with increases in gene expression. The reverse is expected for TFs that primarily repress gene expression. To explore these effects in our data, we performed variant-sensitive motif scanning and, for each TF position weight matrix, noted which cis-eVariant alleles favor TF binding and which alleles associate with higher gene expression (Fig S23; Table S4). While many TFs are known to act as both activators and repressors in different contexts (Mavrothalassitis & Ghysdael, 2000), several TF motifs display associations with gene expression that suggest a tendency to activate. Most notably, alleles creating or strengthening many ETS family motifs associated with increased gene expression in ∼70% of instances, suggesting the corresponding TFs tend to activate gene expression. The tendency for ETS motif-strengthening alleles to associate with increased gene expression and the lack of motif-strengthening alleles associated with decreased gene expression mirrors previous findings in pancreatic islets (Viñuela et al., 2020).

Cis-e/sQTL signals may be specific to a tissue, or shared across tissues, and sharing may reflect the extent to which local genomic function is shared across tissues (Aguet et al., 2017; GTEx Consortium, 2020). We determined the fraction of signals shared -- defined here as credible set overlap -- between lung/nasal epithelial and whole blood, conditional on a gene being tested in both tissues (Fig S24). The fraction of lung and nasal epithelial cis-eQTL signals shared with blood was 46.9% and 44.7%, respectively. Sharing was higher for cis-sQTL signals (63.5% and 60.2%, respectively; Fisher’s Exact Test p-value for lung=9.98×10^-192^ and odds ratio = 1.97). Primary signals were more likely to be shared than secondary signals (Fig S24; for lung cis-eQTL, Fisher’s Exact Test p = 2.9×10^-69^ and odds ratio = 1.66). Relative to unshared signals, shared eQTL signals showed stronger enrichment in active TSS chromatin states (Fig S25-28; Table S5) and shared sQTL signals were enriched in strong transcription chromatin states.

To gauge the contribution of each ancestry to cis-eQTL discovery, we examined differences in MAFs across ancestry groups for each cis-eQTL signal in whole blood and PBMCs (the tissue with the largest sample size and the tissue with the greatest fraction of non-EUR samples, respectively) (Fig S29-32). A sample was assigned to an ancestry if it had global genetic ancestry >= 75% from that ancestry, or >= 50% in the case of AMR ancestry (see Methods). This revealed that inclusion of participants of African (AFR) ancestry had an outsized influence on eQTL signal discovery, consistent with previous eQTL analyses within TOPMed (Kachuri et al., 2023) and expectations based on the greater genetic heterogeneity and smaller linkage disequilibrium (LD) blocks in AFR populations relative to non-AFR populations (D. E. Reich et al., 2001; Shifman et al., 2003; The 1000 Genomes Project Consortium, 2015). For example, 5.9% of 25,894 unique PBMC cis-eVariants had MAF >= 0.01 in AFR samples and MAF < 0.001 in all other ancestries, compared to 0.3% for EUR samples. (Fig S32). As many studies use only EUR samples, we additionally examined MAF differences using only EUR and AFR samples. 15.3% of 63,784 unique whole blood cis-eVariants had MAF >= 0.01 in AFR samples and MAF < 0.001 in EUR, compared to 0.2% of variants with MAF >= 0.01 in EUR samples and MAF < 0.001 in AFR. For PBMCs, these values were 9.7% and 0.7%, respectively.

### Large whole blood sample size enables discovery of lower MAF cis-e/sQTL signals

Due to sample size limitations, previously published cis-e/sQTL studies frequently test only variants with MAF of 0.01 or greater (GTEx Consortium, 2020; Kerimov et al., 2021; Scott et al., 2016; Varshney et al., 2017; Viñuela et al., 2020; Võsa et al., 2021). To explore our ability to detect cis-e/sQTL signals involving rarer variants, we ran a second set of whole blood cis-e/sQTL scans applying a 0.001 MAF threshold (corresponding to a minor allele count (MAC) threshold of 13, comparable to the MAC threshold (14) in GTEx whole blood (N=670) when applying a MAF threshold of 0.01). With the lower MAF threshold, the average number of variants tested against each gene more than doubled (7,866 vs 17,195 for the cis-eQTL scan). Notably, the fraction of tested variants that were specific to one ancestry increased as the MAF threshold decreased (0.2% of variants in MAF > 0.01 scan vs 16.7% of variants in MAF > 0.001 scan, when considering the three well-represented ancestries (EUR, AFR, AMR)); the fraction of tested variants that were specific to AFR samples accounted for most of these ancestry-specific variants (Fig S33-35). While the number of cis-e/sGenes showed little change compared to a 0.01 MAF threshold (19,465 and 19,394 cis-eGenes at 0.01 and 0.001 MAF thresholds, respectively, and 8,795 vs 8,873 cis-sGenes), the number of total signals detected increased by ∼9% (from from 69,766 to 76,545 cis-eQTLs, and 35,770 to 39,001 cis-sQTLs). As expected, low MAF cis-eQTLs had larger effect sizes than high MAF cis-eQTLs (Fig 2D) (Liu et al., 2022). The top PIP variant for 16,217 cis-eQTL and 7,302 cis-sQTL signals had MAF < 0.01. However, a majority of the signals with variant MAF < 0.01 had MAF >= 0.01 in at least one of the well-represented ancestries (Fig S36) and could therefore be said to represent common variants in at least one ancestry. 7,479 cis-eQTL and 3,290 cis-sQTL signals had MAF < 0.01 in all three well-represented ancestries. cis-e/sQTL signals with MAF < 0.01 in all three ancestries generally showed directionally similar enrichment patterns as those with MAF >= 0.01 in all three ancestries, though the degree of enrichment often differed (Fig S37-38; Table S6): for example, cis-eQTL signals with MAF < 0.01 were more strongly enriched in the active TSS chromatin state than those signals with MAF >= 0.01 (Fig S38), and cis-sQTL signals with MAF < 0.01 were more strongly enriched in splice regions and genic enhancers than those with MAF >= 0.01 (Fig S37).

### Integration of trans-e/sQTL signals with cis signals identify regulatory relationships and biological pathways

To identify genetic variants that may impact gene expression and splicing in trans, we performed trans-e/sQTL scans at a MAF >= 0.05 using gene - variant pairs on separate chromosomes. Detection of trans-e/sQTL signals is more difficult than detection of cis signals, as trans signals are weaker than cis signals and an untargeted trans scan entails a higher multiple testing burden. We identified 1 - 1,725 trans-eGenes and 0 - 127 trans-sGenes per tissue (5% FDR; Fig 3A, S39). A saturation analysis in whole blood revealed a near-linear relationship between sample size and the number of trans-eGenes discovered, suggesting that trans-eGene discovery is not saturated at the current sample sizes (Fig S40). Trans-eQTLs had smaller effect sizes than cis-eQTLs (Fig 3B).

**Figure 3.**
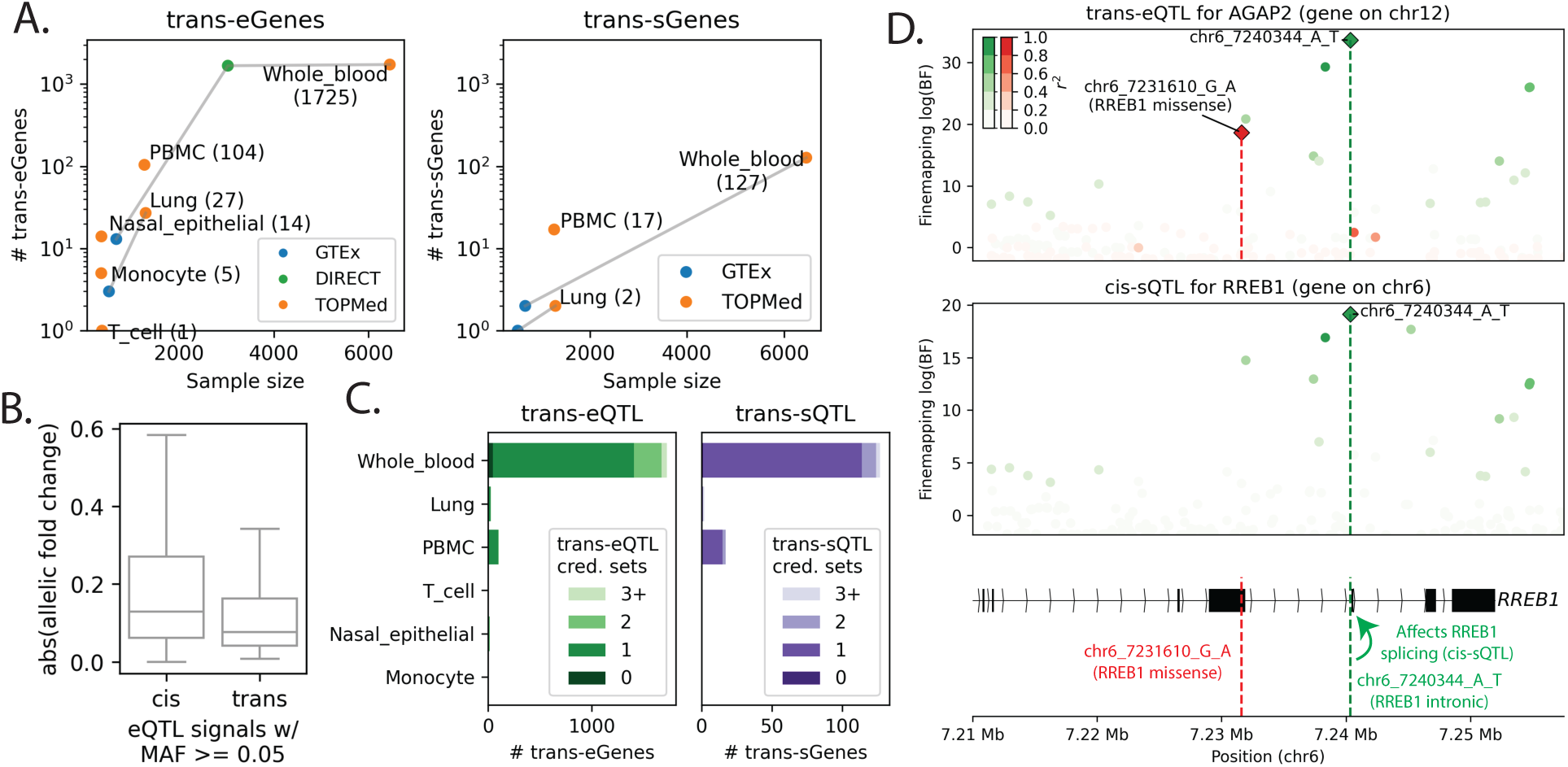
trans-e/sQTL results. (A) number of trans-eGenes and trans-sGenes as a function of sample size. TOPMed tissues are linked to the corresponding GTEx and DIRECT tissues. Unlike TOPMed and GTEx, DIRECT did not apply a MAF threshold. TOPMed and GTEx defined trans as ‘di!erent chromosome’, while DIRECT defined trans as ’di!erent chromosome or gene - variant pair *→* 5Mb apart’. (B) E!ect size (absolute allelic fold change) for trans-eQTL and cis-eQTL with MAF *→* 0.05 in whole blood (cis-eQTL e!ect sizes from MAF *→* 0.001 scan). (C) Number of trans-eQTL and trans-sQTL credible sets discovered when fine-mapping the region around primary trans-e/sQTL signals (+/- 1Mb). (D), (E) Two (of three total) whole blood trans-eQTL signals for gene *AGAP2*. One colocalizes with a *RREB1* cis-sQTL, one contains a *RREB1* missense variant, suggesting that both variants impact *AGAP2* expression via distinct functional e!ects on *RREB1*. The credible set variants for the third *AGAP2* trans-eQTL signal are in an *RREB1* intron.

In whole blood, after linkage-disequilibrium (LD)-based clumping of primary trans-eVariants 614 unique trans-eVariants were discovered for 1,725 trans-eGenes; 171 variants were trans-eVariants for >1 gene (accounting for 1,282 total trans-eGenes; Table S7). Trans-eVariants were enriched in missense and 3’ UTR variants, though only a small fraction overlapped these annotations (Fig S41-42). Our top trans-eVariant (ranked by number of trans-eGenes) was rs946588154 (chr7_50342615_A_G), which was a trans-eVariant for 260 genes. This variant was a cis-eQTL credible set variant for genes *IKZF1* and *GRB10* and a cis-sQTL variant for *IKZF1* in whole blood. The second ranked trans-Variant was rs1354034 (chr3_56815721_T_C), which had 82 trans-eGenes and was not a cis-e/sQTL in any tissue.

In whole blood, we identified 61 unique trans-sVariants (Table S8). The four trans-sVariants with the largest number of trans-sGenes in whole blood included variants rs946588154 (chr7_50342615_A_G; 16 trans-sGenes) and rs1354034 (chr3_56815721_T_C; 30 trans-sGenes) discussed above; rs6939187 (chr6_163408503_T_C; 8 trans-sGenes), a whole blood cis-eQTL for lncRNA CAHM as well as gene *QKI*, which encodes an RNA splicing regulator (Wu et al., 2002); and rs7613875 (chr3_49934081_C_A; 5 trans-sGenes), which sits 5.9kb upstream of the gene encoding splicing regulator *RBM6* (Huber et al., 2012). Overall in whole blood, eleven variants were trans-sVariants for more than one trans-sGene (accounting for 77 total trans-sGenes).

Different trans-eGenes sharing a trans-eVariant might represent genes in a pathway or network. To explore this, we performed gene ontology (GO) and KEGG pathway enrichment analyses for groups of 10+ genes sharing the same whole blood trans-eVariant (Table S9) (Raudvere et al., 2019). 14 of 25 such groups showed significant GO:BP or KEGG pathway enrichments. For example, one *cis*-eVariant (rs16947425; chr17_64066984_C_A) for gene *ERN1*, which encodes endonuclease IRE1a, and gene *PRR29* (unknown function) was a *trans*-eVariant for thirteen *trans*-eGenes, including known *ERN1* downstream target *XBP1* (Shen et al., 2001; Yoshida et al., 2001) and *XBP1* target genes including *DNAJB9*. *ERN1* is a regulator of the endoplasmic reticulum (ER) stress response (Shen et al., 2001; Yoshida et al., 2001), and five of the thirteen *trans*-eGenes were in the “Protein processing in ER” KEGG pathway (39.1-fold enrichment; nominal p=1.0×10^-7^). Among ER response pathways IRE1a-XBP1 is the most highly conserved (Hetz & Papa, 2018; Richardson et al., 2010), with an emerging role in regulation of inflammation and immune response (Grootjans et al., 2016; Pramanik et al., 2018; Richardson et al., 2010). These results suggest that trans signals can provide insight into biological pathways.

### Primary and secondary trans signals converge on potential mediator genes

To identify trans-e/sGenes that may share multiple signals with a potential regulatory cis-e/sGene, we fine-mapped trans-e/sQTL signals within the 2Mb window centered on each trans-e/sGene’s lead trans-e/sVariant (Fig 3C; Table S10). Within the 2Mb windows, 327 of the 1,876 trans-eGene - tissue pairs had >1 trans-eQTL (16 of the 146 trans-sGene - tissue pairs).

In whole blood, trans-eVariants were enriched for overlap with cis-e/sQTL signals and trans-sVariants were enriched in cis-eQTL (Fig S43). 32.9% of unique whole blood trans-eVariants overlap at least one whole blood cis-eQTL or cis-sQTL signal (31.1% for trans-sVariants). Cis-eGenes for cis-eQTL overlapping trans-eQTL were 3.7-fold enriched for transcription factor (TF) genes (p < 1×10^-3^; Fig S44). The overlap between cis and trans signals is consistent with the hypothesis that many trans effects are mediated by cis effects.

To nominate mediating mechanisms and genes for whole blood trans-e/sQTL signals, we colocalized whole blood trans signals with cis signals, and noted whether each trans-e/sQTL credible set contained protein-altering variants or 3’/5’ UTR variants (Fig 3D; Fig S45-47). We also fine-mapped 34,107 EUR GWAS signals for 164 UK Biobank traits (Table S11) and determined whether each trans-e/sQTL credible set colocalized with a GWAS signal for blood cell abundance or other traits (Fig S45-47). Only 15.9% of trans-eQTL credible sets and 16.9% of trans-sQTL credible sets did not colocalize with a cis or GWAS signal or contain protein-altering / UTR variants. 58.8% and 45.8% of trans-eQTL and trans-sQTL credible sets, respectively, colocalized with at least one cis-e/sQTL signal. For some trans-eGenes with multiple credible sets, these annotations suggest that different trans-eQTL signals for the same gene are due to different functional effects on a single mediator gene. For example, the trans-eGene *AGAP2* showed three trans-eQTL signals, one of which colocalized with a *RREB1* cis-sQTL and one of which represented a *RREB1* missense variant (Fig 3D).

Across all tissues, we found 30 cis-e/sGenes with > 1 cis-e/sQTL signal colocalizing with > 1 trans-e/sQTL signal from at least one trans-e/sGene (158 unique cis-trans gene pairs) (Table S12). *IKZF1* cis signals accounted for 76.6% (121 / 158) of these multi-colocalizing gene pairs. Of 146 gene pairs where > 1 trans-eQTL from gene 1 colocalized with > 1 cis-eQTL from gene 2, directions of effect were consistent across the colocalizing signals (variants that increased expression of gene 1 either always increased or always decreased expression of gene 2) in 140 (95.9%) of the pairs (Table S12). In the most extreme case, *trans*-eGene *BTN3A3* showed 4 whole blood *trans*-eQTL signals that colocalized with 4 *cis*-eQTL signals for its known regulator *NLRC5;* for all four signal pairs, the allele associated with increased *NLRC5* expression associated with increased *BTN3A3* expression, consistent with NLRC5’s role in activating *BTN3A3* (Dang et al., 2020).

Only one multi-colocalizing gene pair was observed in a tissue other than whole blood. The GTEx Consortium previously reported an *ENOX1* cis-eQTL colocalization with a *COL5A1* trans-sQTL in lung tissue. TOPMed lung results support and extend these previous findings: we find two *ENOX1* cis-eQTL signals colocalized with two *COL5A1* trans-sQTLs (Fig S48).

### GWAS signals frequently colocalize with secondary e/sQTL signals

To assess the utility of this dataset to identify target genes and nominate causal variants for GWAS signals, we colocalized all TOPMed cis- and trans-e/sQTL signals with the 34,107 fine-mapped EUR UK BioBank GWAS signals from 164 traits (Table S11). We additionally ran cis- and trans-e/sQTL scans on EUR subsets of the RNA-seq data (Fig S49A,B; Table S13) in order to obtain ancestry-matched e/sQTL results for colocalization with the EUR GWAS signals. Because the goal of e/sQTL - GWAS integration is commonly to interpret the GWAS signals, here we examine how many GWAS signals colocalize with at least one e/sQTL signal, rather than the inverse.

10,611 GWAS signals (31.1%) colocalized with at least one cross-ancestry cis-/trans-e/sQTL in at least one tissue (SuSiE-coloc PP4 posterior probability of colocalization >= 0.8; Fig 4A,B; 94.8% of colocalized GWAS signals colocalized with only cis signals, 1.7% with only trans signals, and 3.6% with at least one cis and at least one trans signal). 9,410 GWAS signals (27.6%) colocalized with at least one EUR cis-/trans-e/sQTL (Fig S49C), suggesting that in this case the sample size advantage of the cross-ancestry e/sQTL scans outweighed the advantage of ancestry-matching. However, we also observed 887 GWAS signals that colocalized with EUR e/sQTL but not cross-ancestry e/sQTL. We highlight the GWAS colocalizations with cross-ancestry e/sQTL here.

**Figure 4.**
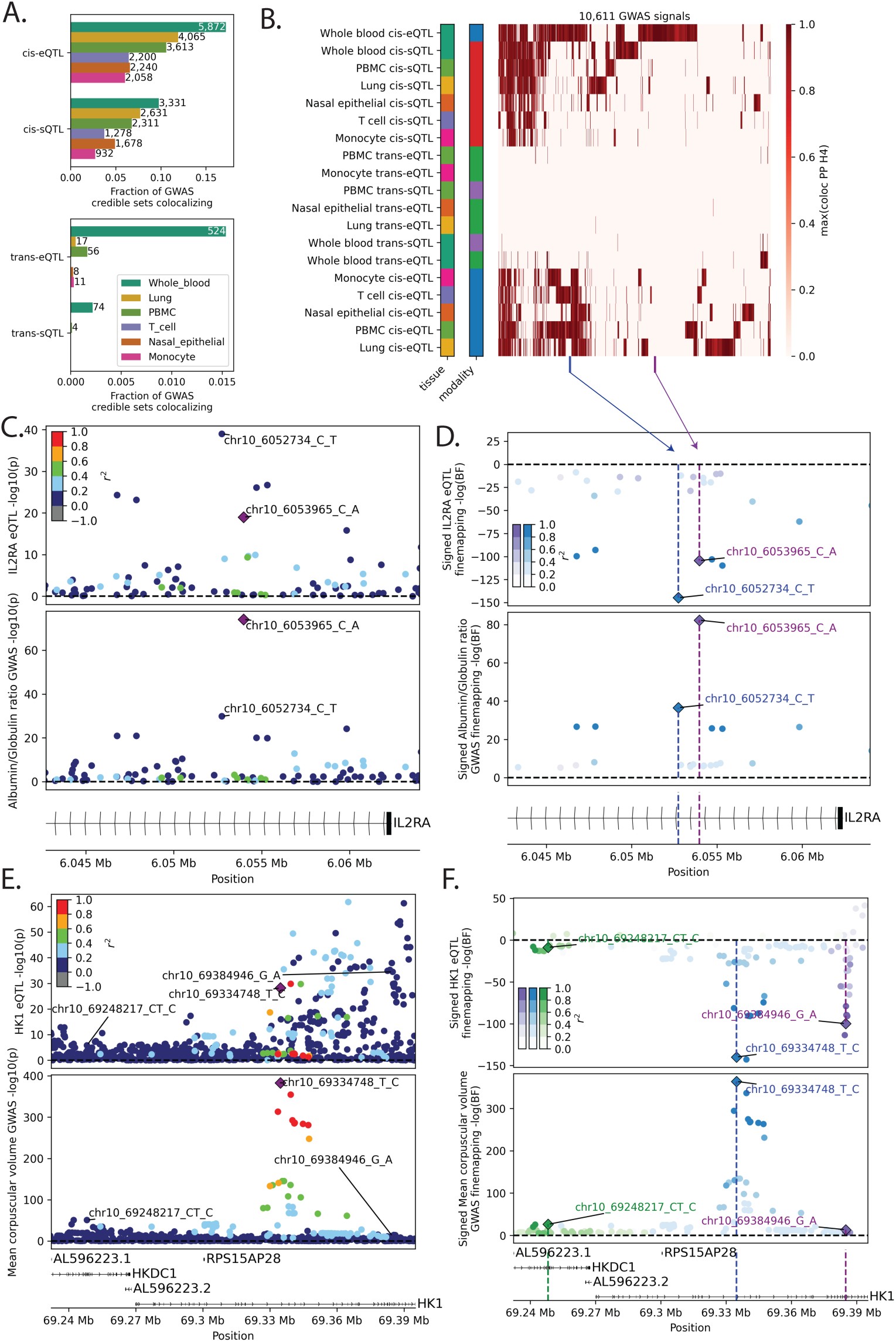
Colocalization of e/sQTL with UKBB GWAS signals. (A) Number of GWAS signals colocalizing with at least one e/sQTL credible set from each tissue and modality (cross-ancestry e/sQTL scans). (B) Heatmap displaying, for each GWAS signal with at least one e/sQTL colocalization, the maximum coloc posterior probability of colocalization for each tissue and modality. (C,D) Two *IL2RA* cis-eQTL signals colocalize with two albumin/globulin ratio GWAS signals. Marginal p-values are displayed in (C), and log Bayes factors for each of the two colocalizing e!ects, represented by the two colors, are displayed in (C); for the eQTL panel, the sign of each variant reflects the direction of e!ect on the gene’s expression for the GWAS trait-increasing allele in the colocalizing GWAS e!ect. (E,F) Three *HK1* cis-eQTL signals colocalize with three mean corpuscular volume GWAS signals.

GWAS traits with the greatest fraction of GWAS signals colocalizing with an e/sQTL signal generally corresponded to tissue relevant traits; for example, monocyte QTL signals were particularly likely to colocalize with ‘Monocyte count’ GWAS signals (Fig S50). In whole blood, 5,872, 3,331, 524, and 74 GWAS signals colocalized with cis-eQTL, cis-sQTL, trans-eQTL, and trans-sQTL signals, respectively, and thousands of GWAS signals colocalized with multiple e/sQTL types, especially cis-eQTL and cis-sQTL (Fig S51). 41.9% of the 10,611 GWAS signals colocalizing with at least one cross-ancestry e/sQTL signal colocalized with a signal from more than one gene (across all tissues and modalities) (Fig S52). In whole blood, 44.6% of GWAS signals with a cis-e/sQTL colocalization colocalized only with cis-e/sQTL signal(s) from the nearest tested gene; when the colocalization was only with cis-sQTL signals, the colocalization was more likely to involve the nearest gene (relative to colocalizations with cis-eQTL signals only; Fig S53). 2,876 of the 10,611 GWAS signals (27.1%) colocalized only with secondary signals (Fig S54) and a majority (7,096.0; 66.9%) colocalized with at least one secondary signal, emphasizing the importance of secondary e/sQTL signals in nominating effector genes for GWAS hits.

We identified 659 instances in which multiple neighboring GWAS signals for a given trait colocalized with multiple e/sQTL signals from the same gene and same e/sQTL type (Table S14). For example, two cis-eQTL signals at the *IL2RA* locus colocalize with two GWAS signals for albumin/globulin ratio (Fig 4C,D; S55). Both cis-eQTL signals are single-variant credible sets (chr10_6052734_C_T; chr10_6053965_C_A), and the signals are in a single *IL2RA* intron. The sequence surrounding chr10_6052734_C_T was previously shown via a CRISPR activation screen to regulate *IL2RA* expression (Simeonov et al., 2017). Three cis-eQTL signals for *HK1* colocalized with three GWAS signals for mean corpuscular volume (red blood cell volume; Fig 4E,F; S56). *HK1* is critical to red blood cell function; mutations in *HK1* are known to cause nonspherocytic haemolytic anemia (Jamwal et al., 2019; van Wijk et al., 2003). In the most extreme case, four ‘Monocyte count’ GWAS signals colocalized with four whole blood *CEBPB* cis-eQTL signals (Fig S57); *CEBPB* regulates monocyte development, survival, and gene expression (Huber et al., 2012; Mildner et al., 2017; Tamura et al., 2017), though these GWAS signals also colocalized with e/sQTL signals for other nearby genes (all four colocalized with at least one *SMIM25* cis-e/sQTL in at least one TOPMed tissue, including two colocalizing with two whole blood *SMIM25* cis-e/sQTLs; and three colocalized with *LINC01270* cis-eQTLs), suggesting that *CEBPB* may not be the target gene, or only target gene, at this locus.

### e/sQTL tissue diversity and sample size both contribute to GWAS colocalization analyses

Many GWAS signals have no known e/sQTL colocalization. One explanation for this is that they affect a trait via a different molecular mechanism. Alternatively, the GWAS signal might correspond to an e/sQTL signal that is highly specific to a tissue, timepoint, or context, or the GWAS signal might correspond to an e/sQTL signal that is too weak to be detected in a dataset. To explore the relative impact of tissue type breadth vs. sample size in detecting e/sQTL colocalizations with GWAS signals, we colocalized GTEx autosomal cis-e/sQTL signals from 49 tissues with the PanUKBB GWAS signals and compared the results to TOPMed autosomal cis-e/sQTL colocalization results (Fig S58). 12,033 GWAS signals colocalized with at least one GTEx cis-e/sQTL signal, compared to 10,208 colocalizing with at least one TOPMed cis-e/sQTL signal (MAF >= 0.01 scans). GWAS signals colocalizing with GTEx but not TOPMed cis-e/sQTL tended to colocalize in a smaller number of GTEx tissues than those colocalizing with both GTEx and TOPMed cis-eQTL signals, suggesting that these reflected more tissue-specific effects (Fig S58B) that might not be present in the more restricted TOPMed tissue set. In whole blood (GTEx N = 670; TOPMed N = 6,454), 2,897 GWAS signals colocalized with a GTEx cis-e/sQTL signal compared to 7,091 with a TOPMed cis-e/sQTL, and in lung (GTEx N = 515; TOPMed N = 1,291), these values were 3,142 and 5,381 for GTEx and TOPMed, respectively. GWAS signals colocalizing with TOPMed but not GTEx whole blood cis-e/sQTL signals tended to colocalize with weaker cis-eQTL signals than those colocalizing with both TOPMed and GTEx whole blood cis-e/sQTL signals (Fig S58C). These results demonstrate that tissue breadth as well as sample size have a considerable impact on colocalization detection.

To determine if the number of GWAS signals colocalizing with a whole blood eQTL signal has already reached saturation in our dataset, we colocalized the PanUKBB GWAS signals with the cis-eQTLs identified using nested subsets of the whole blood data (Fig S59). We found that the number of colocalizing GWAS signals continues to increase up to the current sample size, though at an increasingly slow pace. This suggests that GWAS signals not colocalizing with eQTL signals at the current sample size may colocalize in even larger eQTL datasets in the tested tissues.

## Discussion

Here we present cis- and trans-e/sQTL results from six tissues and cell types profiled as part of the TOPMed program. The uniform application of modern genotyping and gene expression profiling platforms (WGS and RNA-seq), combined with large sample sizes and individual-level data (as opposed to a meta-analysis), enables fine mapping of tens of thousands of secondary e/sQTL signals with high resolution.

Our results complement those in previous studies, most notably GTEx, DIRECT, and eQTLGen. For example, in whole blood we detect 69,766 total cis-eQTL signals for 19,468 genes, 51,037 (73.2%) of which were not detected in GTEx, DIRECT, or eQTLGen blood (see Methods). Of these 51,037, 2,760 (5.4%) colocalize with at least one PanUKBB GWAS signal, demonstrating the utility of the TOPMed data in interpreting GWAS signals. For instance, one PanUKBB GWAS signal for “Blood clot, DVT, bronchitis, emphysema, asthma, rhinitis, eczema, allergy diagnosed by doctor” has lead variant rs7936323 (chr11_76582714_G_A), and this variant and variants in high LD have previously been associated with allergy, asthma, and immune traits (Asai et al., 2018; Astle et al., 2016; Ferreira et al., 2017). The signal is intergenic, and previous publications have cited *LRRC32* and *EMSY* as candidate target genes (Elias et al., 2019). The *LRRC32* gene encodes the glycoprotein A repetitions predominant (GARP) protein, which helps regulate regulatory T cell activity and transforming growth factor β (TGF-β) release from T cells; mutations in *LRRC32* have previously been linked to immune disorders in humans and mice (Lehmkuhl et al., 2021; C. H. Wallace et al., 2018). *EMSY* is involved in transcriptional regulation including regulation of interferon stimulated genes (Ezell et al., 2012). This GWAS signal does not colocalize with any GTEx cis-e/sQTL signals. In TOPMed, we identify a colocalization between this GWAS signal and a *LRRC32* cis-eQTL (a four variant credible set including rs7936323 (chr11_76582714_G_A), with PIP = 0.31). The colocalization is not discovered using GTEx cis-e/sQTL credible sets, and the TOPMed cis-eQTL signal does not overlap DIRECT *LRRC32* conditional cis-eQTLs (or their R^2^ >= 0.8 LD proxies) or the eQTLGen *LRRC32* primary cis-eQTL or its R^2^ >= 0.8 LD proxies. Our results, especially in whole blood, therefore complement these previous studies.

In regards to trans-e/sQTL discovery, we discover substantially more whole blood trans-eGenes (1,725) than GTEx (thirteen), a similar number as DIRECT (1,670) but fewer trans-eGenes than studies that test a more restricted set of variants (e.g., eQTLGen detected 6,298 trans-eGenes, but tested only 10,317 trait associated variants) or use a more expansive definition of trans (Liu et al., 2022; Yao et al., 2017). GTEx lung and whole blood trans-eQTLs partially replicated in TOPMed (2/3 GTEx lung trans-eQTLs; 7/13 whole blood trans-eQTLs); 2/3 GTEx lung or whole blood trans-sQTLs replicated in TOPMed (Supplementary methods; Table S15; Table S16). Trans analyses are underpowered and prone to artifacts, leading to lower overlap in trans-eGenes between studies than one might expect. Only 78.1% and 26.9% of TOPMed whole blood trans-eGenes are trans-eGenes in eQTLGen and DIRECT, respectively (58.0% of DIRECT trans-eGenes are trans-eGenes in eQTLGen). While some of this is attributable to basic differences in testing procedures, this limited overlap also reflects the difficulty of these analyses. Nevertheless, ∼85% and ∼80% of eQTLGen and DIRECT primary trans-eQTLs showed the same direction of effect in TOPMed (regardless of statistical significance). Using the π_1_ statistic (Storey & Tibshirani, 2003) we estimate that the proportions of primary eQTLGen and DIRECT trans-eQTLs that replicate in TOPMed are 0.653 and 0.632, respectively.

Genomic windows with relatively large numbers of TOPMed trans-eQTLs showed elevated density of primary eQTLGen trans-eQTLs (Fig S60), and TOPMed trans-eVariants shared across many trans-eGenes frequently overlapped previously-established trans-eQTL hotspots (Fig S61) (Yao et al., 2017). Our top trans-eVariants (ranked by number of corresponding trans-eGenes) closely mirrored findings in previous studies; for example, our top trans-eVariant, rs946588154 (chr7_50342615_A_G; a trans-eVariant for 260 genes) is in perfect LD (1000G EUR populations based on LDlink) with rs149007767 (chr7_50330658_C_T) (Machiela & Chanock, 2015), which was a trans-eQTL hotspot in eQTLGen (top associated SNP for 522 eQTLGen trans-eGenes) and DIRECT. DIRECT and TOPMed both nominate *IKZF1* and *GRB10* as associated cis-eGenes that may be mediating the trans effect (Brown et al., 2023). The second highest number of trans signals was observed for rs1354034 (chr3_56815721_T_C), with 82 trans-eGenes. This was the top associated variant for 543 trans-eGenes in eQTLGen, and was a trans-eQTL in many additional studies (Kolberg et al., 2020; Mao et al., 2019; Nath et al., 2017). (Kolberg et al., 2020) identified the trans signal as highly specific to platelets. rs1354034 (chr3_56815721_T_C) is upstream of / intronic in gene *ARHGEF3*, for which it has been identified as a cis-eQTL in other studies (Kolberg et al., 2020; Zou et al., 2017). This variant also represents a protein QTL hotspot (Sun et al., 2023) and has been associated with a variety of blood cell traits, most commonly platelet volume and platelet count (Buniello et al., 2019; Chen et al., 2020; Gieger et al., 2011; J. Li et al., 2013; Vuckovic et al., 2020).

We show that even with 6,454 samples, we have not saturated cis-/trans-eQTL discovery. Considered alongside the fact that many e/sQTL signals are tissue and cell-type specific (GTEx Consortium, 2020), this suggests that many additional cis- and trans-e/sQTL signals remain to be discovered with greater sample sizes in a large array of tissues. While e/sQTLs are often employed to help interpret GWAS signals, GWAS hits and eQTLs face different selective constraints, so the overlap between the two is often limited (Mostafavi et al., 2023). This highlights the importance of larger QTL studies that are powered to detect more subtle/secondary effects. While the current study improves upon past studies in this regard, with still greater sample sizes (in a wider array of tissues) we anticipate that many as-yet-undiscovered e/sQTL signals will colocalize with GWAS signals.

Trans-e/sQTL signals have many potential explanations. Previous studies have found significant overlap between trans and cis-eQTL signals (GTEx Consortium, 2020; Yao et al., 2017), suggesting that trans-e/sQTL signals might be mediated by cis-e/sQTLs. Other potential (non-mutually exclusive) underlying explanations include direct changes to a protein sequence (for example, a missense variant in a transcription factor which alters the expression of that TF’s target genes); changes in post-transcriptional regulation (for example, a change in a TF gene’s 3’ or 5’ UTR that changes the efficiency of the TF’s mRNA translation and thereby affects the expression of the TF target genes) (Leppek et al., 2018; Steri et al., 2018); changes in cell type composition; and changes in a trait that in turn affects gene expression. We find that most trans signals (at current sample size) also have cis effects, suggesting they may be explainable by cis mediation. Larger analyses are needed to evaluate if this remains true as power to detect trans effects improves and to further characterize functional enrichment signals for trans-eVariants, which is challenging with the limited number of currently detected signals. A substantial fraction of trans signals (36.1% of whole blood trans-eQTLs) colocalize with GWAS signals for blood cell type abundance traits (e.g., neutrophil count); in some cases the trans signal may drive the GWAS signal, but in other cases the causality may be reversed (the change in cell type abundance may cause genes with differential expression across cell types to appear to be trans-eGenes). While the use of gene expression PCs should correct for cell type proportions, future single cell RNA-seq e/sQTL analyses are needed to fully dissect these effects. Future work could also incorporate cell type proportion estimates to improve power and to allow testing of cell type interaction effects, which can enable the detection of cell type specific QTLs. Careful examination of appropriate single cell reference panels representative of TOPMed participants and cell type estimation methods will be important for such future work. (Kim-Hellmuth et al., 2020)

While the TOPMed samples for several tissues were collected from more ancestrally diverse cohorts than previous studies, European ancestry donors are still over-represented in this study relative to other populations. As our analysis and other TOPMed eQTL studies have shown (Araujo et al., 2023; Kachuri et al., 2023), greater ancestral diversity will enhance our ability to detect eQTL/sQTL variants which are rare in European ancestry populations but common in other populations. Similarly, cohort-level demographic differences limit some of our conclusions; for example, overlap between whole blood and lung cis-eQTL signals may be affected by the notable ancestry, age, and disease phenotype differences between the donors in the contributing TOPMed cohorts.

While several TOPMed cohorts have longitudinal data (e.g., MESA), the current study does not attempt to utilize this. Future studies could examine changes in e/sQTL behavior over time, e.g. interactions between e/sQTL and age. Because metadata such as diet or medications is often limited, we do not examine any context-specific effects. Context-specific e/sQTLs are an area of increasing interest and have been proposed as one explanation for limited eQTL - GWAS overlap (Soskic et al., 2022).

In summary, we show that while many e/sQTL signals surely remain to be discovered, the joint analysis of TOPMed RNA-seq and WGS data defines e/sQTL signals at exceptionally high resolution, generating a useful resource for those studying gene expression and the effects of human genetic variation.

## Methods

### RNA-seq library preparation and sequencing

#### Gene-Environments and Admixture in Latino Asthmatics (GALA II) and Study of African Americans, Asthma, Genes, & Environments (SAGE)

Peripheral blood samples were collected into PAXgene Blood RNA tubes (PreAnalytiX, Hombrechtikon, Switzerland) and stored in the PAXgene tube in a −80°C freezer.

Total RNA was isolated from a PAXgene tube using a MagMAX for Stabilized Blood Tubes RNA Isolation Kit (4452306; Applied Biosystems). Globin depletion was performed using GLOBINclear Human (AM1980; Thermo Fisher Scientific). RNA integrity and yield were assessed using an Agilent 2100 Bioanalyzer (Agilent Technologies).

Total RNA was quantified using the Quant-iT RiboGreen RNA Assay Kit and normalized to 5 ng/µl. An aliquot of 300 ng for each sample was transferred into library preparation, which was an automated variant of the Illumina TruSeq Stranded mRNA Sample Preparation Kit. This method preserves strand orientation of the RNA transcript. It uses oligo dT beads to select messenger RNA from the total RNA sample. It is followed by heat fragmentation and complementary DNA synthesis from the RNA template. The resultant complementary DNA then goes through library preparation (end repair, base A addition, adapter ligation and enrichment) using Broad-designed indexed adapters substituted in for multiplexing. After enrichment, the libraries were quantified with quantitative PCR using the KAPA Library Quantification Kit for Illumina Sequencing Platforms and then pooled equimolarly. The entire process was in 96-well format and all pipetting was done using either an Agilent Bravo or a Hamilton Starlet instrument. Pooled libraries were normalized to 2 nM and denatured using 0.1 N NaOH before sequencing. Flow cell cluster amplification and sequencing were performed according to the manufacturer’s protocols using the HiSeq 4000. Each run was a 101-base pair paired-end read with an eight-base index barcode. Each sample was targeted to 50 million reads. Data were analyzed using the Broad Picard Pipeline, which includes demultiplexing and data aggregation.

#### COPDGene Study (COPDGene)

Whole blood samples were collected into PAXgene Blood RNA tubes (PreAnalytiX, Hombrechtikon, Switzerland) at the second study visit (five-year follow-up). Total RNA was extracted from PAXgene™ Blood RNA tubes using the Qiagen PreAnalytiX PAXgene Blood miRNA Kit (Qiagen, Valencia, CA). The extraction protocol was performed either manually or with the Qiagen QIAcube extraction robot according to the company’s standard operating procedure. Extracted RNA samples with a minimum RIN > 6 and concentration > =10 μg/ul were sequenced.

Initial QC entailed RNA quantification using the Quant-iT RNA assay (Invitrogen) and RNA integrity analysis using a fragment analyzer (Advanced Analytical). Samples were failed if the total amount, concentration, or integrity of RNA was too low.

Poly-A selection and cDNA synthesis were performed using the TruSeq Stranded mRNA kit as outlined by the manufacturer (Illumina). All steps were automated on the Perkin Elmer Sciclone NGSx Workstation to reduce batch to batch variability and to increase sample throughput. Final RNASeq libraries were quantified using the Quant-it dsDNA High Sensitivity assay, and library insert size distribution was checked using a fragment analyzer (Advanced Analytical). Samples where adapter dimers constitute more than 3% of the electopherogram area were failed prior to sequencing.

Sequencing was carried out on the NovaSeq sequencer. The processing pipeline consisted of the following elements: (1) base calls generated in real-time on the NovaSeq6000 instrument (RTA 3.1.5); (2) demultiplexed, unaligned BAM files produced by Picard ExtractIlluminaBarcodes and IlluminaBasecallsToSam were converted to FASTQ format using SamTools bam2fq (v1.4); (3) sequence read and base quality were checked using the FASTX-toolkit (v0.0.13).

#### Multi-Ethnic Study of Atherosclerosis (MESA)

MESA data was generated in two phases (a small pre-pilot phase and a pilot phase) and across two sequencing centers (at the Northwest Genomics Center (NWGC) and at the Broad Institute of MIT and Harvard).

##### Pre-pilot phase

The pre-pilot included peripheral blood mononuclear cells (PBMCs) isolated at the University of Vermont from Exam 1 (2000-2002) or at Wake Forest University from Exam 5 (2010-2012).

Three separate sets of PBMC RNA samples were used in the pre-pilot phase:

1. 32 MESA Exam 1 cryopreserved PBMC samples processed at the University of Vermont (taken from −145C freezers, thawed and RNA extracted using a Trizol Protocol described below). 16 samples were sent to the Broad and 16 to NWGC for RNA-seq library preparation and sequencing.
2. 40 MESA Exam 5 PBMC samples processed at Wake Forest University. These underwent automated extraction of RNA using the Qiagen AllPrep DNA/RNA Mini Kit (Catalogue # 80204) and the QIAcube. 20 samples were sent to the Broad and 20 to the NWGC for library preparation and sequencing.
3. Non-MESA control samples from the University of Vermont (none of which were used in e/sQTL scans). These were prepared from five donors, with three types of samples prepared from each donor: (1) fresh PBMC samples (RNA extracted from Paxgene collection tube following the Paxgene protocol); (2) past PBMCs (PBMCs prepared immediately, but frozen with RNAlater (Millipore-Sigma #R0901) overnight at −20C, then thawed with RNA extracted using the Qiagen AllPrep DNA/RNA Mini Kit (Catalogue # 80204)); and (3) cryopreserved PBMCs (cryopreserved PBMCs were prepared with no additive, then step frozen to −145°C, thawed, and RNA prepared, blood collected in Cell Preparation Tube/citrate (BD Vacutainer CPT #362761). RNA was extracted using the Qiagen AllPrep DNA/RNA Mini Kit (Catalogue # 80204). For each of these three preparations, RNA obtained from volunteers 1-3 had 250ng and was sent to both the NWGC and the Broad Institute, while RNA obtained from volunteers 4 and 5 had 500ng and was sent to both the NWGC and the Broad Institute.

The pre-pilot additionally included a small number of whole blood samples, which were not analyzed in this work.

##### Pilot phase

Exam 1 RNA was extracted at the University of Vermont from cryopreserved PBMCs using the Trizol protocol described below. RNA of all Exam 5 samples (PBMCs, monocytes and T cells) were extracted at Wake Forest using the QIAcube and the Qiagen AllPrep DNA/RNA Mini Kit (Catalogue # 80204). The Wake Forest laboratory shipped the extracted RNA samples to the University of Vermont for assignment of Pilot Study ID and plating; the University of Vermont shipped all plates containing RNA to the TOPMed Sequencing Centers. If a MESA participant had more than one RNA sample (i.e., Exam 1 PBMC, Exam 5 PBMC, Exam 5 monocyte, and/or Exam 5 T cell), these samples were aliquoted to the same 96-well plate. Frozen samples were shipped overnight in batches on dry ice.

##### Trizol RNA isolation protocol

This protocol requires the Qiagen RNeasy Mini Kit Cat No. 74104.

First, cells were thawed quickly at 37°C for ∼2 minutes, then pelleted by centrifuging at 500 x g for 5 minutes (2400 rpm) after which the freezing media was decanted.

For the lyse/phase separation, 1 mL TRIzol reagent was added and the mixture was vortexed. This was incubated at room temperature for 5 minutes. 0.2 mL cholorform was added, the tube was shaken vigorously by hand for 15 seconds, and this was incubated at room temperature for 2-3 minutes. The sample was then centrifuged at 12000 x g for 15 minutes at room temperature. 600 ul of the upper phase was transferred to a fresh RNase-free tube, 1.5x (∼900 ul) volume of 70% ethanol was added and the tube was inverted to mix

For binding, washing, and elution, 700 ul of the sample at a time was transferred to an RNeasy spin cartridge. This was centrifuged at 12000 x g for 15 seconds at room temperature and flow-through was discarded; this spin step was repeated until all of the sample had been processed. Next, 350 ul RW1 was added and this mixture spun at 12000 x g for 15 seconds and flow-through was discarded. Then, 500 ul RPE buffer was added, and this was spun for 15 seconds at full speed and flow-through discarded. Another 500 ul RPE buffer was added, and this was spun for 2 minutes at full speed and flow-through discarded. Lastly 30 ul RNase-Free Water was added to the center of cartridge, and this was incubated for 1 minute at room temperature before spinning for 2 minutes at max speed.

##### Broad library preparation and sequencing

Quantification of total RNA was accomplished using the Quant-iT^TM^ RiboGreen® RNA Assay Kit. Quality was assessed by measuring RNA degradation. Samples were assigned an RQS (RNA Quality Score) value based off of a 0-10 scale with 10 being the most intact RNA. To calculate the RQS, the Caliper LabChip GX was used to perform an electrophoresis-based separation to measure the fragment sizes of each sample. The RQS was calculated based on the 18S and 28S peak areas and heights of the resultant electropherogram, as well as the total RNA area. The calculation also took into consideration the FastRegion Area (the region between the LM and 18S peaks), which represents smaller, more degraded RNA fragments. After upfront quality assessment, samples were plated into diluted daughter aliquots with an input target of 250ng total RNA. Daughter concentrations were confirmed with RiboGreen quantification.

After quantification, 2 uL of External RNA Controls Consortium (ERCC) controls (using a 1:1000 dilution) were spiked into a 200ng aliquot of each sample destined for library construction. These consist of a set of unlabeled, polyadenylated transcripts, 250 to 2000nt in length designed to be added to an RNA analysis experiment after sample isolation, allowing for control of several sources of variability including quality of the starting material, the level of cellularity and RNA yield, the platform employed, and batch to batch variability.

The 200ng aliquot for each sample was continued into library preparation which used an automated variant of the Illumina TruSeq^TM^ Stranded mRNA Sample Preparation Kit. Input RNA first underwent PolyA selection with the use of oligo-dT purification beads. For optimal purification, the polyA selection process was performed in two sequential rounds. The mRNA was subsequently fragmented and primed for first strand synthesis, while the beads were eluted and washed away. The first strand of cDNA was synthesized from the mRNA template using reverse transcriptase. To create double-stranded cDNA, the mRNA template was removed and the second strand synthesized using the first cDNA strand as a template. AMPure XP beads were used to purify the ds cDNA from the reaction mix. The resulting product was blunt-ended cDNA. The 3’ blunt ends of the ds cDNA were subsequently adenylated with a single ‘A’ nucleotide. This provided a complementary overhang for the ligation of adapters and prevented the cDNA fragments from ligating to each other during this ligation reaction, thereby reducing chimera formation. Molecular adapters were then ligated to the ends of the ds cDNA to serve as primers for PCR enrichment. Each adapter was a unique molecular barcode specific for each well location of the 96-well plate. After enrichment, samples were amplified using PCR and the cDNA libraries subsequently quantified using PicoGreen and then pooled in equimolarity. Pools were quantified using qPCR and then normalized to 2nM. Afterwards, pools were denatured using 0.1 N NaOH prior to sequencing to create single-stranded DNA to be loaded onto the sequencers.

Flowcell cluster amplification and sequencing were performed according to the manufacturer’s protocols using the HiSeq 4000. The runs were 101bp paired-end with an eight-base index barcode read. Data was analyzed using the Broad Picard Pipeline which includes de-multiplexing and data aggregation.

##### NWGC library preparation and sequencing

Total RNA submitted by the investigator was verified using the Quant-iT RNA Assay Kit (Invitrogen, cat# Q33140). The NWGC required a minimum of 225ng of RNA per sample. Samples with less than 225 ng of RNA were failed in quality control. RNA quality was measured using the RIN score as assessed by the Agilent 2100 Bioanalyzer (Agilent, Santa Clara, CA). The RIN software algorithm determines the RIN score by assessing the electropherogram, including the 18S and 28S peak heights. Lower RIN scores indicate greater degradation of the total RNA. The NWGC requires RNA with a RIN score >5.0. Samples with RIN scores below 5.0 were considered as quality control failures by the NWGC.

Total RNA was normalized to 5ng/ul in a total volume of 50ul on the Perkin Elmer Janus Workstation (Perkin Elmer, Hopkington, MA). Poly-A selection and cDNA synthesis were performed using the TruSeq Stranded mRNA kit as outlined by the manufacturer (Illumina, cat#RS-122-2103). All steps were automated on the Perkin Elmer Sciclone NGSx Workstation to reduce batch to batch variability and to increase sample throughput. Total RNA was subject to two rounds of poly-A selection through sequential binding of poly-A RNA to oligo d(T) beads and washing away of unbound RNA. Purified mRNA was then eluted from the beads, fragmented and randomly primed for first strand synthesis using the SuperScript III reverse transcriptase (Invitrogen, cat#18080085). The original RNA template was degraded and double stranded cDNA was made using the first strand of cDNA as a template. The resulting cDNA was purified using AMPure XP beads (Beckman Coulter, A63882).

Double stranded cDNA proceeded through a series of shotgun library steps using the TruSeq Stranded mRNA kit, as outlined by the manufacturer. Library molecules are adenylated (A-tailing) to accommodate the T overhang of the Illumina Truseq adapters. Full length adapters were then ligated to the cDNA fragments, followed by an AMPure XP cleanup to remove unligated adapters. The NWGC uses a dual indexing strategy designed to avoid index hopping and to uniquely identify each library. Using unique dual indexing when multiplexing samples as well as implementing perfect barcode matching during demultiplexing ensures that samples are not contaminated at low levels with mismatched reads. Adapter ligated ds cDNA molecules were amplified by 13 cycles of PCR and subjected to a final 1X AMPure XP cleanup to remove carry over primers. All library preparation steps were carried out on the Perkin Elmer Sciclone NGSx Workstation.

Final RNA-seq libraries were quantified using the Quant-it dsDNA High Sensitivity assay. Library insert size distribution was checked using the DNA1000 assay on the Agilent 2100 Bioanalyzer. Samples where adapter dimers constituted more than 4% of the electropherogram area were failed prior to sequencing. Successful libraries were normalized to 10nM for submission to sequencing.

Ninety-six normalized and indexed libraries were pooled together and denatured before cluster generation on a cBot. The 96-plex pools were loaded on eight lanes of a flow cell and sequenced on a HiSeq4000 using illumina’s HiSeq 4000 reagents kit (cat# FC-410-1001,1002). For cluster generation, every step is controlled by cBot. When cluster generation is complete, the clustered patterned flow cells are then sequenced with sequencing software HCS (HiSeq Control Software v3.4.0.38). The runs are monitored for %Q30 bases using the SAV (Sequencing Analysis Viewer). Using RTA 2 (Real Time Analysis 2 v2.7.7) ) the BCLs (base calls) were de-multiplexed.

#### Women’s Health Initiative (WHI)

The WHI RNA samples were collected from Long Life Study (LLS) participants as part of the LLS Blood Protocol using the PreAnalytiX PAXgene blood tubes. After collection in participant homes throughout the US, PAXgene tubes were the last of five tubes drawn from each participant, mixed carefully (inverted 8-10 times), kept at room temperature for a minimum of 2 hours post draw, and shipped overnight with cool packs to the Fred Hutch Specimen Processing Lab (SPL). Upon receipt at the SPL, techs froze the PAXgene tubes at −80 degrees C until they could be transferred to the Fred Hutch Public Health Sciences Biomarker Lab, where the vials were kept frozen at −80 degrees C until a sufficient number for an extraction run was assembled.

Within about a month of collection, the lab extracted total RNA, including miRNA, using the PreAnalytiX method (PAXgene Blood miRNA Kit Handbook, Qiagen, 05/2009) designed for use with the PAXgene blood collection tubes. A qualitative assessment by agarose gel electrophoresis of RNA integrity was done at the time of extraction. The RNA was quantified by NanoDrop.

The elution volume of 76 µL of extracted RNA was divided between two RNA ‘Parent’ vials without further dilution, frozen at −80 degrees C, and shipped overnight on dry ice to the WHI biorepository for long term storage at −80 degrees C.

Extracted RNA was sent to the Northwest Genomics Center (NWGC) for library prep and sequencing.

Initial QC entailed RNA quantification using the Quant-iT RNA assay (Invitrogen) and RNA integrity analysis using a fragment analyzer (Advanced Analytical). Samples were failed if the total amount, concentration, or integrity of RNA was too low.

Poly-A selection and cDNA synthesis were performed using the TruSeq Stranded mRNA kit as outlined by the manufacturer (Illumina). All steps were automated on the Perkin Elmer Sciclone NGSx Workstation to reduce batch to batch variability and to increase sample throughput. Final RNASeq libraries were quantified using the Quant-it dsDNA High Sensitivity assay, and library insert size distribution was checked using a fragment analyzer (Advanced Analytical). Samples where adapter dimers constitute more than 3% of the electropherogram area were failed prior to sequencing.

Sequencing was carried out on the NovaSeq sequencer. The processing pipeline consisted of the following elements: (1) base calls generated in real-time on the NovaSeq6000 instrument (RTA 3.1.5); (2) demultiplexed, unaligned BAM files produced by Picard ExtractIlluminaBarcodes and IlluminaBasecallsToSam were converted to FASTQ format using SamTools bam2fq (v1.4); (3) sequence read and base quality were checked using the FASTX-toolkit (v0.0.13).

#### Lung Tissue Research Consortium (LTRC)

The lung tissue samples were aliquoted and stored at −80 °C. Reagents and components of the Qiagen Allprep DNA/RNA/miRNA Universal Kit (Qiagen, Catalog # 80224) were used for the simultaneous extraction of DNA and RNA from the Lung Tissue samples. The extractions were performed at the Channing Division of Network Medicine at the Brigham and Women’s Hospital. 10-20 mg of the frozen Lung Tissue samples were weighed using sterile scalpel under frozen conditions. The extractions were performed as per the kit manufacturer’s instructions. The RNA was eluted in TE (Tris EDTA, pH=7.0) buffer. The eluted RNA was stored in NUNC cryotubes (Fisher Scientific, cat # 12-565-170N) at −80 ° C.

RNA samples were randomized into 16 plates. The RNA samples were quantified using RiboGreen dye (Thermo Fisher, catalog # Quant-iT P 7581). Minimum 12 μl of DNA sample was plated in 96 well plate (0.8 mL deep well plate by ABgene, catalog # AB- 07565) to get minimum RNA mass of 500 ng/ sample.

Extracted RNA was sent to the Northwest Genomics Center (NWGC) for library prep and sequencing.

At the NWGC, initial QC entailed RNA quantification using the Quant-iT RNA assay (Invitrogen) and RNA integrity analysis using a fragment analyzer (Advanced Analytical). Samples were failed if the total amount, concentration, or integrity of RNA was too low.

Total RNA was normalized to 7.5ng/ul in a total volume of 50ul on the Perkin Elmer Janus Workstation (Perkin Elmer, Janus II). Poly-A selection and cDNA synthesis were performed using the TruSeq Stranded mRNA kit as outlined by the manufacturer (Illumina, cat#RS-122-2103). All steps were automated on the Perkin Elmer Sciclone NGSx Workstation to reduce batch to batch variability and to increase sample throughput. Final RNASeq libraries were quantified using the Quant-it dsDNA High Sensitivity assay, and library insert size distribution was checked using a fragment analyzer (Advanced Analytical; kit ID DNF474). Samples where adapter dimers constitute more than 4% of the electopherogram area were failed prior to sequencing. Technical controls (K562,Thermo Fisher Scientific, cat# AM7832) were compared to expected results to ensure that batch to batch variability was minimized. Successful libraries were normalized to 10nM for submission to sequencing.

Barcoded libraries were pooled using liquid handling robotics prior to loading. Sequencing was carried out on the NovaSeq sequencer.

The processing pipeline consisted of the following elements: (1) base calls generated in real-time on the NovaSeq6000 instrument (RTA 3.1.5); (2) demultiplexed, unaligned BAM files produced by Picard ExtractIlluminaBarcodes and IlluminaBasecallsToSam were converted to FASTQ format using SamTools bam2fq (v1.4); (3) sequence read and base quality were checked using the FASTX-toolkit (v0.0.13).

#### Framingham Heart Study (FHS)

Peripheral whole blood samples (2.5 mL) were collected from FHS participants using PAXgene™ tubes (PreAnalytiX, Hombrechtikon, Switzerland), incubated at room temperature for 4 h for RNA stabilization, and then stored at − 80 °C until use.

For one subset of samples, total RNA was isolated using a standard protocol using a PAXgene Blood RNA Kit at the FHS Genetics Laboratory. Tubes were allowed to thaw for 16 h at room temperature. Cell pellets were collected after centrifugation and washing. Cell pellets were lysed in guanidinium-containing buffer. The extracted RNA was tested for its quality by determining absorbance readings at 260 and 280 nm using a NanoDrop ND-1000 UV spectrophotometer. The Agilent Bioanalyzer 2100 microfluidic electrophoresis (Nano Assay and the Caliper LabChip system) was used to determine the integrity of total RNA. For the rest of the samples, total RNA was isolated at the TOPMed contract laboratory at Northwest Genomics Center.

Library prep and sequencing was performed at Northwest Genomics Center. Initial QC entails RNA quantification using the Quant-iT RNA assay (Invitrogen) and RNA integrity analysis using a fragment analyzer (Advanced Analytical). Samples are failed if the total amount, concentration, or integrity of RNA is too low.

Poly-A selection and cDNA synthesis are performed using the TruSeq Stranded mRNA kit as outlined by the manufacturer (Illumina). All steps are automated on the Perkin Elmer Sciclone NGSx Workstation to reduce batch to batch variability and to increase sample throughput. Final RNASeq libraries are quantified using the Quant-it dsDNA High Sensitivity assay, and library insert size distribution is checked using a fragment analyzer (Advanced Analytical). Samples where adapter dimers constitute more than 3% of the electopherogram area are failed prior to sequencing.

Sequencing is carried out on the NovaSeq sequencer. The processing pipeline consists of the following elements: (1) base calls generated in real-time on the NovaSeq6000 instrument (RTA 3.1.5); (2) demultiplexed, unaligned BAM files produced by Picard ExtractIlluminaBarcodes and IlluminaBasecallsToSam are converted to FASTQ format using SamTools bam2fq (v1.4); (3) sequence read and base quality are checked using the FASTX-toolkit (v0.0.13).

#### Subpopulations and Intermediate Outcome Measures In COPD Study (SPIROMICS)

RNA was extracted from PAXgene blood tubes using a magnetic-bead extraction method using the Perkin-Elmer (now Revvity) Magnetic Separation Module I (MSMI) extraction robotics system. The principle behind magnetic bead separation is that after cell lysis the nucleic acid binds to the beads, while the proteins and other unwanted materials do not bind. Multiple washes of the beads remove any non-specific binding of these unwanted materials.

After receipt of the samples into the UNC BioSpecimen Processing Facility the Paxgene blood tubes were kept at room temperature for a minimum of 4 hours to a maximum of 72 hours after which the tubes are frozen at −80°until extraction occurs, in multiple of 6 samples. If samples arrived frozen they were immediately placed into −80°C. Clinical sites were instructed to hold samples at room temperature prior to freezing for a minimum of 4 hours to a maximum of 72 hours. Prior to lysate preparation, extraction and placement on the MSMI system, frozen PAXgene blood tubes to be processed are removed from the freezer and incubated at room temperature overnight. After overnight incubation the blood from the PAXgene tubes are pelleted at 3250 x g for 10 minutes at 15°C. The cell pellets are washed 1 time after being resuspended in RNASE-free water. At this time the pellet was resuspended in 500ul RLT lysis buffer (Qiagen) containing 1% 2-ME, pipetted to break up the pellet and frozen at −80°C for a week before extraction on the MSMI using the chemagic™ PAXgene RNA Reagent Kit - CMG-1084. Prior to loading on the machine for extraction the prepared lysates were removed from the freezer and allowed to thaw after which 1.5ml of RNAse-free water was added to each pellet in 500ul aliquots vortex gently and then 50ul of proteinase K was added to each tube prior to loading onto the machine. Extraction was performed using “chemagic™ RNA Blood 24 drying VD100806.che” protocol with DNase. Samples were eluted into 155ul of MSMI elution buffer and were quantitated by UV absorbance on a Denovix reader followed by RNA quality analysis on an Agilent Tapestation.

Extracted RNA was sent to the Northwest Genomics Center (NWGC) for library prep and sequencing.

At the NWGC, initial QC entailed RNA quantification using the Quant-iT RNA assay (Invitrogen) and RNA integrity analysis using a fragment analyzer (Advanced Analytical). Samples were failed if the total amount, concentration, or integrity of RNA was too low.

Total RNA was normalized to 7.5ng/ul in a total volume of 50ul on the Perkin Elmer Janus Workstation (Perkin Elmer, Janus II). Poly-A selection and cDNA synthesis were performed using the TruSeq Stranded mRNA kit as outlined by the manufacturer (Illumina, cat#RS-122-2103). All steps were automated on the Perkin Elmer Sciclone NGSx Workstation to reduce batch to batch variability and to increase sample throughput. Final RNASeq libraries were quantified using the Quant-it dsDNA High Sensitivity assay, and library insert size distribution was checked using a fragment analyzer (Advanced Analytical; kit ID DNF474). Samples where adapter dimers constitute more than 4% of the electopherogram area were failed prior to sequencing. Technical controls (K562,Thermo Fisher Scientific, cat# AM7832) were compared to expected results to ensure that batch to batch variability was minimized. Successful libraries were normalized to 10nM for submission to sequencing.

Barcoded libraries are pooled using liquid handling robotics prior to loading. Sequencing was carried out on the NovaSeq sequencer.

The processing pipeline consisted of the following elements: (1) base calls generated in real-time on the NovaSeq6000 instrument (RTA 3.1.5); (2) demultiplexed, unaligned BAM files produced by Picard ExtractIlluminaBarcodes and IlluminaBasecallsToSam were converted to FASTQ format using SamTools bam2fq (v1.4); (3) sequence read and base quality were checked using the FASTX-toolkit (v0.0.13).

### RNA-seq mapping

RNA-seq reads were mapped to the GRCh38 reference genome. We used a GRCh38 reference fasta file that included ERCC spike-ins and excluded ALT, HLA, and decoy contigs, and used a collapsed GENCODE v30 gene annotation. The collapsed annotation was generated using a GTEx consortium script: https://github.com/broadinstitute/gtex-pipeline/commits/master/gene_model/collapse_annotation

.py, and ERCC annotations were appended.

Reads were mapped with STAR v. 2.6.1d (except for MESA, which used STAR v 2.5.3a with an index built from the GENCODE v26 gene annotation), using default parameters except --twopassMode Basic --outFilterMultimapNmax 20 --alignSJoverhangMin 8 --alignSJDBoverhangMin 1 --outFilterMismatchNmax 999 --outFilterMismatchNoverLmax 0.1 --alignIntronMin 20 –alignIntronMax 1000000 --alignMatesGapMax 1000000 --outFilterType BySJout --outFilterScoreMinOverLread 0.33 --outFilterMatchNminOverLread 0.33

--limitSjdbInsertNsj 1200000 --outSAMstrandField intronMotif --quantMode TranscriptomeSAM GeneCounts --outSAMtype BAM Unsorted --outSAMunmapped Within --chimSegmentMin 15 –chimJunctionOverhangMin 15 --chimOutType Junctions WithinBAM SoftClip --chimMainSegmentMultNmax 1 --outSAMattributes NH HI AS nM NM ch --outSAMattrRGline ID:rg1 SM:sm1

Duplicates were marked using picard (v. 2.18.17) MarkDuplicates with default parameters except ASSUME_SORT_ORDER=coordinate.

Gene counts, transcript per million (TPM) expression values, and QC metrics for each sample (including MESA samples) were computed using RNA-SeQC v. 2.3.3 (Graubert et al., 2021) with parameter --stranded rf, using the collapsed GENCODE v30 gene annotation.

### Matching RNA-seq samples to whole genome genotypes

For each RNA-seq sample, we run vt discover2 on the RNA-seq .bam file, restricting to reads with mapping quality >= 20, to identify candidate variant sites. The output from vt discover2 is converted to a crude diploid genotype at each site by approximately ’round(2 * AD / DP)’. These genotypes are compared to the genotypes for all samples in TOPMed freeze 9b, restricted to PASS variants with minor allele frequency (MAF) >= 0.05 TOPMed-wide in coding exons as defined by GENCODE v34. The comparison calculates non-reference genotype concordance as:

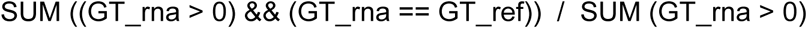

where the sums are over all sites with TOPMed-wide minor allele frequency >= 0.05 in coding exons, and integers GT_rna, GT_ref represent genotypes for the RNA-seq or a whole genome reference sample respectively, coded as 0, 1 or 2 non-reference alleles. The comparison to all sequenced samples in freeze 9b separates into three clusters: putative matches with high concordance (85% - 93%), putative matches to a related sample (55% - 65%), and then putative unmatched samples (40% - 50%). We declare an RNA-seq sample as unmatched if its highest genotype concordance with any sample in freeze 9b is below 83%.

For MESA cohort samples only, expected WGS - RNA matches are used and matches where the RNA-seq sample failed fingerprinting or expression-based sex check are dropped.

733 donors contributed data from more than one tissue or cell type (Fig S62), including 334 donors from the MESA cohort that contributed T cell, monocyte, and PBMC RNA-seq samples, and 324 donors from the COPDGene cohort that contributed nasal epithelial and whole blood samples.

### Glocal ancestry estimation

Local ancestry was inferred using RFMix v2 (Maples et al., 2013) with the following option: --node-size=5. For reference haplotypes used in local ancestry inference, we obtained the Human Genome Diversity Panel (HGDP) (J. Z. Li et al., 2008) and processed the data according to (C. Wang et al., 2014), ending up with 938 individuals and 639,958 autosomal SNPs. We then condensed the 53 populations in HGDP into 7 super-populations: (1) Sub-Saharan Africa (n=104), (2) Central/South Asia (n=200), (3) East Asia (n=229), (4) Europe (n=154), (5) Native America (n=63), (6) Oceania (n=28), (7) Middle East (n=160). After running

RFMix, we summed up inferred local ancestry across all genetic windows of each individual to calculate global ancestry proportions, corresponding to the seven super-populations. Local ancestry was not used in our analyses; global ancestry was used to describe donor ancestry / admixture and define population groupings as described below.

Genetic ancestry refers to segments of an individual’s genome that have been inherited from their ancestors. In practice we approximate this from some quantitative measure of genetic sharing between individuals in our study and reference populations. As such, genetic ancestry is an estimate that has some error and variability. <u>(Lewis et al., 2022; Mathieson & Scally, 2020)</u>.

### Assignment of individuals to population clusters

Unless otherwise specified, a sample was assigned to an ancestry if the sample’s estimated % global genetic ancestry for the ancestry in question was at least 75%. If a sample was < 75% for all ancestries (for example, 25% AFR, 25% AMR, and 50% EUR), it was left unassigned and omitted from the analysis; in some tissues, this left a substantial fraction of samples unassigned to an ancestry (e.g., 1,845 of 6,454 (28.6%) whole blood samples). The number of samples assigned to each ancestry in each tissue under these criteria are listed in Table S17.

Genetic variation is continuous and we acknowledge that imposing a discrete structure using somewhat arbitrary cut-offs is problematic <u>(Lewis et al., 2022; Mathieson & Scally, 2020)</u>. We selected global ancestry thresholds to create groupings for cross-population comparisons of e/sQTLs. These comparisons are intended to provide insight into genetic architecture of gene expression and the effects of varying demographic histories. Use of a broad global ancestry estimation also helps avoid the intrinsic challenges associated with the distinct population descriptors used between TOPMed studies and the demographic nuances of each study.

We selected a lower threshold (50%) for the AMR ancestry to increase participant inclusion in the related analyses, and because AMR reference panels include many individuals with significant admixture themselves (esp. EUR and AFR) <u>(The 1000 Genomes Project Consortium,</u> <u>2015)</u>.

### Genotypes

Genotypes for WGS samples matching an RNA-seq sample were extracted from the freeze 9b TOPMed BCF files (Danecek et al., 2021). Only autosomal + chrX SNPs and short indels (< 50 bp) were used.

### Selection of unrelated subjects

Autosomal SNPs with FILTER=PASS and MAF >= 1% were used for determining subject relatedness. Relatedness was calculated using KING v. 2.2.7 (Manichaikul et al., 2010) (options --degree 5 --related). An unrelated set of subjects for downstream analysis was generated from the KING output as follows. First, subjects were ranked according to the number of inferred relatives they had (related meaning 4th degree or greater relatedness). Next, the individual with the fewest relatives was added to the unrelated sample set, and any subjects that this individual was related to were dropped from further consideration. This step was iteratively performed until all samples had either been added to the unrelated sample set or had been dropped from the analysis.

### Genotype PCA

PCA was performed on the genotypes of the unrelated subjects, utilizing a set of LD pruned and thinned common (MAF >= 1%) SNPs. EIGENSOFT (Patterson et al., 2006; Price et al., 2006) (git commit 09ed563f) was used for the PCA, computing the top 15 PCs (`smartpca.perl` with options `-k 15 -m 0`). The genotype PCA is plotted in Fig S63.

### RNA-seq QC

No hard thresholds were set on individual QC metrics; a PCA-based approach was used to label samples as outliers or non-outliers, and outlier samples were excluded from e/sQTL scans. For each cohort/tissue combination, we performed DESeq2 size factor-based normalization of gene counts (Love et al., 2014) (as implemented in pyqtl (https://github.com/broadinstitute/pyqtl) function `deseq2_normalized_counts`), dropped genes for which fewer than 10% of samples had a normalized count of at least 10, filled zeros with a value equal to ½ of the minimum observed non-zero value for that gene, log_10_ transformed the matrix, and then performed PCA. A sample was labeled as an outlier in PC space if any of the following criteria were met:

● The sample’s Mahalanobis distance, computed with the top 5 PCs, corresponded to a chi-square p-value < 0.001
● Along any of the top 10 PCs, the sample’s absolute deviation from the median, normalized by the median absolute deviation across all samples for that PC, was >= 5.

### Selection of samples for e/sQTL scans

For each tissue, samples for the cis-eQTL scan were selected via the following procedure:

1). Exclude samples that were outliers in PC space
2). Exclude samples without a known WGS match
3). Exclude samples where the subject (WGS match) is not in the unrelated subject set.
4). Exclude samples with unclear sex based on gene expression. Sex was inferred using TPM values for genes XIST (on chrX) and RPS4Y1 (on chrY) (Fig S64).
5). Keep only one sample per subject. This was done randomly, except:

● For samples from the SPIROMICS cohort, samples from the baseline timepoint were preferred, if the subject had a sample from this timepoint.
● For samples from the MESA cohort, samples from exam 5 were preferred, if the subject had a sample from this timepoint.

### cis-eQTL scans

For each tissue, using the samples selected for inclusion in the cis-eQTL scan:

1). Gene counts were filtered to include only autosomal and chrX genes

2). Genes counts were normalized using the edgeR TMM procedure (Robinson et al., 2010), as implemented in pyqtl function `edger_cpm`.

3). Lowly expressed genes, defined as those where < 20% of samples have a TPM value of >

0.1, were dropped.

4). TMM-normalized gene expression values were inverse normal transformed.

To generate gene expression PCs to be used as covariates in the cis-eQTL scans, we performed PCA on the inverse normal transformed gene expression matrix, excluding genes with low mappability (mappability < 0.5, using mappability values from (Saha & Battle, 2018); genes without mappability scores were dropped prior to PCA as well).

Covariates for each tissue were:

● Whole blood: cohort + inferred sex + 10 genotype PCs + 100 gene expression PCs
● Lung: inferred sex + 10 genotype PCs + 75 gene expression PCs
● Nasal epithelial: inferred sex + 10 genotype PCs + 30 gene expression PCs
● T-cells: inferred sex + 10 genotype PCs + 30 gene expression PCs
● PBMCs: inferred sex + 10 genotype PCs + 30 gene expression PCs
● Monocytes: inferred sex + 10 genotype PCs + 30 gene expression PCs

SNPs and indels meeting the following criteria were tested:

● Marked as PASS in the TOPMed VCF files.
● MAF >= 1% in the scan samples (for whole blood, two sets of scans were run, one using MAF >= 1% and one using MAF >= 0.1%)

For each gene, genetic variants within 1Mb of the gene TSS were tested. Gene TSS locations were determined using pyqtl’s gtf_to_tss_bed function.

The number of gene expression PCs to use for each tissue was determined by examining the relationship between the number of cis-eGenes detected and the number of PCs used as covariates, and selecting the point at which the number of cis-eGenes began to level off (Fig S65).

cis-eQTL scans were performed using tensorQTL v. 1.0.7 (Taylor-Weiner et al., 2019), modified to add a second inverse normalization of the gene expression values following residualization against the scan covariates (modified code at https://github.com/porchard/tensorqtl/tree/5ea048f2705035df1cb87e59eb143a54805cadeb).

Permutations were used to identify eGenes (tensorQTL mode = cis; q-value lambda = 0 and seed = 2021). Full summary statistics were computed using tensorQTL mode cis_nominal. cis-eQTL signals for cis-eGenes were fine-mapped using the SuSiE (G. Wang et al., 2020) implementation in tensorQTL. For monocytes, T cells, and nasal epithelial samples, we set the SuSiE L parameter (number of non-zero effects to consider when fitting the SuSiE model) to 10. For lung, PBMCs, and whole blood, we ran SuSiE with multiple L values (10 and 20 for lung and PBMCs; 10, 20, and 30 for whole blood) and for each gene selected the minimum L greater than or equal to the maximum number of credible sets discovered across tested Ls. By default the SNP PIP values reported by SuSiE represent an aggregation across single effects; we therefore calculated single-effect PIP values using the Bayes factor matrix (PIP_ij = BF_ij / sum(BF_j), for SNP i and single effect j, where BF represents Bayes factors and the sum is across SNPs; in practice the difference between the single-effect PIP values and the PIP values reported by SuSiE tends to be extremely minor). A small minority of credible sets were duplicates (credible sets containing the same SNPs and PIP values); such duplicate credible sets were collapsed into single credible sets.

Cis-eQTL analyses on the subset of genetically-inferred EUR ancestry samples were performed in the same manner as full-sample cis-eQTL analyses, except for the number of phenotype PCs and SuSiE L parameters (listed in Table S18). The number of phenotype PCs was selected by examining the relationship between PCs used and eGenes discovered.

For some downstream analyses, we utilized only the top PIP variant per credible set. In the case that two variants had the same PIP (i.e., were in perfect LD), we arbitrarily selected one of them (the first one based on ascending alphabetical-order sorting).

### cis-sQTL scans

We quantified splicing using LeafCutter (Y. I. Li et al., 2018) intron excision ratios.

Prior to quantifying splicing, samples were remapped in a variant-aware manner using STAR (Dobin et al., 2012) v. 2.6.1d with WASP (van de Geijn et al., 2015) filtering (`--waspOutputMode SAMtag --varVCFfile $vcf --outSAMattributes NH HI AS nM NM ch vW`), otherwise using the same mapping parameters used in the initial STAR mapping. Reads were filtered to uniquely mapped reads passing WASP filtering (`samtools view -h -q 255 $bam | grep -v “vW:i:[2-7]”`) and exon-exon junction counts were computed using regtools (Cotto et al., 2023) v. 0.5.(Cotto et al., 2023) (`regtools junctions extract -a 8 -m 50 -M 500000 -s 1 filtered.bam`).

We used LeafCutter’s leafcutter_cluster_regtools.py to cluster introns (`--minclureads round(tissue N/5) --mincluratio 0.001 --maxintronlen 500000`), additionally modifying the procedure to exclude introns supported by fewer than (round(tissue N / 10)) samples. We mapped the intron clusters to genes using the map_clusters_to_genes.R GTEx (GTEx Consortium, 2020) script (implementing LeafCutter function map_clusters_to_genes).

We removed introns with no counts in the majority of the samples, and removed introns with low variability across samples using the intron cluster fraction Z-score filter introduced in (GTEx Consortium, 2020) (removing introns where three or fewer samples have abs(cluster fraction z-score) > 6 and no more than three samples have abs(cluster fraction z-score) > 0.25). The resulting matrix was normalized using LeafCutter’s `prepare_phenotype_table.py` script, and then each splicing phenotype was inverse normalized. This matrix was used as input to the cis- and trans-sQTL scans, and splicing PCs used as scan covariates were calculated on this matrix.

We used inferred sex, 10 genotype PCs, 10 splicing phenotype PCs, and (for whole blood) TOPMed cohort as scan covariates (Fig S66).

SNPs and indels meeting the following criteria were tested:

● Marked as PASS in the TOPMed VCF files.
● MAF >= 1% in the scan samples (for whole blood, two sets of scans were run, one using MAF >= 1% and one using MAF >= 0.1%)

For each gene, genetic variants within 1Mb of the gene TSS were tested. Gene TSS locations were determined using pyqtl’s gtf_to_tss_bed function.

cis-sQTL scans were performed using tensorQTL v. 1.0.7 (Taylor-Weiner et al., 2019), modified to add a second inverse normalization of the splicing phenotypes following residualization against the scan covariates (modified code at https://github.com/porchard/tensorqtl/tree/5ea048f2705035df1cb87e59eb143a54805cadeb).

Permutations were used to identify sGenes (tensorQTL mode = cis; q-value lambda = 0 and seed = 2021, grouping phenotypes by gene). To get gene-level cis-sQTL credible sets, we identified introns that have a variant association strong enough to pass genome-wide FDR 5% threshold (based on tensorQTL mode cis_nominal significant phenotype - variant pairs), fine-mapped cis-sQTL signals for each such intron using the SuSiE implementation in tensorQTL (SuSiE L = 10), and then, for each gene, collapsed credible sets across introns by identifying overlapping credible sets and keeping the credible set with larger max PIP.

Cis-sQTL analyses on the subset of genetically-inferred EUR ancestry samples were performed in the same manner as full-sample cis-sQTL analyses (including leafcutter phenotype generation), except for the number of phenotype PCs and SuSiE L parameters (listed in Table S18). The number of phenotype PCs was selected by examining the relationship between PCs used and sGenes discovered.

For some downstream analyses, we utilized only the top PIP variant per credible set. In the case that two variants had the same PIP (i.e., were in perfect LD), we arbitrarily selected one of them (the first one based on ascending alphabetical-order sorting).

### Determining primary vs non-primary signals

For any analyses performed prior to fine-mapping, ‘primary’ refers to the variant most strongly associated with the expression/splicing of each gene.

Following fine-mapping, to rank cis-eQTL signals for each gene as primary, secondary, tertiary, etc., for each gene with a fine-mapped signal we fit a single, joint eQTL model using the same covariates and phenotypes as in the cis-eQTL scan and including all of the top PIP variants for each of the gene’s credible sets. The variants were then ranked by their corresponding coefficient p-values from this model. Rarely, a single variant was the top PIP variant for more than one cis-eQTL credible set (2 such cases for lung cis-eQTL scans; 4 such cases for PBMC cis-eQTL scan; 73 and 92 such cases for MAF < 0.001 and MAF < 0.01 for whole blood cis-eQTL scans, respectively). In such cases, the relative order of the credible sets was determined based on the credible set ID (credible set ‘1’ being ranked above credible set ‘2’, etc).

The procedure for ranking cis-sQTL signals was the same, except that in the case that a gene’s cis-sQTL credible sets were derived from > 1 splicing phenotype, individual models were run for each of the splicing phenotypes to obtain the variant coefficient p-values before generating the gene-level credible set rankings.

In some analyses, e.g. functional enrichment analyses, e/sVariants were collapsed across genes so as not to be double-counted in the case that the same variant was an e/sVariant for > 1 gene. In such cases an e/sVariant might correspond to the primary signal for one gene but not another. If the e/sVariant was not either primary or non-primary for all related genes, it was excluded from that analysis.

### Gene and variant mappability and cross-mappability

Gene mappability, variant mappability, and cross-mappability between genes was calculated as described in (Saha & Battle, 2018) using a NextFlow implementation of their pipeline (available at https://github.com/porchard/crossmap-nextflow) and the uncollapsed GENCODE v30 GTF file, with exon kmer length set to 100 bps, UTR kmer length set to 36 bps, and allowing two mismatches.

### trans-eQTL scans

trans-eQTL scans were performed using the same normalized gene expression matrices and covariates as cis-eQTL scans, except we dropped some gene expression PCs from the covariate matrices to avoid adjusting out trans effects. To determine which gene expression PCs might capture trans effects and therefore should be dropped from the covariate matrix for the trans-eQTL scan, we used tensorQTL to test for an association between each variant and each gene expression PC, using the same covariates as in the cis-eQTL scan minus the gene expression PCs. The strongest association observed for each gene expression PC and each tissue is shown in Fig S67. For whole blood, PCs 3, 25, and many PCs beyond PC50 strongly (p < 1×10^-15^) associated with a genetic variant. The variant most strongly associated with PC3 was rs2814778 (chr1_159204893_T_C), which is the known causal variant for benign neutropenia and is associated with neutrophil percentage and lymphocyte percentage (Charles et al., 2018; D. Reich et al., 2009; Reiner et al., 2011); PC3 may therefore capture cell type abundance, and we elected to keep PC3 as a covariate. The variant most strongly associated with PC25 was chr3_56815721_T_C, which was previously identified as a trans-eQTL in several studies (Kolberg et al., 2020; Mao et al., 2019; Nath et al., 2017); we elected to exclude PC25 it as a covariate. In addition, we excluded whole blood gene expression PCs beyond PC50. No PCs were dropped for any other tissues.

All trans-eQTL scans tested variants with MAF >= 0.05. To reduce the probability of mapping artifacts, we removed variants with mappability < 1. To identify trans-eGenes and their respective trans-eVariants, we used tensorQTL mode --trans to test for associations between variant - gene pairs on separate chromosomes. Due to the large number of pairs tested, we saved summary statistics for pairs with nominal p-value < 1×10^-5^ only. Mappability-related artifacts may trigger false-positive trans signals as described in (Saha & Battle, 2018). We therefore excluded genes with mappability < 0.8 from analysis, filtered out variant - gene pairs where the gene cross-maps to a gene w/in 1Mb of the variant, and tested only protein-coding genes and lincRNAs.

We used permutations to determine the significance of associations. We repeatedly generated an inverse normalized phenotype and tested it against all variants, performing 20,000 such permutations and recording the strongest association per chromosome achieved in each permutation. We then used the beta-approximated p-value approach from FastQTL (Ongen et al., 2016) to compute the adjusted p-value for each gene’s strongest association, based on the most extreme permuted associations on any chromosome except the gene’s chromosome. We then applied Benjamini-Hochberg correction to these adjusted p-values to get genome-wide FDRs. trans-eGenes were those with FDR <= 5%.

For each trans-eGene, we fine-mapped trans-eQTL signals in the 2Mb region centered on the primary trans-eVariant using the tensorQTL SuSiE implementation with SuSiE L = 10.

Trans-eQTL analyses on the subset of genetically-inferred EUR ancestry samples were performed in the same manner as full-sample trans-eQTL analyses, except for the number of phenotype PCs and SuSiE L parameters (listed in Table S18). For whole blood (EUR) trans-eQTL scans, phenotype PCs 19, 22 and 27 were dropped as they associated with a likely trans-eVariant.

### trans-sQTL scans

trans-sQTL scans were performed using the same normalized splicing phenotype matrices and covariates as cis-sQTL scans.

All trans-sQTL scans tested variants with MAF >= 0.05. To reduce the probability of mapping artifacts, we removed variants with mappability < 1. To identify trans-sGenes and their respective trans-sVariants, we used tensorQTL mode --trans to test for associations between variant - splicing phenotype pairs on separate chromosomes. Due to the large number of pairs tested, we saved summary statistics for pairs with nominal p-value < 1×10^-5^ only. Mappability-related artifacts may trigger false-positive trans signals as described in (Saha & Battle, 2018). We therefore excluded genes with mappability < 0.8 from analysis, filtered out variant - gene pairs where the gene cross-maps to a gene w/in 1Mb of the variant, and tested only protein-coding genes and lincRNAs.

We used permutations to determine the significance of associations in a similar manner as was done for trans-eQTL scans, additionally adjusting for the number of splicing phenotypes tested per gene as done in (GTEx Consortium, 2020). In short, we repeatedly generated an inverse normalized phenotype and tested it against all variants, performing 20,000 such permutations and recording the strongest association per chromosome achieved in each permutation. To determine the significance of the most extreme p-value for a gene with X splicing phenotypes, we used the CDF of the first order statistic for a sample size of X from the beta-approximated CDF. We then applied Benjamini-Hochberg correction to these adjusted p-values to get genome-wide FDRs. trans-sGenes were those with FDR <= 5%.

For each trans-sGene, we fine-mapped trans-sQTL signals in the 2Mb region centered on the primary trans-sVariant using the tensorQTL SuSiE implementation with SuSiE L = 10. For each trans-sGene we ran fine mapping using only the splicing phenotype corresponding to the primary trans-sQTL, i.e. the phenotype with the strongest association.

Trans-sQTL analyses on the subset of genetically-inferred EUR ancestry samples were performed in the same manner as full-sample trans-sQTL analyses, except for the number of phenotype PCs and SuSiE L parameters (listed in Table S18).

### Clumping of primary trans signals

For analyses involving primary trans-e/sVariants (for example, counting the number of trans-eGenes per trans-eVariant), highly-linked primary trans-e/sVariants were clumped together to limit double-counting signals that likely derived from the same trans-e/sVariant but that were by chance assigned to separate variants. For each tissue and modality, we calculated the in-sample R^2^ between all trans-e/sVariants and sorted trans-e/sQTL variants according to the most significant trans-e/sQTL p-value for each trans-e/sVariant. All trans-e/sQTL variants in high LD (R^2^ >= 0.9) with the most significant trans-e/sVariant was clumped with that variant, and this procedure was iteratively applied using the remaining unclumped trans-e/sVariants until no variants remained. The trans-e/sVariant with the most significant p-value in each clump became the representative of that clump, and all trans-e/sQTL pairs involving a variant within the clump was reassigned to that representative variant.

For the tissue and modality with the greatest number of hits (whole blood trans-eQTL), this reduced the number of trans-eVariants from 810 to 614.

When fine-mapping trans-e/sQTL signals, the 2Mb fine-mapping windows were centered on the original (unclumped) trans-e/sVariants.

### Allelic fold change

Allelic fold change (Mohammadi et al., 2017) was calculated using the `calculate_afc` function in tensorQTL (v. 1.0.7) and the top PIP variants per credible set (for cis-eQTL signals) or the single variant with the strongest association (for trans-eQTL signals).

### Saturation analyses

Saturation analyses were performed using nested subsets of whole blood samples ranging in size from 500 to 6,000 samples in steps of 500. Generation of the gene expression / splicing phenotype matrix (including Leafcutter intron clustering) and the scan itself was performed in an identical manner as for the full cis-e/sQTL and trans-eQTL scans, except for cis-e/sQTL scans the fine-mapping was performed using a range of different phenotype PCs in order to show that differences between sample sizes were not due to number of PCs used. For cis-eQTL scans, fine-mapping was run with both SuSiE L = 10 and 20, and the final value of L was selected for each gene using the same criteria as in the full cis-eQTL scans; for cis-sQTL scans fine-mapping was run with SuSiE L = 10. For trans-eQTL scans, any phenotype PCs beyond 50 and any phenotype PCs that associated most strongly with variant chr3_56815721_T_C (the variant correlated with gene expression PC25 in the full whole blood cis-eQTL covariates) were dropped from the covariate matrix to avoid inadvertently adjusting out trans effects.

### Chromatin states

Hg38 chromatin states were taken from (Roadmap Epigenomics Consortium et al., 2015). We matched TOPMed tissues to corresponding Roadmap Epigenomics cell type as follows:

| TOPMed tissue | Roadmap Epigenomics cell type |
| --- | --- |
| Whole blood | Primary mononuclear cells from peripheral blood (E062) |
| PBMCs | Primary mononuclear cells from peripheral blood (E062) |
| Lung | Lung (E096) |
| Monocytes | Primary monocytes from peripheral blood (E029) |
| T cells | Primary T cells from peripheral blood (E034) |
| Nasal epithelial | A549 EtOH 0.02pct Lung Carcinoma Cell Line (E114) |

### Functional enrichments

SnpEff annotations <u>(Cingolani et al., 2012)</u> were extracted from the TOPMed freeze 9b VCF file.

Enrichments of cis-e/sQTL credible sets in annotations were determined relative to control credible sets matched on chromosome, MAF, LD, and number of genes tested against. For each variant included in the scan, we calculated the number of variants that it is in LD with (R^2^ >= 0.9; LD calculated using the TOPMed samples in the scan), the number of genes it was tested against in the scan, and the MAF. Then, for each credible set, we took the top PIP variant and selected a control variant that was tested against a similar number of genes and has approximately N LD proxies where N = (size of credible set - 1), and has a similar MAF (as similar as possible after filtering on the first two conditions). Then, the control credible set is that control variant and its LD proxies.

Clumped primary variants were used for trans-e/sQTLs enrichments. Enrichments were determined relative to control variants matched on chromosome and MAF (variants were binned into 50 equally-spaced MAF bins). Only variants included in the trans scan were considered (e.g., all control variants had mappability = 1). Each trans-e/sVariant was represented only once (e.g., a variant that was a trans-eVariant for 2 genes was not double-counted).

We used logistic regression to score enrichments. For a given annotation type (e.g., chromatin states or SnpEff annotations), we build a logistic regression model using the true signals and the control signals (credible sets + control credible sets in the case of cis-e/sQTL signals; primary hits + controls in the case of trans-e/sQTLs). The regression outcome was whether the signal was a true signal (1) or a control signal (0), and predictors were binary indicators of whether any variants in the true / control signal overlapped each of the annotations from that annotation type (e.g. for chromatin states, all the possible chromatin states were predictors in the model). Annotations overlapped by fewer than 1% of signals + control signals were dropped from the model.

### TF motif cis-eVariant overlap vs gene expression directionality analysis

Motif scans were performed using the 540 non-redundant motifs from (D’Oliveira Albanus et al., 2021), using FIMO (v. 5.5.3, with default parameters and a 0-order Markov background model generated using `fasta-get-markov`) (Grant et al., 2011). We used variant-sensitive motif scans to account for the fact that an alternative allele might create a TF binding site missing from the reference genome, or destroy a TF binding site present in the reference genome, by scanning both the reference sequence as well as replacing the reference alleles with alternative alleles.

Using the variant-sensitive motif scans, for each PWM, we counted the number of whole blood cis-eQTL signals overlapping a motif for that PWM (using the top PIP variant in each credible set from the MAF >= 0.001 cis-eQTL scan). We then calculated how often the allele that increased expression of the target gene increased the strength (FIMO score) of the motif hit. We excluded PWMs with less than 20 eQTL - motif hit overlaps. To determine whether any deviation from 0.5 was significant, we computed a p-value with a two-sided binomial test and performed Bonferroni correction across PWMs.

### trans-e/sQTL enrichment in cis-e/sQTL credible sets

Enrichment of clumped primary whole blood trans-e/sQTLs in whole blood cis-e/sQTL credible sets was calculated with a Fisher’s exact test relative to the MAF-matched trans-e/sQTL control variants used in the functional enrichments. Each trans-e/sVariant was counted only once, regardless of the number of trans-e/sGenes associated with it.

### TF gene enrichment amongst cis-eGenes for cis-eQTL overlapping trans-eQTL

This was calculated using a permutation test. First, for each cis-eGene with at least one credible set, we determined whether the gene encodes a TF based on the list of TF gene Ensembl IDs from (Lambert et al., 2018). Then, for each trans-eVariant, we determined which cis-eGenes had a cis-eQTL credible set containing the trans-eVariant, and whether any of those cis-eGenes were TF genes. We then counted the number of trans-eVariants that had at least one associated TF cis-eGene. To generate a null distribution for this statistic, we permuted the gene → is_TF relationships and re-computed the number of trans-eVariants having at least one associated TF cis-eGene, repeating this process 1000 times.

### trans-eQTL GO term enrichment

For each set of >= 10 trans-eGenes sharing a whole blood trans-eVariant (after LD clumping), we performed a GO/KEGG pathway enrichment analysis using gprofiler (Raudvere et al., 2019). As the background set of genes we used all genes tested in the whole blood trans-eQTL scan that had at least one nominal p-value < 1×10^-5^. We only tested GO:BP and KEGG terms/pathways with at least 5 genes and no more than 1000 genes. P-values were Bonferroni corrected within each trans-eVariant.

### Colocalization

Colocalizations were performed with coloc v. 5.2.1 (C. Wallace, 2021) using Bayes factor matrices from SuSiE (coloc.susie function) and default priors. A colocalization was called in the case that the posterior probability of H4 (the posterior probability that the same variant underlies the signal in both modalities) was at least 0.8. In the case that an implausible one-to-many colocalization was implied by the coloc output (e.g., a single cis-eQTL credible set colocalizing with two GWAS credible sets from the same GWAS), the colocalization with the highest posterior probability was kept.

For whole blood cis-e/sQTL - GWAS colocalizations, we used MAF >= 0.01 cis-e/sQTL for consistency across tissues and in the comparison with GTEx. Colocalization results using MAF

>= 0.001 were highly similar to results using MAF >= 0.01.

### PanUKBB GWAS fine-mapping

We used EUR and AFR LD matrices and GWAS summary statistics from PanUKBB (v. 0.3; in hg19 coordinates) for fine-mapping UKBB GWAS signals. For each ancestry we included only pass-QC phenotypes, and to avoid analyzing multiple highly correlated phenotypes we kept only phenotypes from the maximally independent set. We additionally dropped potentially sensitive phenotypes, such as those relating to mental health, sexual activity, alcohol use, and intelligence. This left 172 EUR GWAS phenotypes and 32 AFR GWAS phenotypes (Table S11). Of the 172 EUR GWAS phenotypes and 32 AFR GWAS phenotypes, 166 and 17 phenotypes had at least one genome-wide significant variant.

To determine genomic windows to fine-map, for each GWAS we identified the most genome-wide significant variant not yet in a fine-mapping window, took the 500kb window centered on that variant as a fine-mapping window, and repeated this procedure until no genome-wide significant variants (p <= 5×10^-8^) remained. We then merged overlapping fine-mapping windows such that they would be fine-mapped together. After merging, we dropped windows > 10Mb in size (LD matrices were available only up to a radius of 10Mb). We additionally dropped a small number of windows with an excessive number of variants (> 40,000) in the LD matrix.

Signals were fine-mapped using SuSiE (v. 0.11.92; susie_rss function). Low-confidence variants were excluded, as were variants that failed to lift from hg19 to hg38 or that lifted to the same position as another hg19 variant. We set the SuSiE L parameter to 10, and iteratively raised it to 20, 30, and then 40 in the case that (1) the model failed to converge at the current value of L, or (2) the number of credible sets discovered met or exceeded (L * 0.7). If more than one L was used, we calculated the maximum number of credible sets discovered across all tested values of L, and selected the final model as the one with the smallest L exceeding the maximum number of credible sets discovered. We then lifted SuSiE results to hg38, excluding any fine-mapping windows that did not lift.

### Presence of TOPMed whole blood cis-eQTL signals in GTEx whole blood / DIRECT / eQTLGen

We considered a TOPMed whole blood cis-eQTL signal to be present in GTEx whole blood if the TOPMed cis-eQTL credible set overlapped a GTEx whole blood cis-eQTL credible set.

We considered a TOPMed whole blood cis-eQTL signal to be present in eQTLGen if the TOPMed cis-eQTL credible set overlapped a eQTLGen primary eQTL or it’s LD proxies (R^2^ > 0.8 based on TOPMed whole blood samples).

We considered a TOPMed whole blood cis-eQTL signal to be present in DIRECT if the TOPMed cis-eQTL credible set overlapped a DIRECT conditional eQTL or it’s LD proxies (R^2^ > 0.8 based on TOPMed whole blood samples).

## Code availability

Source code for analyses and figures is available at: https://github.com/porchard/topmed-rnaseq-index

## Data Availability

Individual level data is available through the database of Genotypes and Phenotypes (dbGaP). All accessions are listed in acknowledgments. Cis- and trans-e/sQTL summary statistics and fine-mapping results will be available in the TOPMed Genomic Summary Results repository (dbGaP phs001974).

## Acknowledgments

Molecular data for the Trans-Omics in Precision Medicine (TOPMed) program was supported by the National Heart, Lung and Blood Institute (NHLBI). RNASeq for “NHLBI TOPMed: Whole Genome Sequencing and Related Phenotypes in the Framingham Heart Study” (phs000974)” was performed at the Northwest Genomics Center (HHSN268201600032I). Genome Sequencing for “NHLBI TOPMed: Whole Genome Sequencing and Related Phenotypes in the Framingham Heart Study” (phs000974)” was performed at Broad Genomics (HHSN268201600034I, 3U54HG003067-12S2, 3R01HL092577-06S1).

RNASeq for “NHLBI TOPMed: Genetic Epidemiology of COPD (COPDGene) (phs000951)” was performed at the Northwest Genomics Center (HHSN268201600032I). Genome Sequencing for “NHLBI TOPMed: Genetic Epidemiology of COPD (COPDGene) (phs000951)” was performed at Broad Genomics (HHSN268201500014C) and the Northwest Genomics Center (3R01HL089856-08S1).

RNASeq for “NHLBI TOPMed - NHGRI CCDG: Genes-Environments and Admixture in Latino Asthmatics (GALA II) (phs000920)” was performed at Broad Genomics (HHSN268201600034I). Genome Sequencing for ““NHLBI TOPMed - NHGRI CCDG: Genes-Environments and Admixture in Latino Asthmatics (GALA II) (phs000920)” was performed at NYGC Genomics (3R01HL117004-02S3).

RNASeq for “NHLBI TOPMed: Study of African Americans, Asthma, Genes and Environment (SAGE) (phs000921)” was performed at Broad Genomics (HHSN268201600034I). Genome Sequencing for “NHLBI TOPMed: Study of African Americans, Asthma, Genes and Environment (SAGE) (phs000921)” was performed at NYGC Genomics (3R01HL117004-02S3) and the Northwest Genomics Center (HHSN268201600032I).

RNASeq for “NHLBI TOPMed: SubPopulations and InteRmediate Outcome Measures In COPD Study (SPIROMICS) (phs001927)” was performed at Northwest Genomics Center (HHSN268201600032I). Genome Sequencing for “NHLBI TOPMed: SubPopulations and InteRmediate Outcome Measures In COPD Study (SPIROMICS) (phs001927)” was performed at Broad Genomics (HHSN268201600034I).

RNASeq for “NHLBI TOPMed: MESA and MESA Family AA-CAC (phs001416)” was performed at Northwest Genomics Center (HHSN268201600032I) and Broad Genomics (HHSN268201600034I). Genome Sequencing for “NHLBI TOPMed: MESA and MESA Family AA-CAC (phs001416)” was performed at Broad Genomics (3U54HG003067-13S1, HHSN268201600034I, HHSN268201500014C).

RNASeq for “NHLBI TOPMed: Women’s Health Initiative (WHI) (phs001237)” was performed at Broad Genomics (HHSN268201600034I). Genome Sequencing for “NHLBI TOPMed: Women’s Health Initiative (WHI) (phs001237)” was performed at Broad Genomics (HHSN268201500014C).

RNASeq for “NHLBI TOPMed: Lung Tissue Research Consortium (LTRC) (phs001662)” was performed at Northwest Genomics Center (HHSN268201600032I). Genome Sequencing for “NHLBI TOPMed: Lung Tissue Research Consortium (LTRC) (phs001662)” was performed at Broad Genomics (HHSN268201600034I).

Core support including centralized genomic read mapping and genotype calling, along with variant quality metrics and filtering were provided by the TOPMed Informatics Research Center (3R01HL-117626-02S1; contract HHSN268201800002I). Core support including phenotype harmonization, data management, sample-identity QC, and general program coordination were provided by the TOPMed Data Coordinating Center (R01HL-120393; U01HL-120393; contract HHSN268201800001I). We gratefully acknowledge the studies and participants who provided biological samples and data for TOPMed.

The Framingham Heart Study (FHS) acknowledges the support of contracts NO1-HC-25195, HHSN268201500001I and 75N92019D00031 from the National Heart, Lung and Blood Institute and grant supplement R01 HL092577-06S1 for this research. We also acknowledge the dedication of the FHS study participants without whom this research would not be possible. Dr. Vasan is supported in part by the Evans Medical Foundation and the Jay and Louis Coffman Endowment from the Department of Medicine, Boston University School of Medicine.

The WHI program is funded by the National Heart, Lung, and Blood Institute, National Institutes of Health, U.S. Department of Health and Human Services through contracts 75N92021D00001, 75N92021D00002, 75N92021D00003, 75N92021D00004, 75N92021D00005.

The COPDGene project described was supported by grants from the National Heart, Lung, and Blood Institute (U01 HL089897 and U01 HL089856), and by NIH contract 75N92023D00011. The content is solely the responsibility of the authors and does not necessarily represent the official views of the National Heart, Lung, and Blood Institute or the National Institutes of Health. The COPDGene project is also supported by the COPD Foundation through contributions made to an Industry Advisory Committee that has included AstraZeneca, Bayer Pharmaceuticals, Boehringer Ingelheim, Genentech, GlaxoSmithKline, Novartis, Pfizer, and Sunovion. A full listing of COPDGene investigators can be found at: http://www.copdgene.org/directory.

This study utilized biological specimens and data provided by the Lung Tissue Research Consortium (LTRC) supported by the National Heart, Lung, and Blood Institute (NHLBI). The LTRC TOPMed project was also supported by P01 HL114501 and R01 HL133135.

Whole genome sequencing (WGS) for the Trans-Omics in Precision Medicine (TOPMed) program was supported by the National Heart, Lung and Blood Institute (NHLBI). WGS for “NHLBI TOPMed: Multi-Ethnic Study of Atherosclerosis (MESA)” (phs001416.v3.p1) was performed at the Broad Institute of MIT and Harvard (3U54HG003067-13S1). Centralized read mapping and genotype calling, along with variant quality metrics and filtering were provided by the TOPMed Informatics Research Center (3R01HL-117626-02S1). Phenotype harmonization, data management, sample-identity QC, and general study coordination, were provided by the TOPMed Data Coordinating Center (3R01HL-120393-02S1), and TOPMed MESA Multi-Omics (HHSN2682015000031/HSN26800004). The MESA projects are conducted and supported by the National Heart, Lung, and Blood Institute (NHLBI) in collaboration with MESA investigators. Support for the Multi-Ethnic Study of Atherosclerosis (MESA) projects are conducted and supported by the National Heart, Lung, and Blood Institute (NHLBI) in collaboration with MESA investigators. Support for MESA is provided by contracts 75N92020D00001, HHSN268201500003I, N01-HC-95159, 75N92020D00005, N01-HC-95160, 75N92020D00002, N01-HC-95161, 75N92020D00003, N01-HC-95162, 75N92020D00006, N01-HC-95163, 75N92020D00004, N01-HC-95164, 75N92020D00007, N01-HC-95165, N01-HC-95166, N01-HC-95167, N01-HC-95168, N01-HC-95169, UL1-TR-000040, UL1-TR-001079, UL1-TR-001420, UL1TR001881, DK063491, HL148610, and R01HL105756. The authors thank the other investigators, the staff, and the participants of the MESA study for their valuable contributions. A full list of participating MESA investigators and institutes can be found at http://www.mesa-nhlbi.org.

The Genes-environments and Admixture in Latino Asthmatics (GALA II) Study was supported by the National Heart, Lung, and Blood Institute of the National Institute of Health (NIH) grants R01HL117004 and X01HL134589; study enrollment supported by the Sandler Family Foundation, the American Asthma Foundation, the RWJF Amos Medical Faculty Development Program, Harry Wm. and Diana V. Hind Distinguished Professor in Pharmaceutical Sciences II and the National Institute of Environmental Health Sciences grant R01ES015794. The GALA II study collaborators include Shannon Thyne, UCSF; Harold J. Farber, Texas Children’s Hospital; Denise Serebrisky, Jacobi Medical Center; Rajesh Kumar, Lurie Children’s Hospital of Chicago; Emerita Brigino-Buenaventura, Kaiser Permanente; Michael A. LeNoir, Bay Area Pediatrics; Kelley Meade, UCSF Benioff Children’s Hospital, Oakland; William Rodriguez-Cintron, VA Hospital, Puerto Rico; Pedro C. Avila, Northwestern University; Jose R. Rodriguez-Santana, Centro de Neumologia Pediatrica; Luisa N. Borrell, City University of New York; Adam Davis, UCSF Benioff Children’s Hospital, Oakland; Saunak Sen, University of Tennessee and Fred Lurmann, Sonoma Technologies, Inc. The authors acknowledge the families and patients for their participation and thank the numerous health care providers and community clinics for their support and participation in GALA II. In particular, the authors thank study coordinator Sandra Salazar; the recruiters who obtained the data: Duanny Alva, MD, Gaby Ayala-Rodriguez, Lisa Caine, Elizabeth Castellanos, Jaime Colon, Denise DeJesus, Blanca Lopez, Brenda Lopez, MD, Louis Martos, Vivian Medina, Juana Olivo, Mario Peralta, Esther Pomares, MD, Jihan Quraishi, Johanna Rodriguez, Shahdad Saeedi, Dean Soto, Ana Taveras; and the lab researcher Celeste Eng who processed the biospecimens.

The Study of African Americans, Asthma, Genes and Environments (SAGE) was supported by by the National Heart, Lung, and Blood Institute of the National Institute of Health (NIH) grants R01HL117004 and X01HL134589; study enrollment supported by the Sandler Family Foundation, the American Asthma Foundation, the RWJF Amos Medical Faculty Development Program, Harry Wm. and Diana V. Hind Distinguished Professor in Pharmaceutical Sciences II. The SAGE study collaborators include Harold J. Farber, Texas Children’s Hospital; Emerita Brigino-Buenaventura, Kaiser Permanente; Michael A. LeNoir, Bay Area Pediatrics; Kelley Meade, UCSF Benioff Children’s Hospital, Oakland; Luisa N. Borrell, City University of New York; Adam Davis, UCSF Benioff Children’s Hospital, Oakland and Fred Lurmann, Sonoma Technologies, Inc. The authors acknowledge the families and patients for their participation and thank the numerous health care providers and community clinics for their support and participation in SAGE. In particular, the authors thank study coordinator Sandra Salazar; the recruiters who obtained the data: Lisa Caine, Elizabeth Castellanos, Brenda Lopez, MD, Shahdad Saeedi; and the lab researcher Celeste Eng who processed the biospecimens.

The authors thank the SPIROMICS participants and participating physicians, investigators, study coordinators, and staff for making this research possible. More information about the study and how to access SPIROMICS data is available at www.spiromics.org. The authors would like to acknowledge the University of North Carolina at Chapel Hill BioSpecimen Processing Facility (http://bsp.web.unc.edu/) and Alexis Lab (https://www.med.unc.edu/cemalb/facultyresearch/alexislab/) for sample processing, storage, and sample disbursements. The University of North Carolina BioSpecimen Processing Facility (RRID: SCR_021290 ; https://bsp.web.unc.edu) was supported in part by NCI Cancer Center Support Grant 5P30CA016086-46 and the NIEHS UNC Center for Environmental Health and Susceptibility Center grant 5P30ES010126.

We would like to acknowledge the following current and former investigators of the SPIROMICS sites and reading centers: Neil E Alexis, MD; Wayne H Anderson, PhD; Mehrdad Arjomandi, MD; Igor Barjaktarevic, MD, PhD; R Graham Barr, MD, DrPH; Patricia Basta, PhD; Lori A Bateman, MS; Christina Bellinger, MD; Surya P Bhatt, MD; Eugene R Bleecker, MD; Richard C Boucher, MD; Russell P Bowler, MD, PhD; Russell G Buhr, MD, PhD; Stephanie A Christenson, MD; Alejandro P Comellas, MD; Christopher B Cooper, MD, PhD; David J Couper, PhD; Gerard J Criner, MD; Ronald G Crystal, MD; Jeffrey L Curtis, MD; Claire M Doerschuk, MD; Mark T Dransfield, MD; M Bradley Drummond, MD; Christine M Freeman, PhD; Craig Galban, PhD; Katherine Gershner, DO; MeiLan K Han, MD, MS; Nadia N Hansel, MD, MPH; Annette T Hastie, PhD; Eric A Hoffman, PhD; Yvonne J Huang, MD; Robert J Kaner, MD; Richard E Kanner, MD; Mehmet Kesimer, PhD; Eric C Kleerup, MD; Jerry A Krishnan, MD, PhD; Wassim W Labaki, MD; Lisa M LaVange, PhD; Stephen C Lazarus, MD; Fernando J Martinez, MD, MS; Merry-Lynn McDonald, PhD; Deborah A Meyers, PhD; Wendy C Moore, MD; John D Newell Jr, MD; Elizabeth C Oelsner, MD, MPH; Jill Ohar, MD; Wanda K O’Neal, PhD; Victor E Ortega, MD, PhD; Robert Paine, III, MD; Laura Paulin, MD, MHS; Stephen P Peters, MD, PhD; Cheryl Pirozzi, MD; Nirupama Putcha, MD, MHS; Sanjeev Raman, MBBS, MD; Stephen I Rennard, MD; Donald P Tashkin, MD; J Michael Wells, MD; Robert A Wise, MD; and Prescott G Woodruff, MD, MPH. The project officers from the Lung Division of the National Heart, Lung, and Blood Institute were Lisa Postow, PhD, and Lisa Viviano, BSN; SPIROMICS was supported by contracts from the NIH/NHLBI (HHSN268200900013C, HHSN268200900014C, HHSN268200900015C, HHSN268200900016C, HHSN268200900017C, HHSN268200900018C, HHSN268200900019C, HHSN268200900020C), grants from the NIH/NHLBI (U01 HL137880, U24 HL141762, R01 HL182622, and R01 HL144718), and supplemented by contributions made through the Foundation for the NIH and the COPD Foundation from Amgen; AstraZeneca/MedImmune; Bayer; Bellerophon Therapeutics; Boehringer-Ingelheim Pharmaceuticals, Inc.; Chiesi Farmaceutici S.p.A.; Forest Research Institute, Inc.; Genentech; GlaxoSmithKline; Grifols Therapeutics, Inc.; Ikaria, Inc.; MGC

Diagnostics; Novartis Pharmaceuticals Corporation; Nycomed GmbH; Polarean; ProterixBio; Regeneron Pharmaceuticals, Inc.; Sanofi; Sunovion; Takeda Pharmaceutical Company; and Theravance Biopharma and Mylan/Viatris.

LK was supported by funding from National Cancer Institute (R00CA246076). LMR is funded by R01AG075884.

