## Supplementary figures for "Cross-cohort analysis of expression and splicing quantitative trait loci in TOPMed"

**A.**

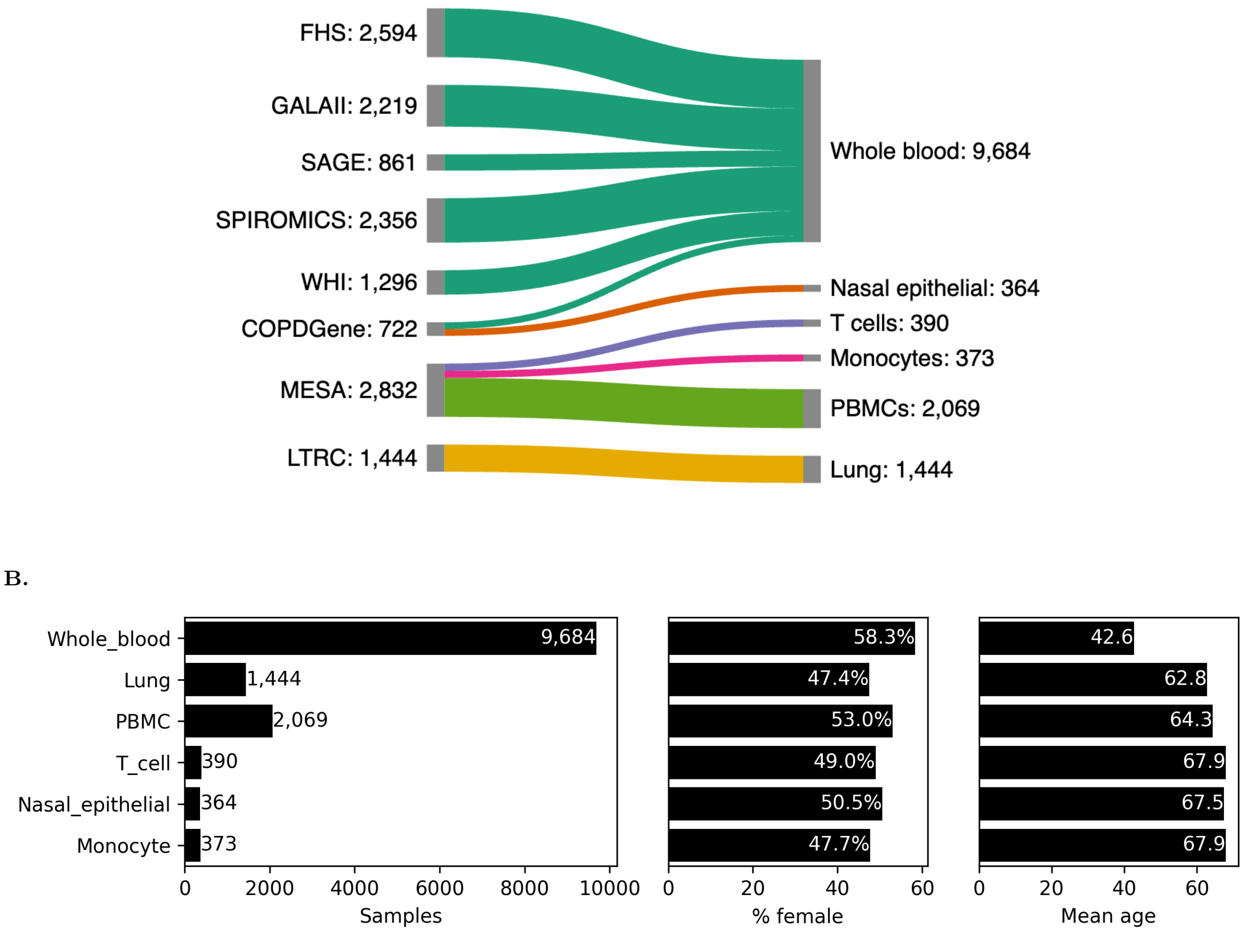


**Figure S1.** RNA-seq sample sizes and donor demographics. Donors are double-counted if contributing $> 1$ sample per tissue. The full TOPMed study names and accompanying abbreviations are: Framingham Heart Study (FHS); Gene-Environments and Admixture in Latino Asthmatics (GALA II); Study of African Americans, Asthma, Genes, & Environments (SAGE); Subpopulations and Intermediate Outcome Measures In COPD Study (SPIROMICS); Women's Health Initiative (WHI); COPDGene Study (COPDGene); Multi-Ethnic Study of Atherosclerosis (MESA); Lung Tissue Research Consortium (LTRC)

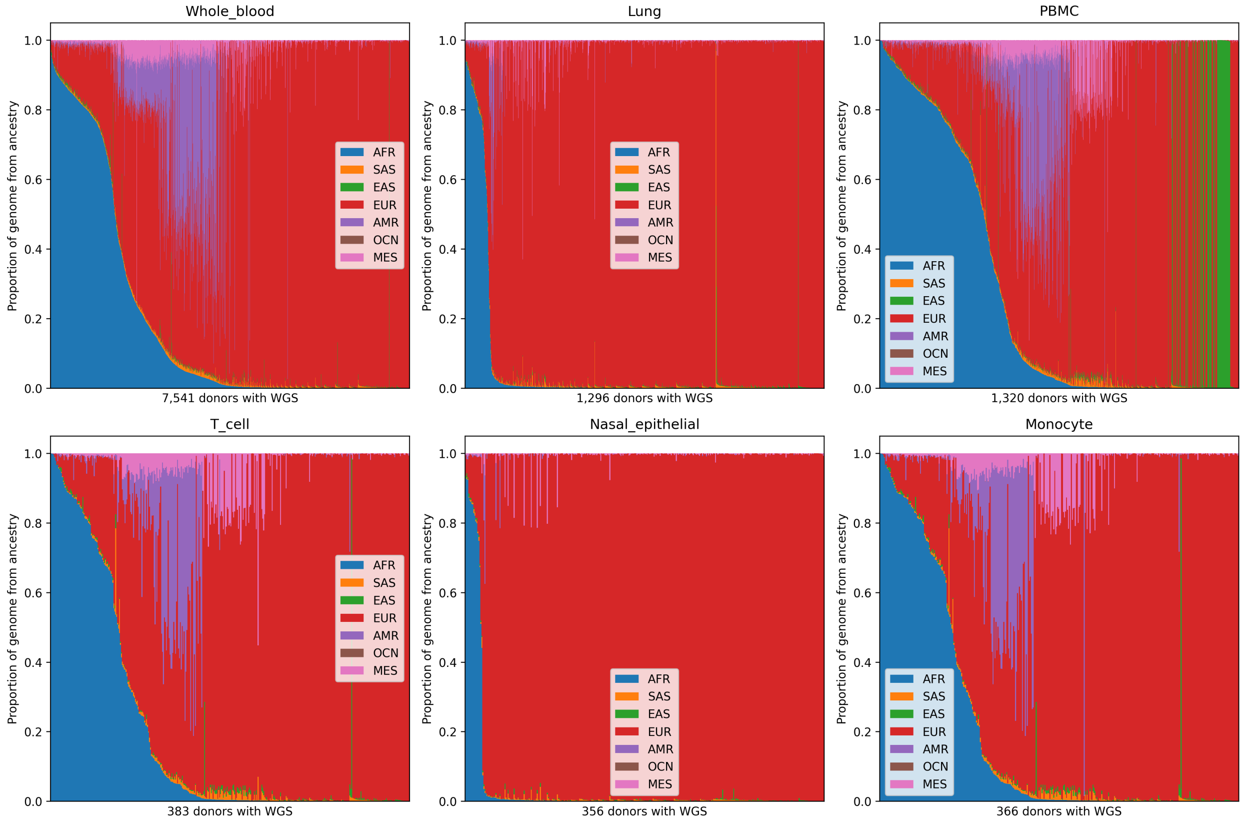


**Figure S2.** RNA-seq donor genetic ancestry as estimated from WGS data. Abbreviations: EUR (Europe), AFR (Sub-saharan Africa), AMR (Native America), EAS (East Asia), MES (Middle East), SAS (Central and South Asia).

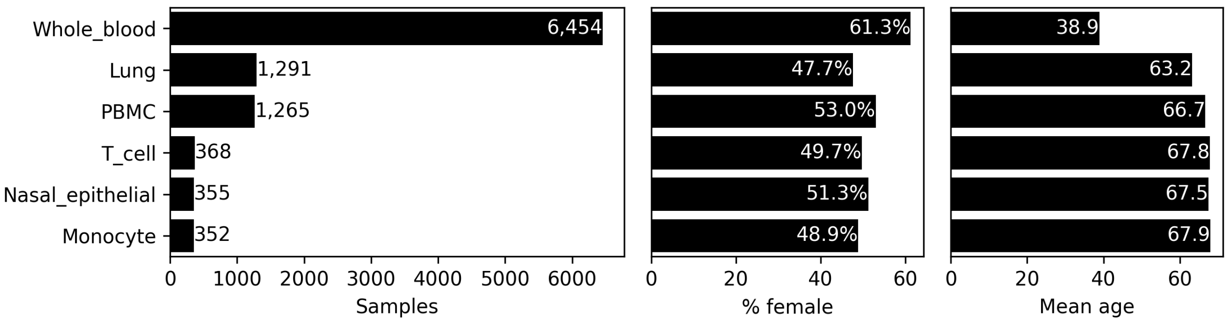


**Figure S3.** Sample sizes and donor demographics for QTL scans.

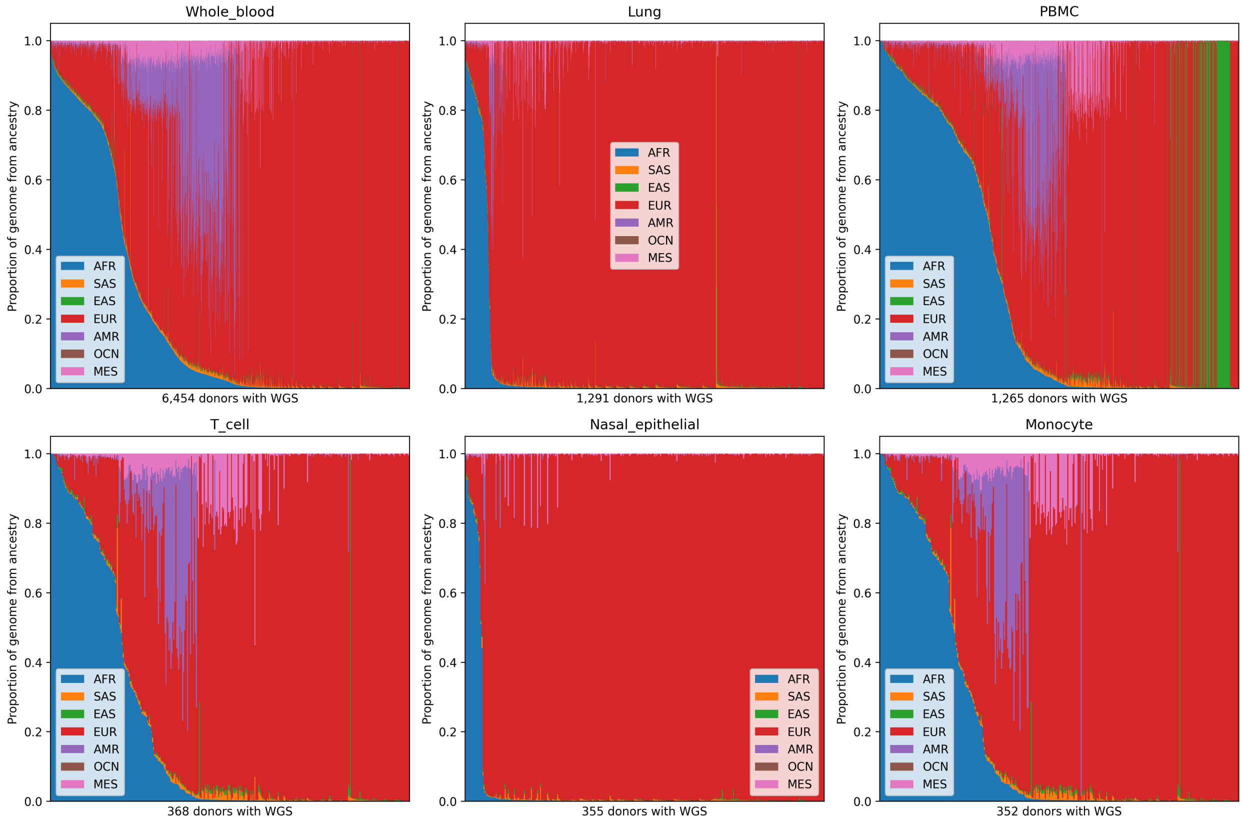


**Figure S4.** Genetic ancestry for RNA-seq samples used in QTL scans. Abbreviations: EUR (Europe), AFR (Sub-saharan Africa) AMR (Native America), EAS (East Asia), MES (Middle East), SAS (Central and South Asia).

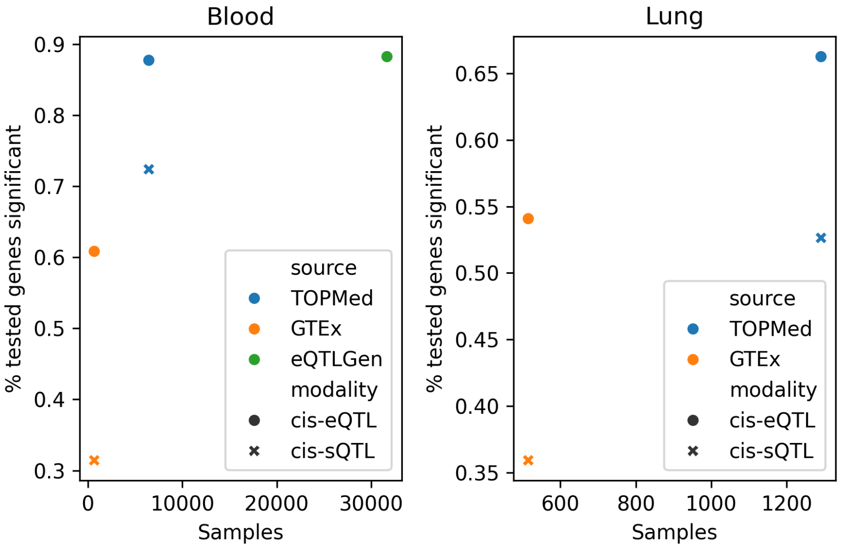


**Figure S5.** Rate of blood and lung cis-e/sGene discovery in TOPMed, GTEx, and eQTLGen. Blood is whole blood in TOPMed and GTEx, and a mix of whole blood and PBMCs in eQTLGen. eQTLGen analyzed only blood eQTLs.

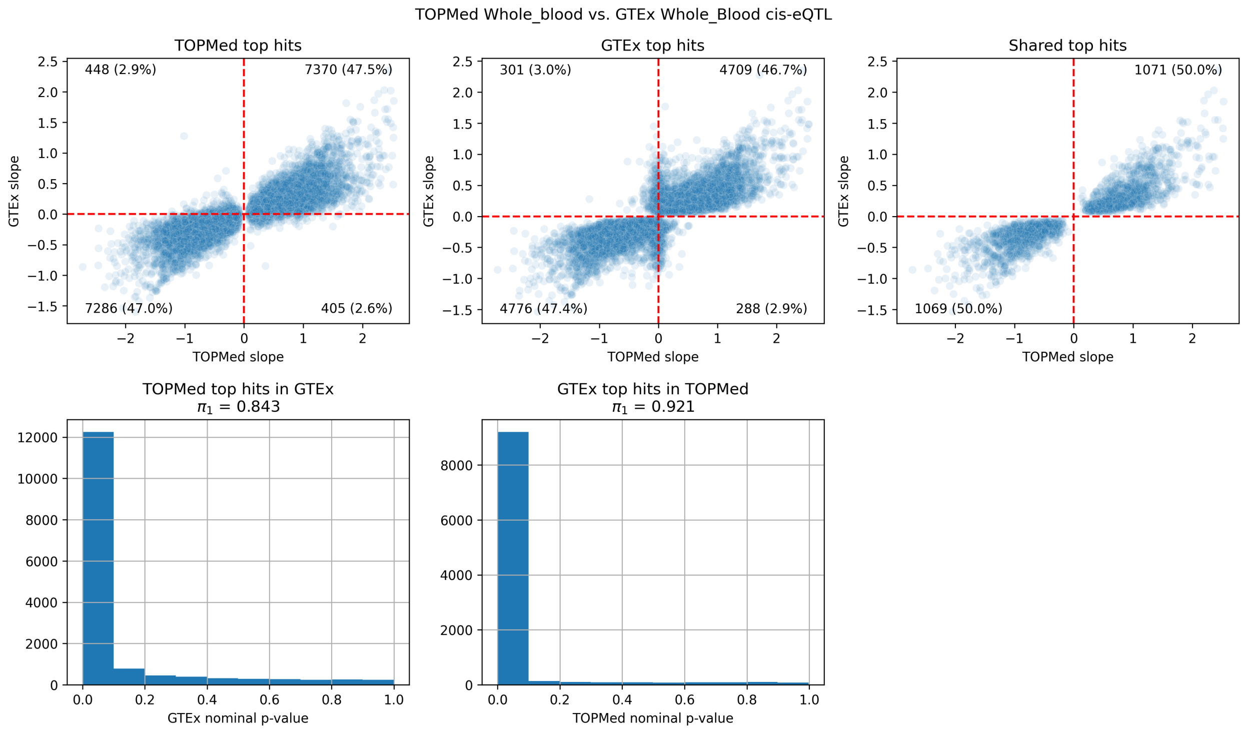


**Figure S6.** TOPMed vs GTEx primary cis-eQTL signals, whole blood. Slope magnitude is not directly comparable due to differences in normalization. $\pi_1$ represents the estimated proportion of true non-null p-values, calculated using the qvalue package in R.

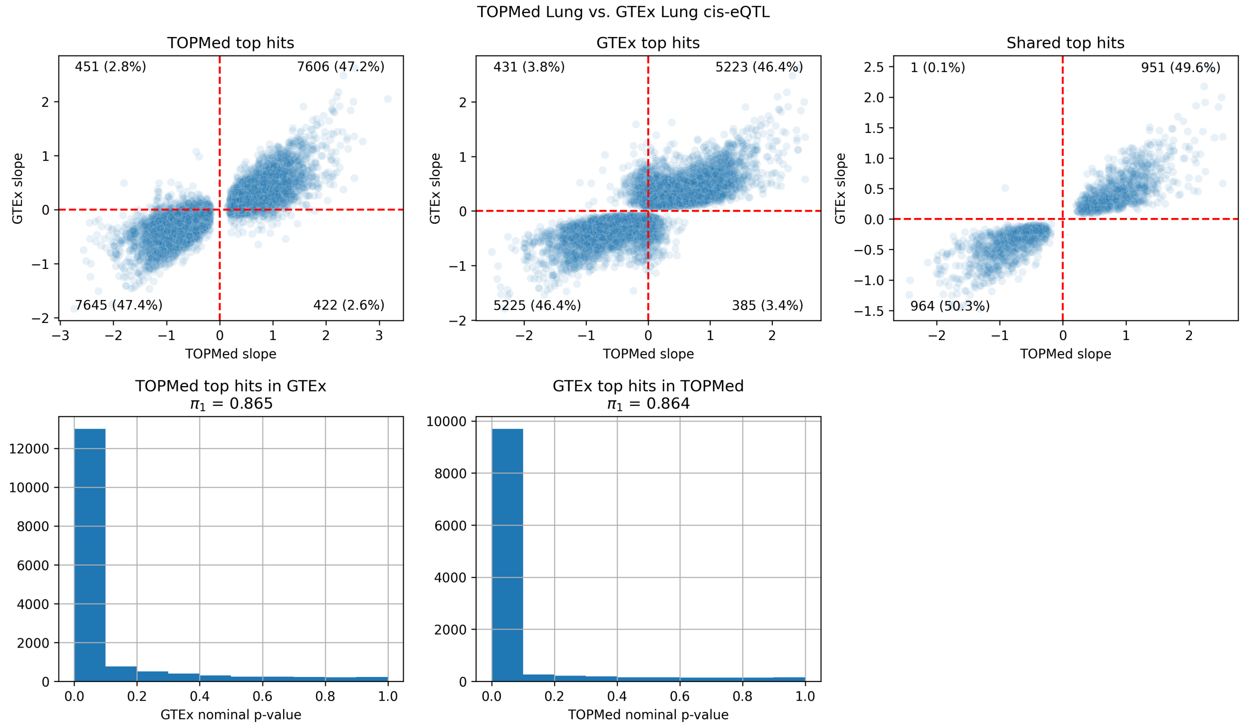


**Figure S7.** TOPMed vs GTEx primary cis-eQTL signals, lung.

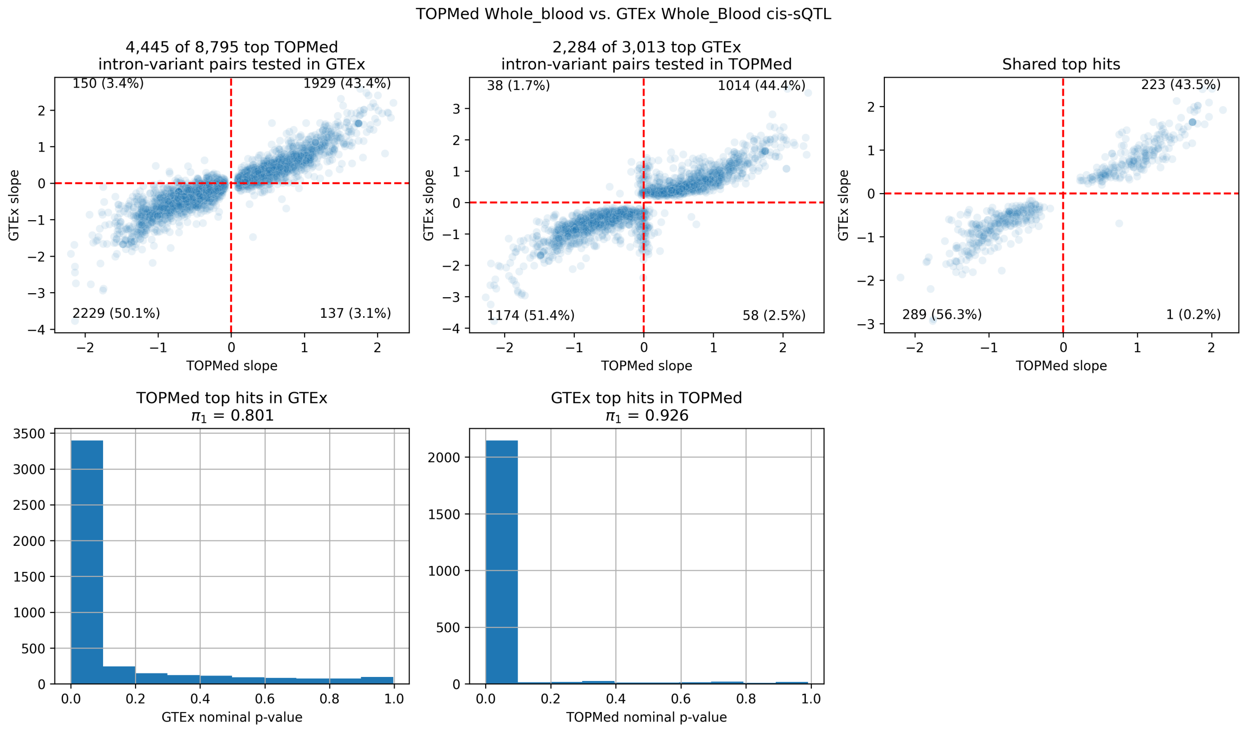


**Figure S8.** TOPMed vs GTEx primary cis-sQTL signals, whole blood.

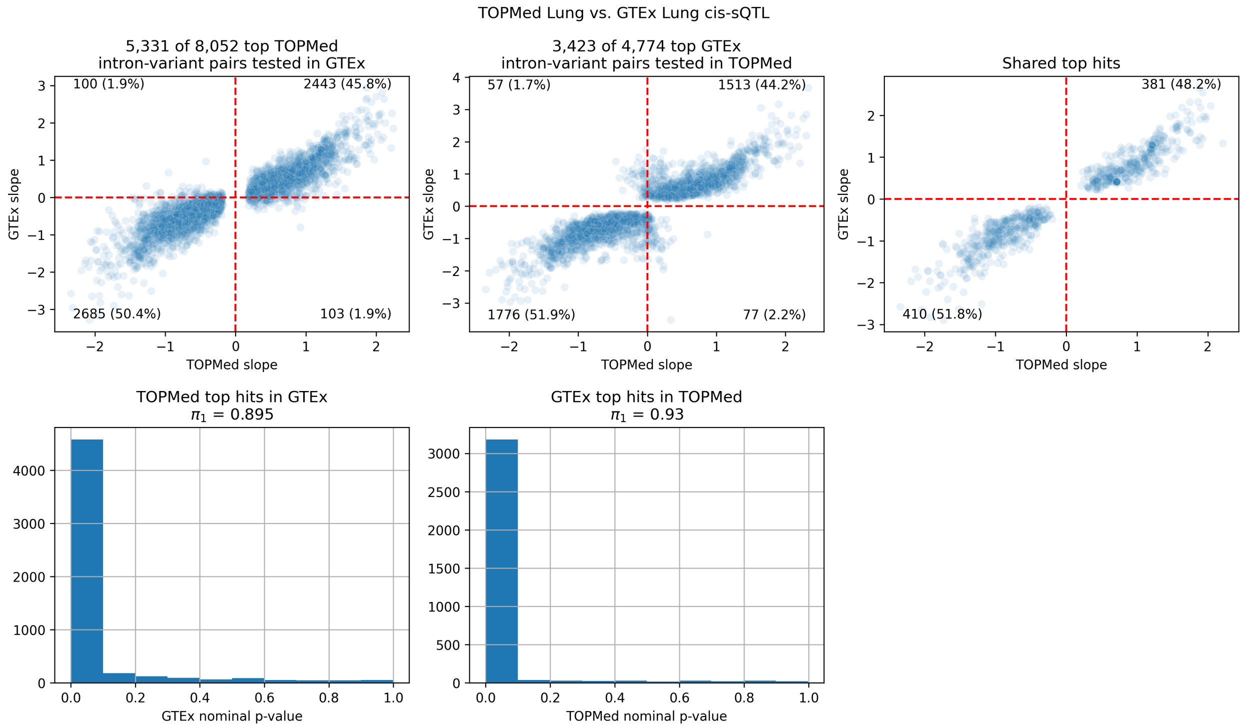


**Figure S9.** TOPMed vs GTEx primary cis-sQTL signals, lung.

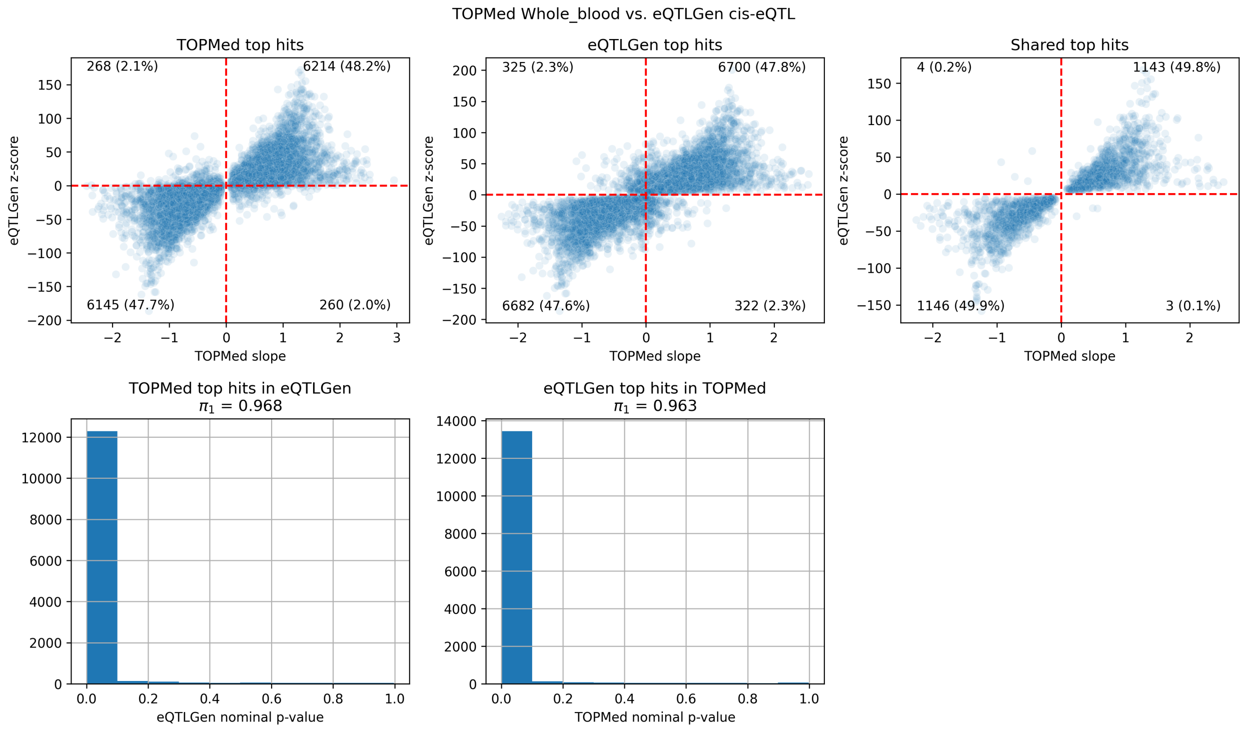


**Figure S10.** TOPMed vs eQTLGen primary cis-eQTL signals, whole blood.

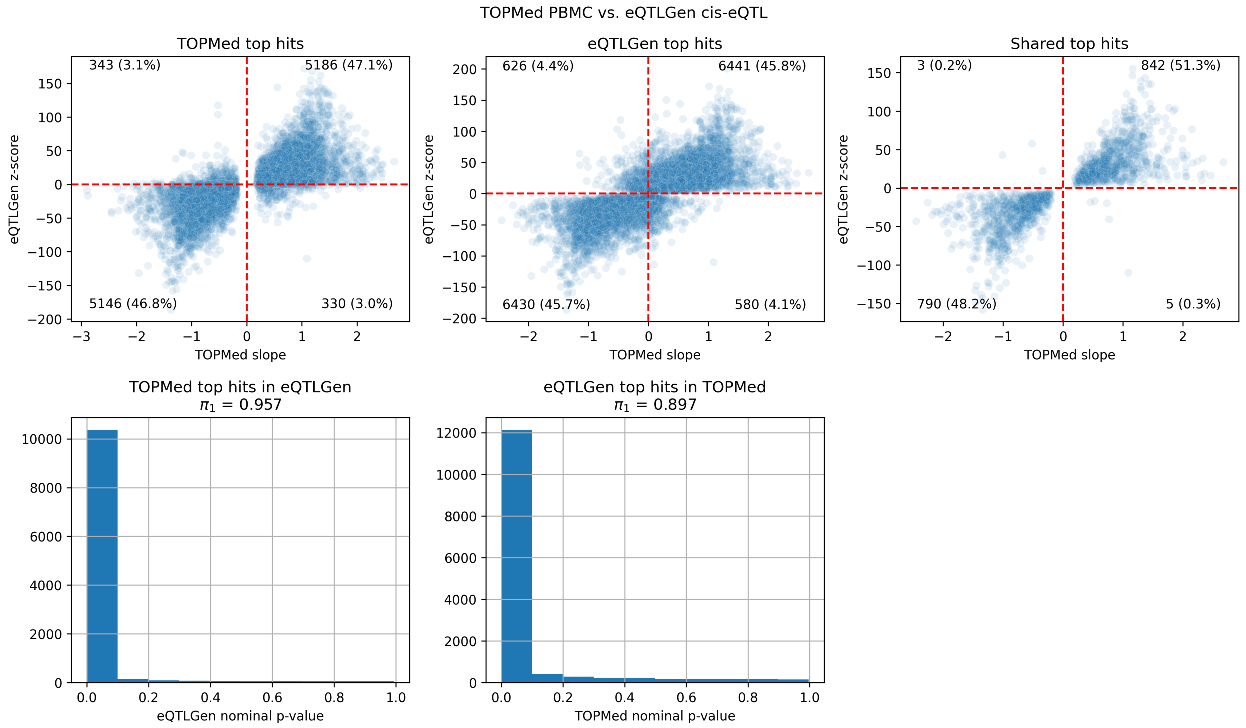


**Figure S11.** TOPMed vs eQTLGen primary cis-eQTL signals, PBMCs.

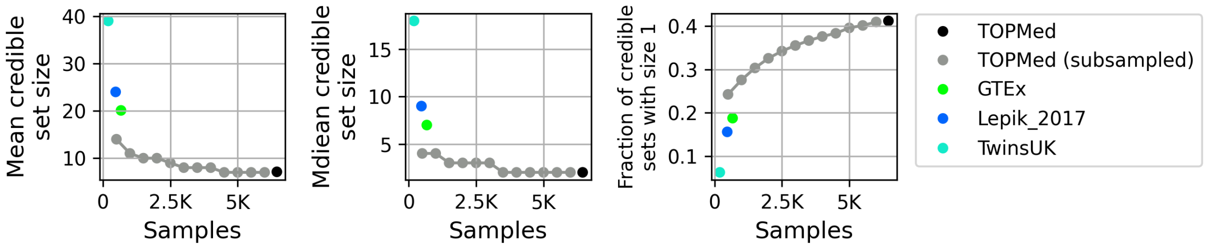


**Figure S12.** Cis-eQTL credible set size for TOPMed (including nested sample subsets), GTEx, TwinsUK, and Lepik_2017 whole blood datasets. TwinsUK and Lepik_2017 results from eQTL-Catalogue. Results are shown for 1% FDR cis-Genes, as eQTL-Catalogue fine-maps QTL signals for 1% FDR cis-e/sGenes.

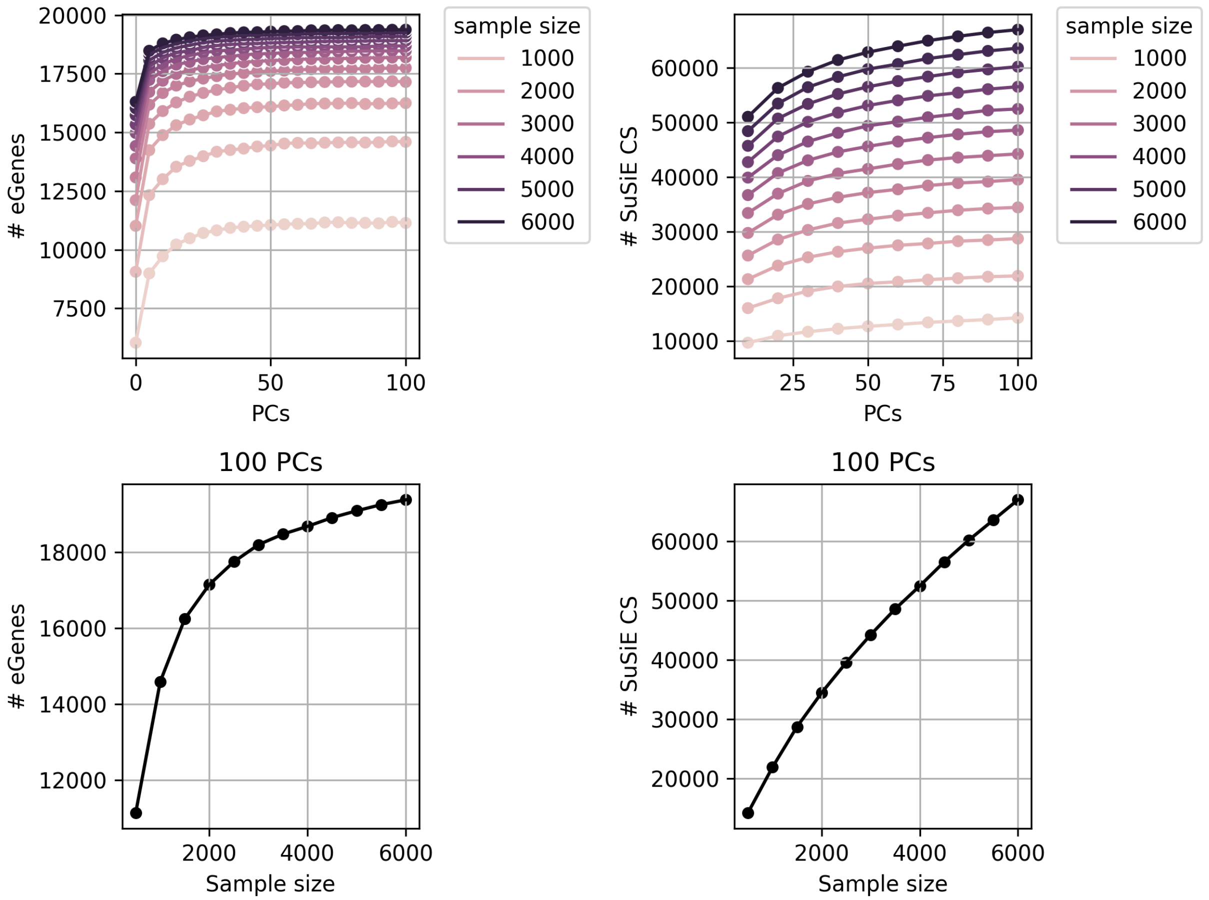


**Figure S13.** Cis-eQTL saturation analysis using nested subsets of whole blood samples (500 - 6,000 samples in steps of 500). Top left: cis-eGene discovery, 5% FDR. Top right: cis-eQTL 95% credible sets from SuSiE fine-mapping. PCs represents the number of gene expression PCs used as covariates in the eQTL scan (generally speaking, as sample size increases the optimal number of PCs used will also increase). To ease comparison across sample sizes, the bottom row displays cis-eGene discovery (left) and cis-eQTL credible set discovery (right) when using 100 gene expression PCs for all samples sizes.

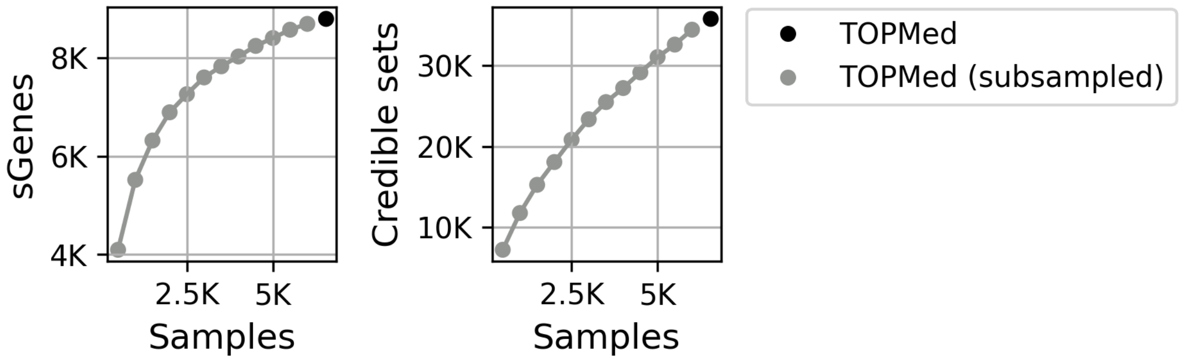


**Figure S14.** Cis-sQTL saturation analysis using nested subsets of whole blood samples (500 - 6,000 samples in steps of 500). Left: cis-sGene discovery, 5% FDR. Right: cis-sQTL 95% credible sets from SuSiE fine-mapping. 10 splicing phenotype PCs were used as covariates for all scans.

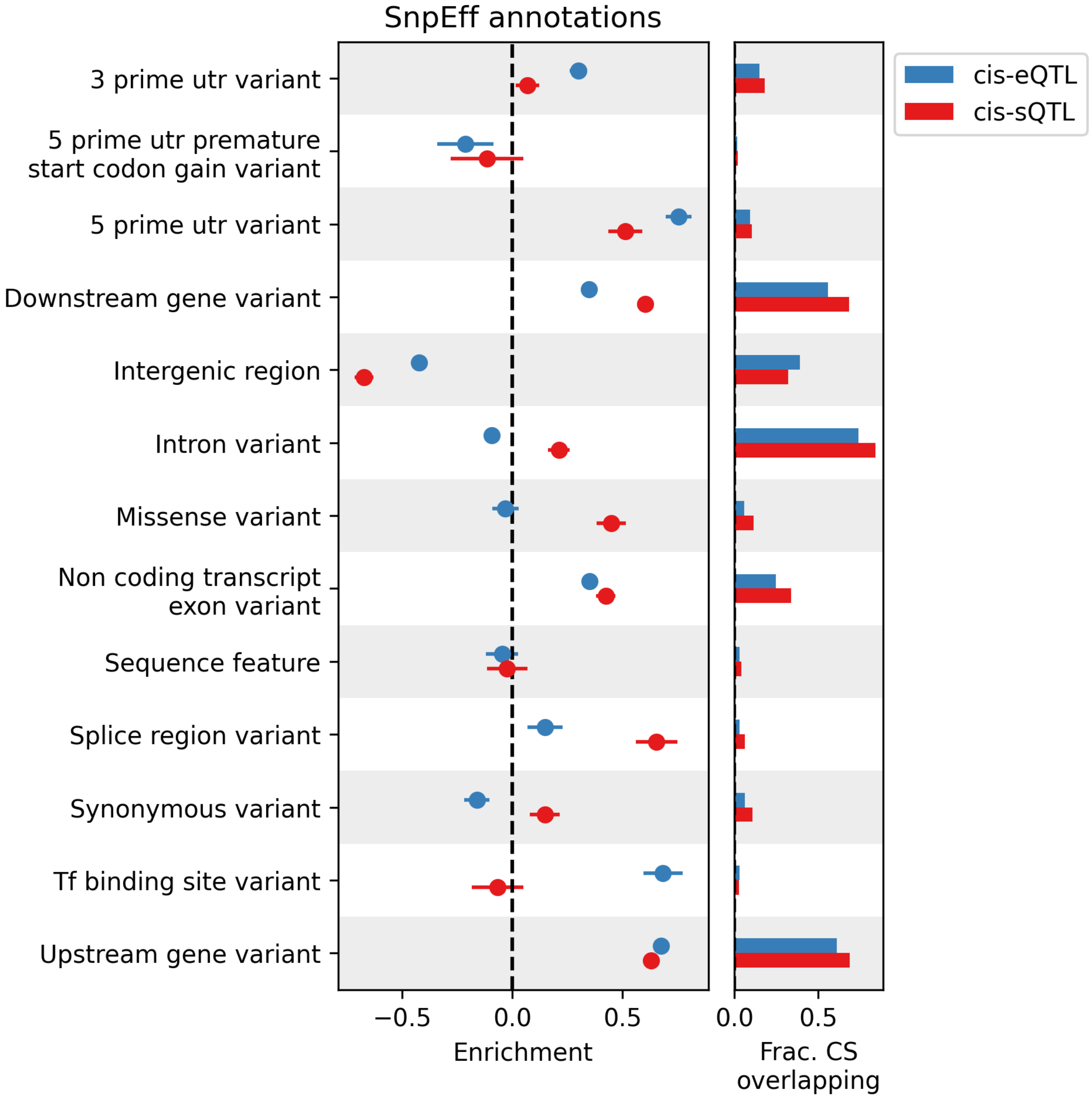


**Figure S15.** Functional annotation enrichments for whole blood cis-e/sQTLs in SnpEff annotations. Enrichment calculated relative to control credible sets matched on MAF, LD, and number of genes tested against; error bars represent 95% confidence intervals.

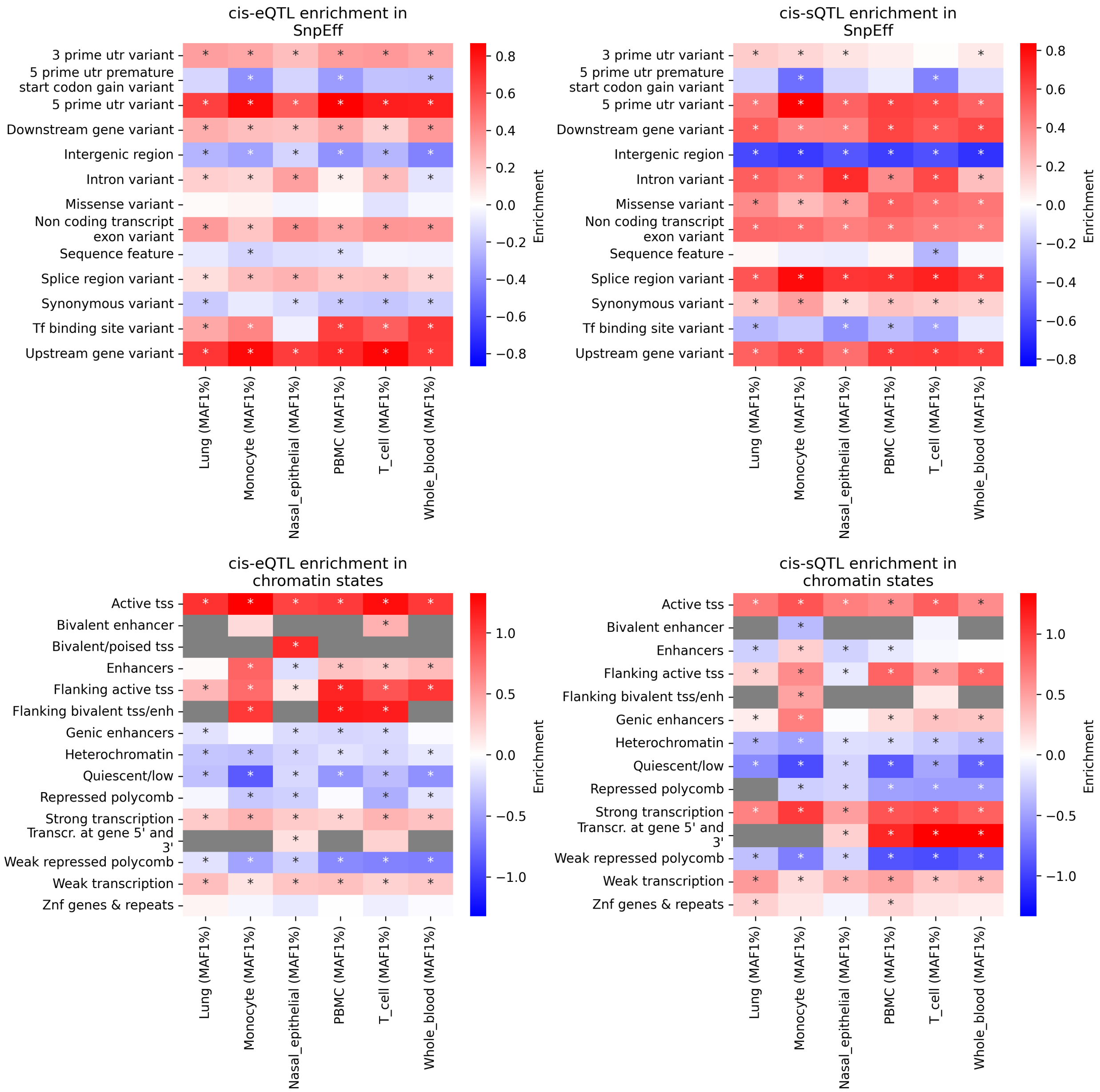


**Figure S16.** Enrichment of cis-e/sQTL credible sets in functional annotations (SnpEff annotations or Roadmap Epigenomics chromatin states). Enrichment calculated relative to control credible sets matched on chromosome, MAF, LD, and number of genes tested against, using logistic regression (see Methods; enrichment = logistic regression coefficient; asterisk denotes nominal p-value $< 0.05$).

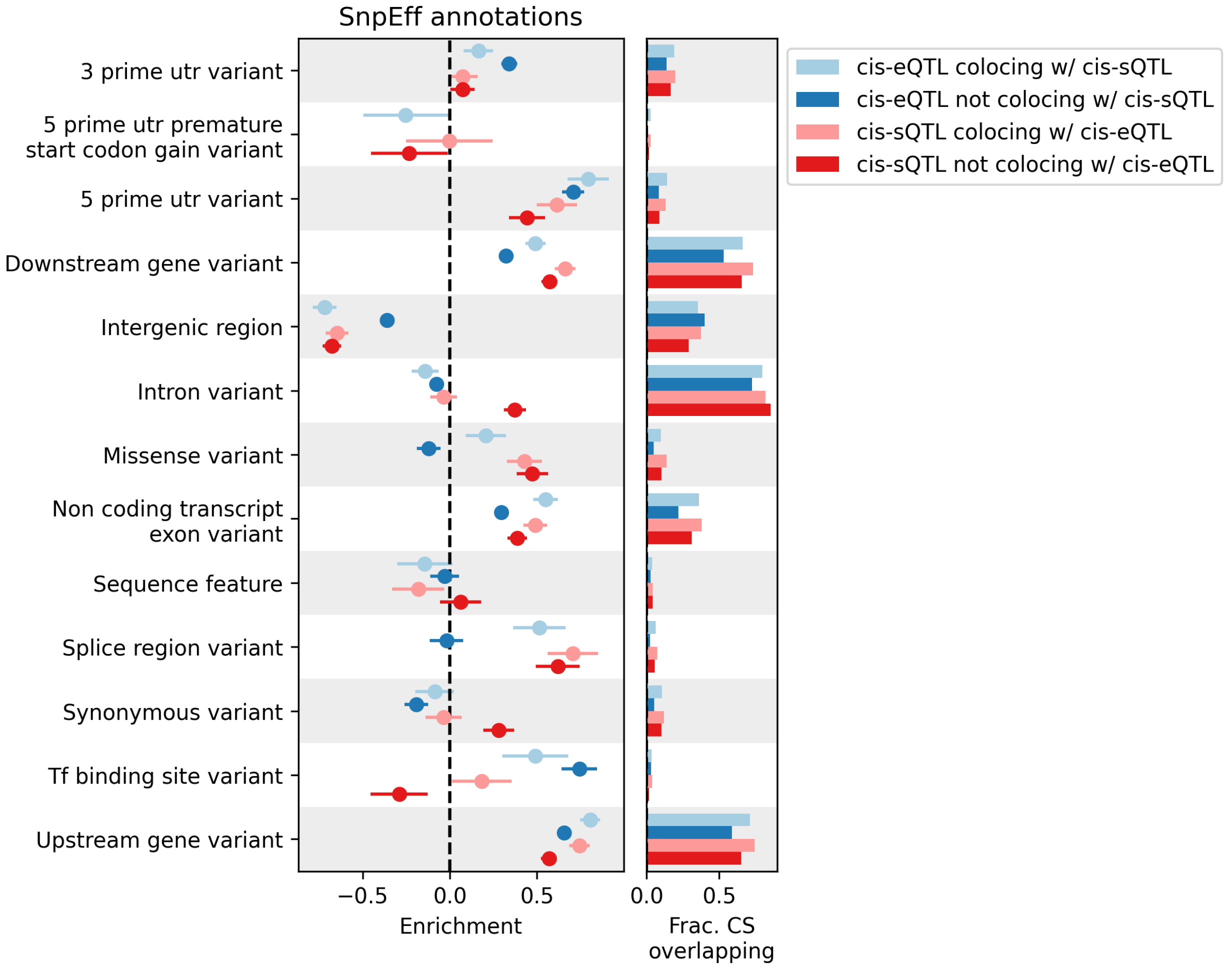


**Figure S17.** Enrichment of whole blood cis-eQTLs that are/are not also whole blood cis-sQTLs, and whole blood cis-sQTLs that are/are not also whole blood cis-eQTLs, in SnpEff annotations. Enrichment calculated relative to control credible sets matched on chromosome, MAF, LD, and number of genes tested against, using logistic regression (see Methods; enrichment = logistic regression coefficient; error bars = 95% confidence intervals).

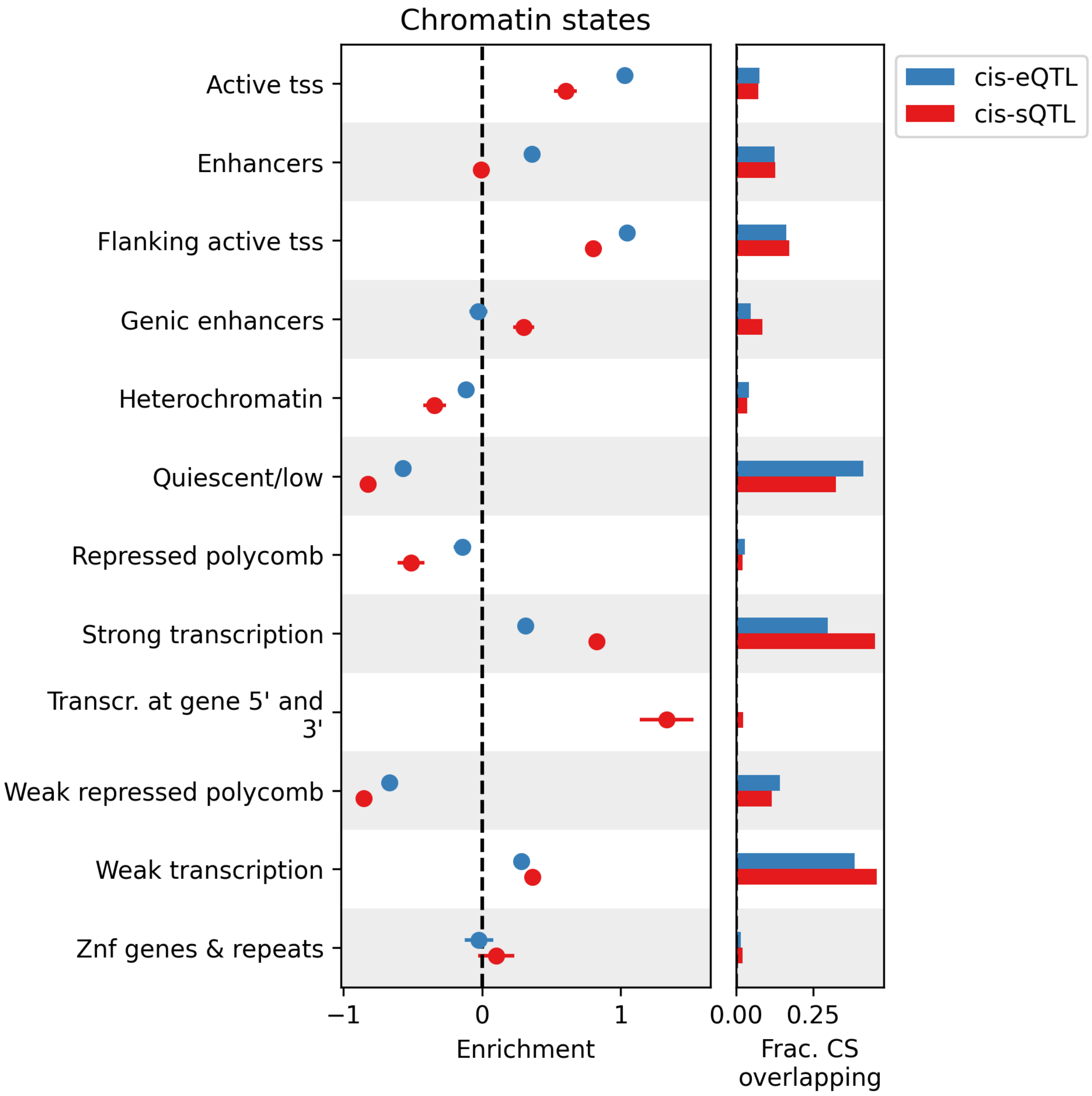


**Figure S18.** Functional annotation enrichments for whole blood cis-e/sQTLs in Roadmap Epigenomics chromatin states. Enrichment calculated relative to control credible sets matched on MAF, LD, and number of genes tested against; error bars represent 95% confidence intervals.

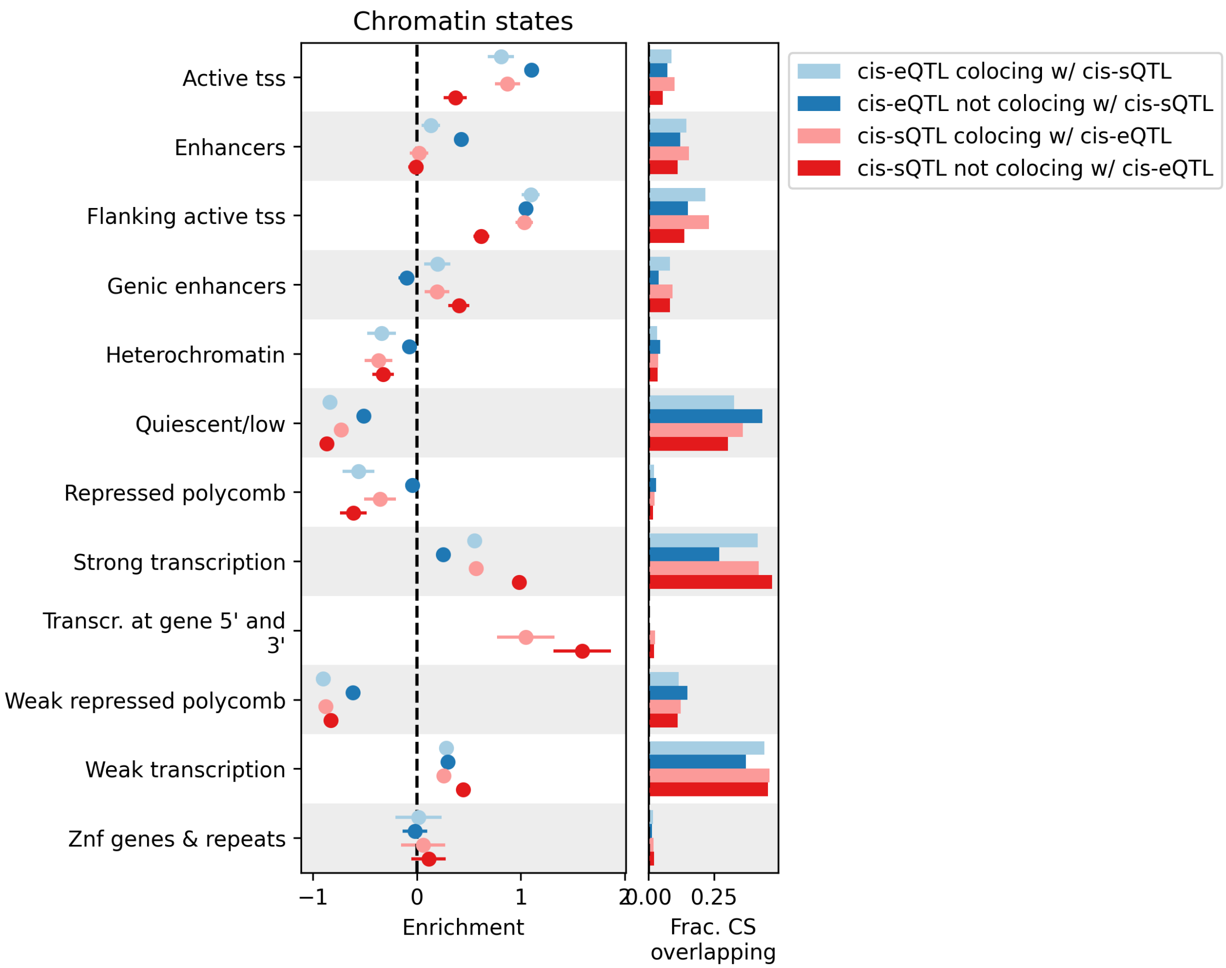


**Figure S19.** Enrichment of whole blood cis-eQTLs that are/are not also whole blood cis-sQTLs, and whole blood cis-sQTLs that are/are not also whole blood cis-eQTLs, in Roadmap Epigenomics chromatin states. Error bars represent 95% confidence intervals.

**A.**

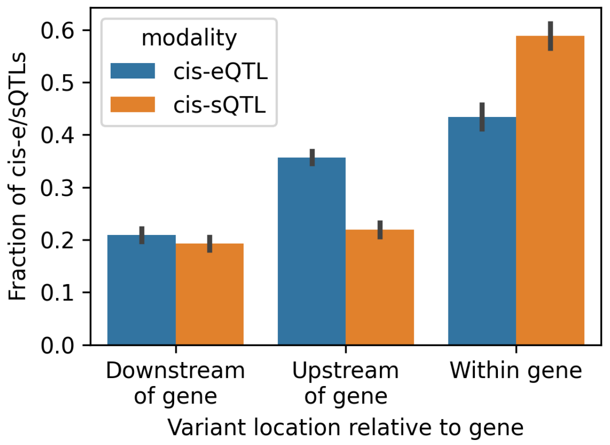


**B.**

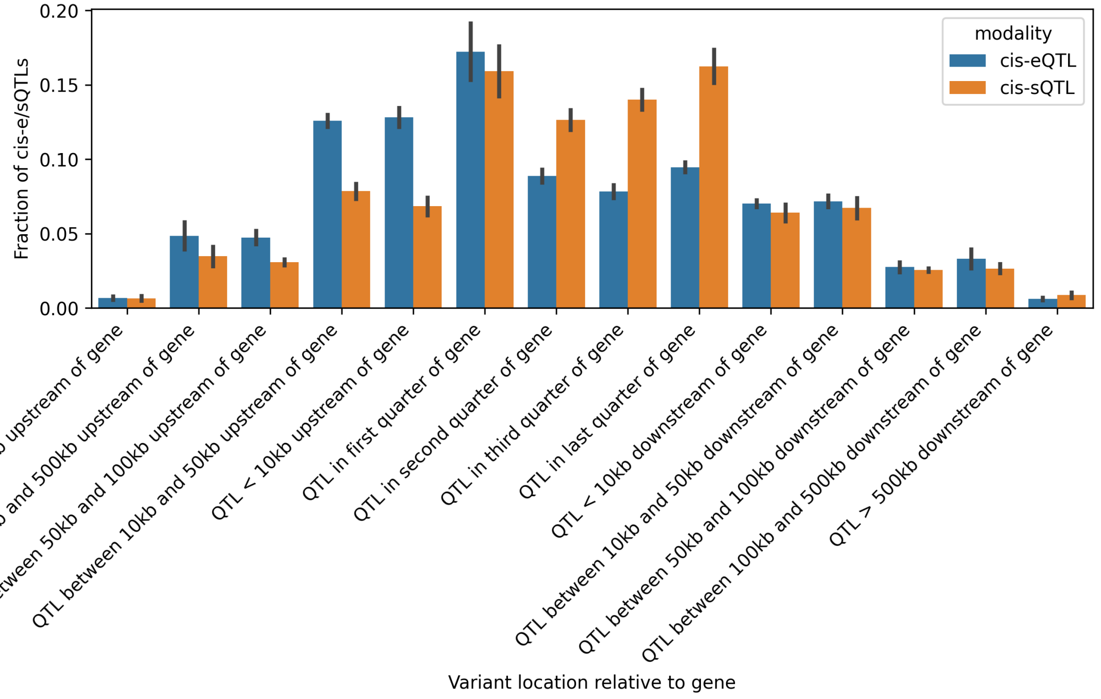


**Figure S20.** Cis-e/sQTL location relative to gene body. Each QTL signal is represented by the top PIP variant in the credible set. Error bars represent standard deviation (over tissues).

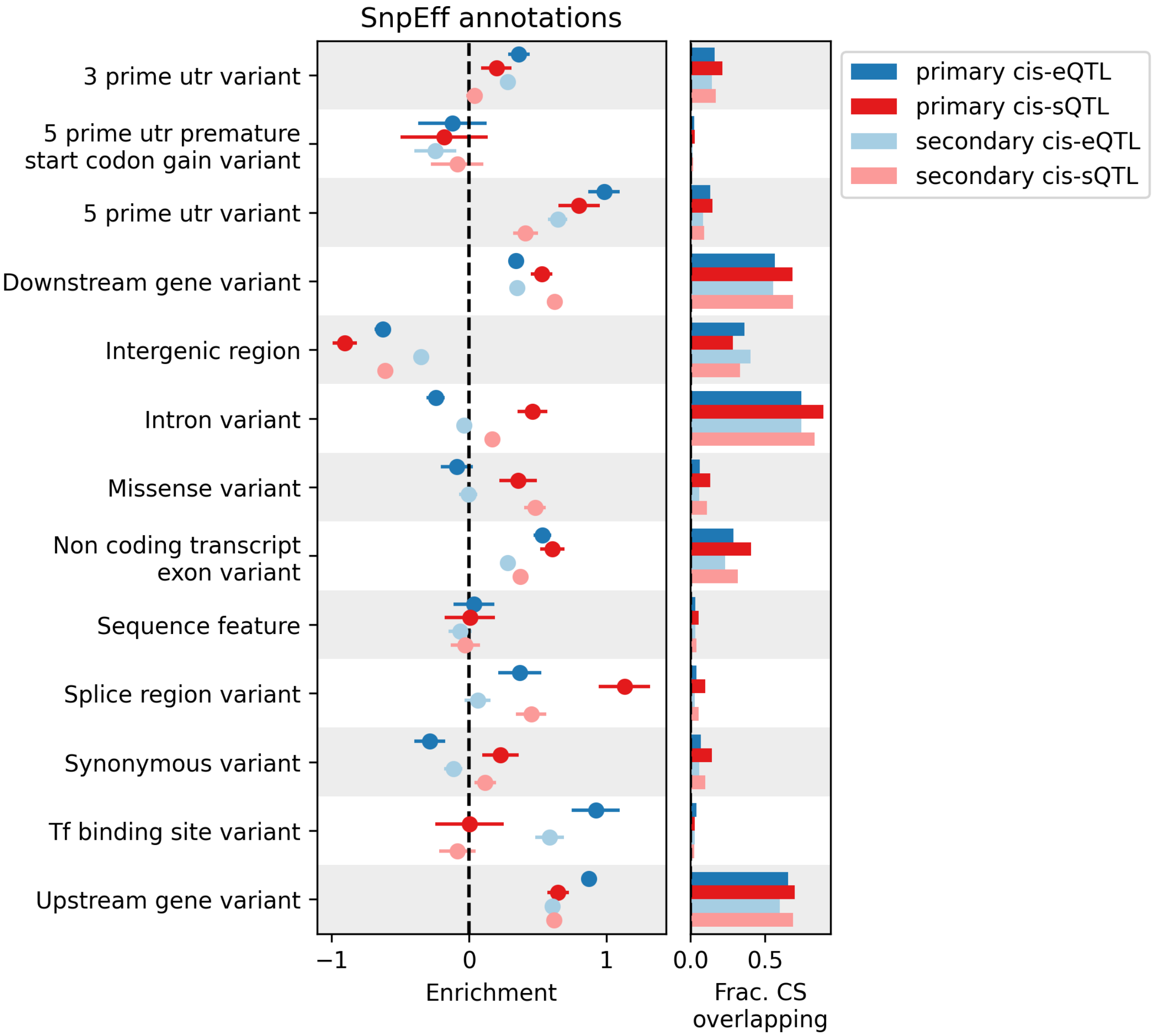


**Figure S21.** Enrichment of whole blood primary and secondary cis-e/sQTLs, in SnpEff annotations. Error bars represent 95% confidence intervals.

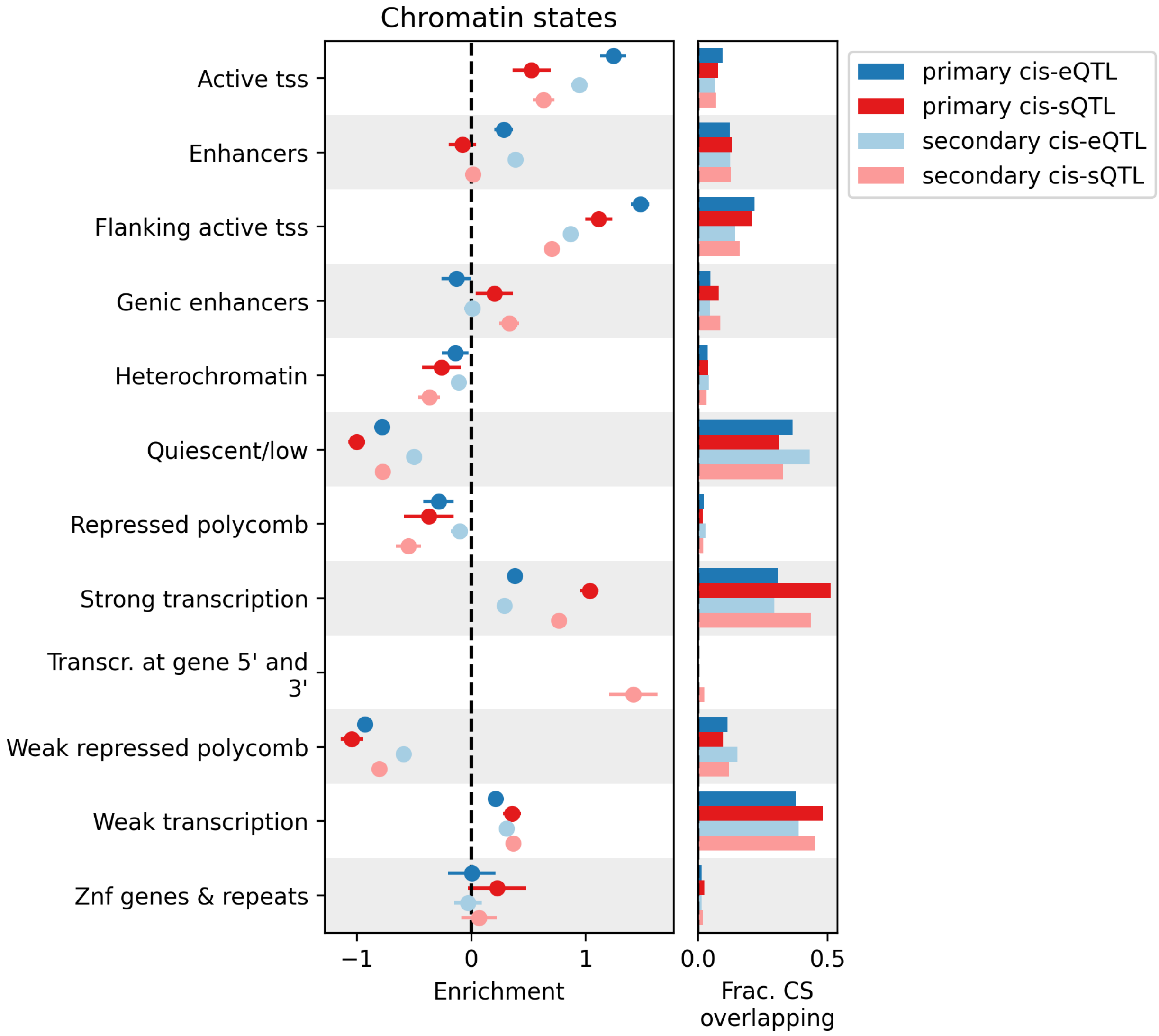


**Figure S22.** Enrichment of whole blood primary and secondary cis-e/sQTLs, in Roadmap Epigenomics chromatin states. Error bars represent 95% confidence intervals.

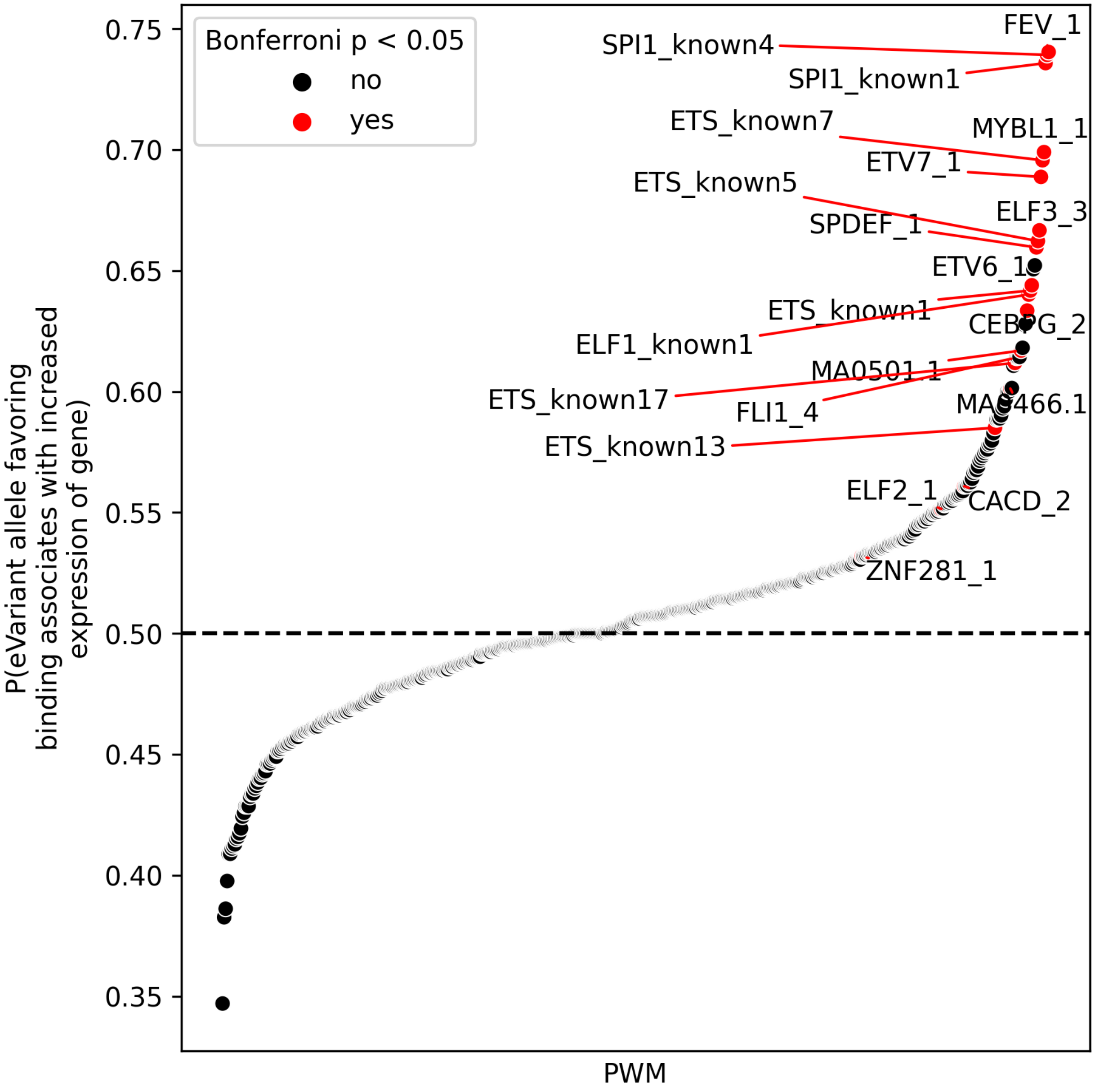


**Figure S23.** For eVariants overlapping TF motifs, does the allele favoring TF binding correlate with increased or decreased gene expression? TF motif scans were performed in an allele-sensitive manner (see Methods). P-values determined via two-sided binominal test. Results reflect top PIP variants in whole blood cis-eQTL scan

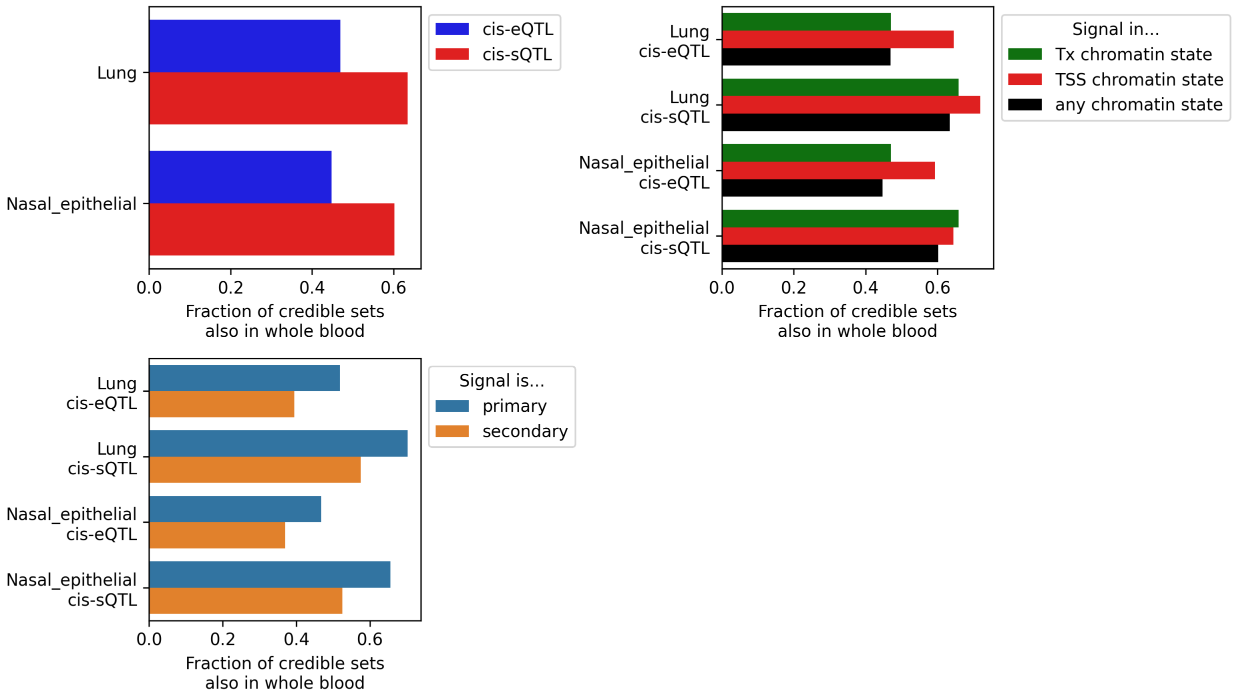


**Figure S24.** Fraction of lung and nasal epithelial cis-e/sQTL also identified in whole blood (top left), and subdivided by lung / nasal epithelial chromatin state (top right) or whether the signal was a primary or a secondary signal (bottom left). TSS state = ”Active TSS”, ”Flanking Active TSS”, or ”Bivalent/Poised TSS states”; Tx state = ”Transcr. at gene 5' and 3'”, ”Strong transcription”, and ”Weak transcription” states

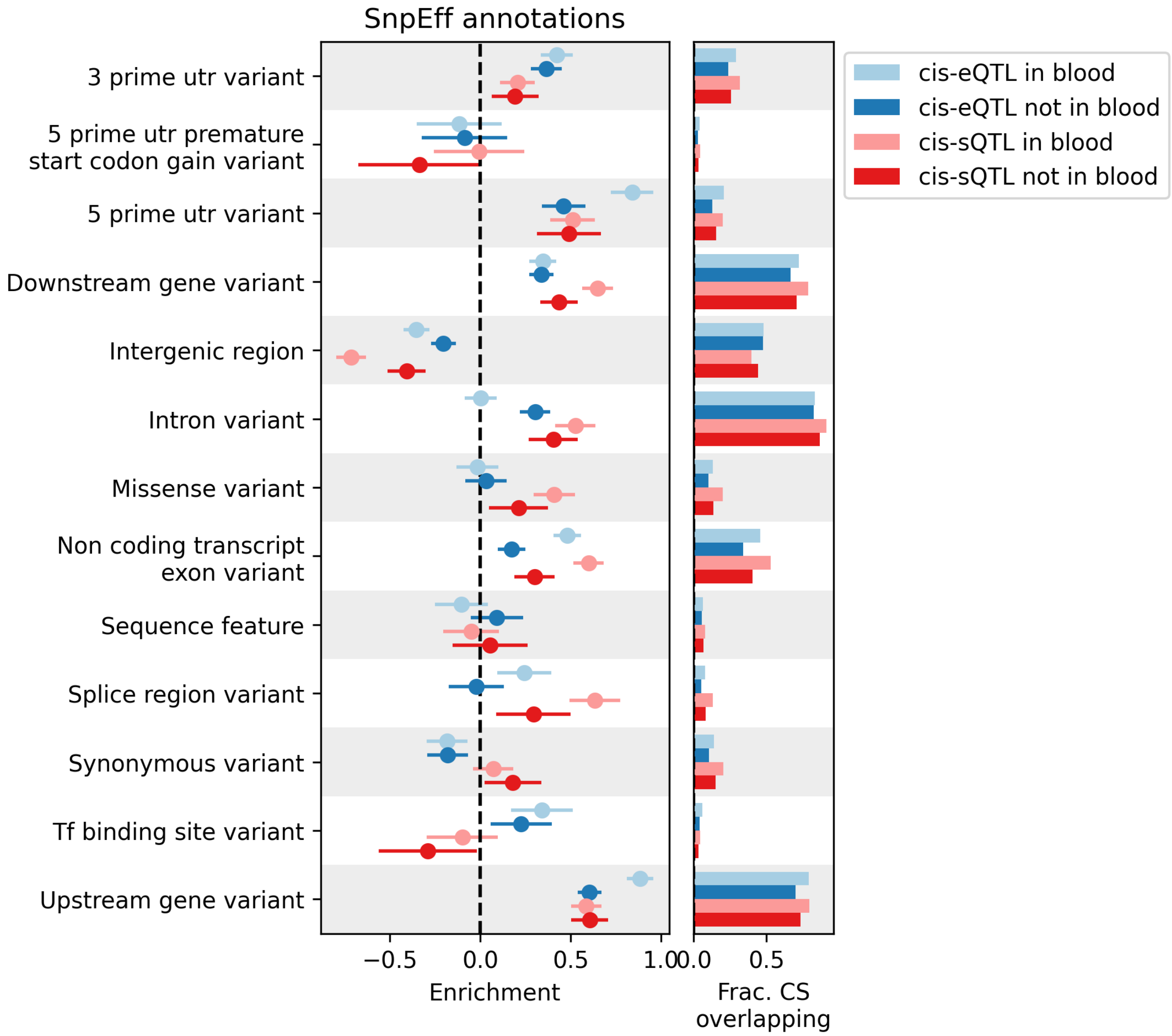


**Figure S25.** Enrichment of lung cis-e/sQTLs that are/are not shared with whole blood, in SnpEff annotations. Error bars represent 95% confidence intervals.

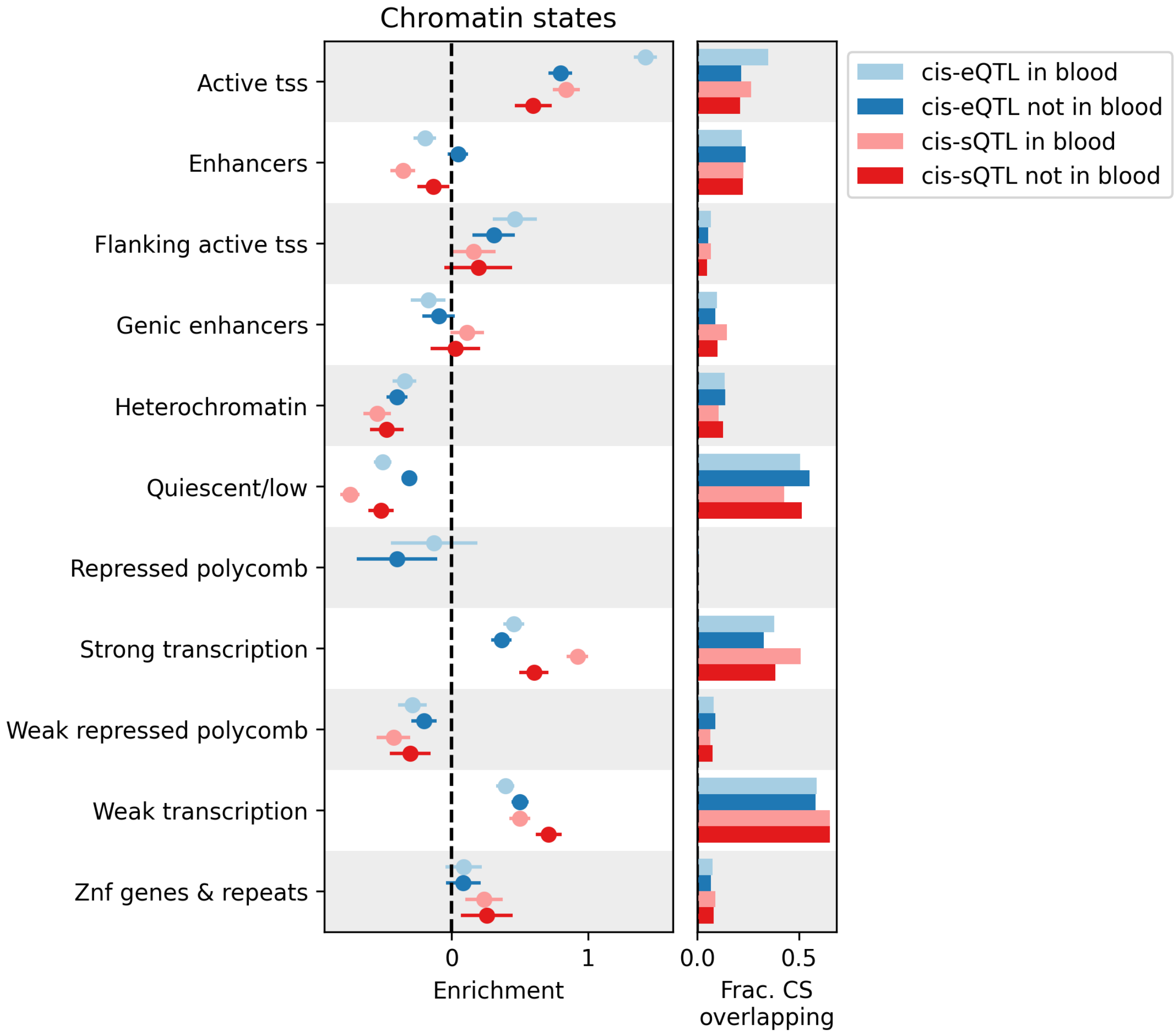


**Figure S26.** Enrichment of lung cis-e/sQTLs that are/are not shared with whole blood, in Roadmap Epigenomics chromatin states. Error bars represent 95% confidence intervals.

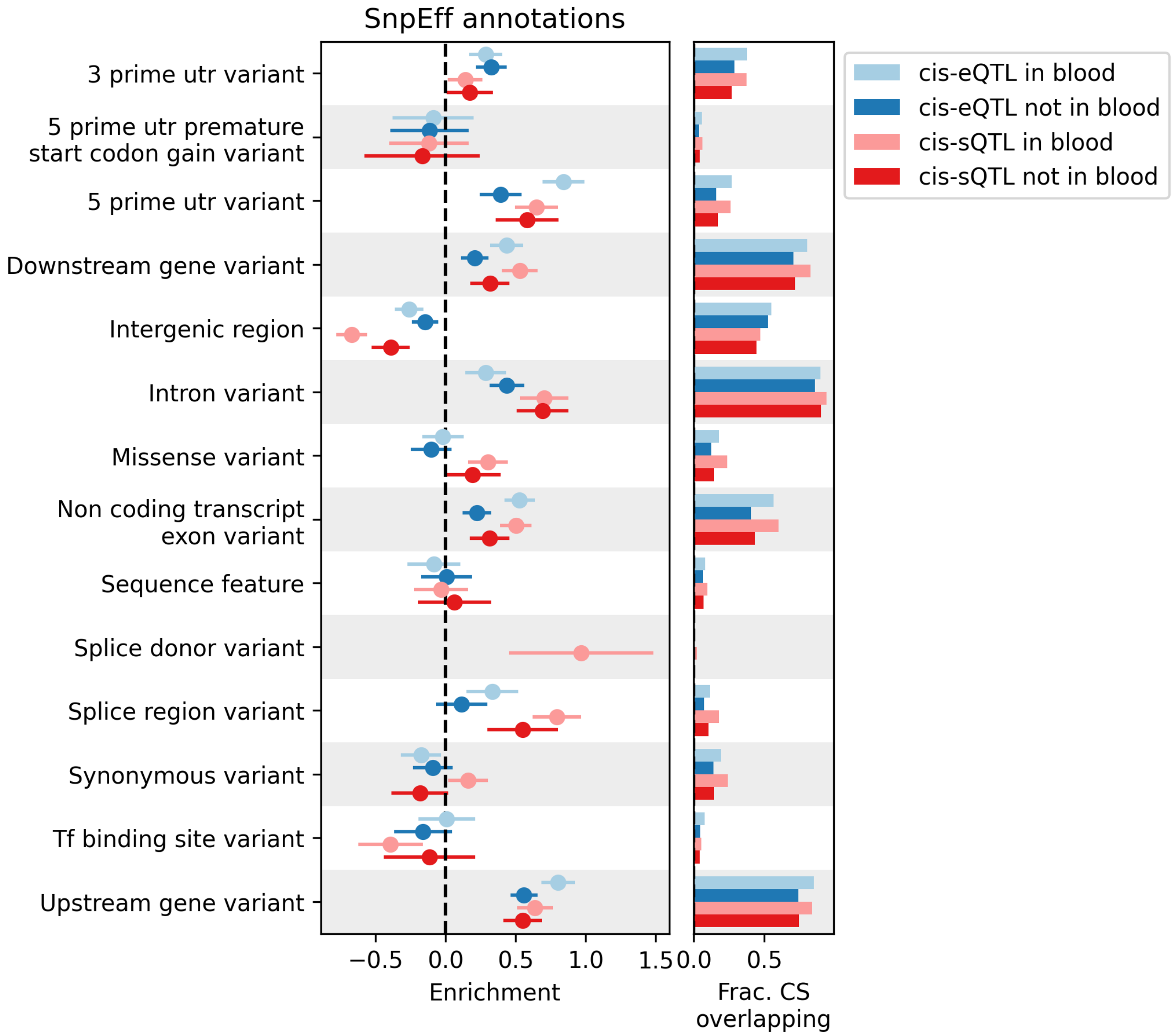


**Figure S27.** Enrichment of nasal epithelial cis-e/sQTLs that are/are not shared with whole blood, in SnpEff annotations. Error bars represent 95% confidence intervals.

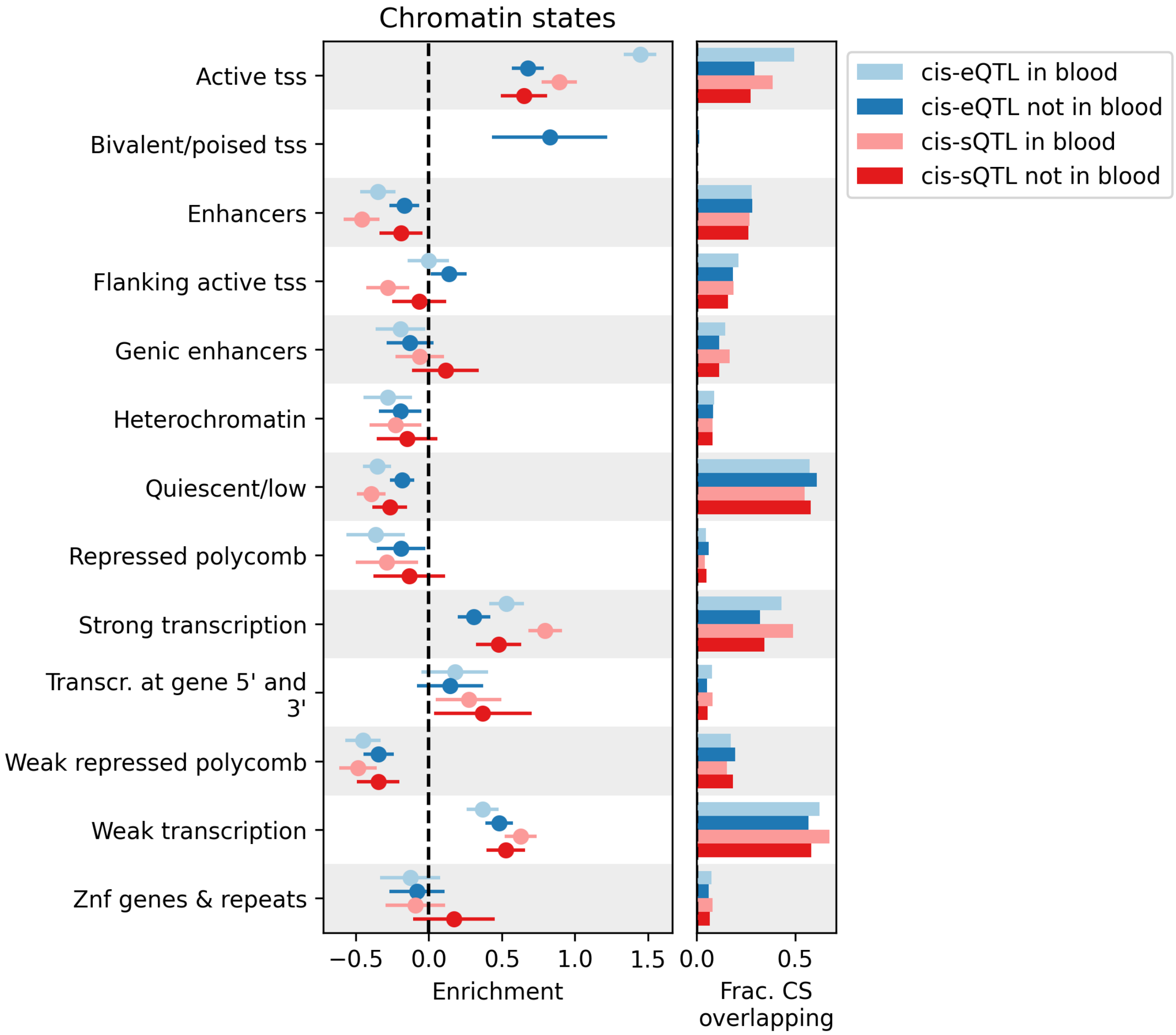


**Figure S28.** Enrichment of nasal epithelial cis-e/sQTLs that are/are not shared with whole blood, in Roadmap Epigenomics chromatin states. Error bars represent 95% confidence intervals.

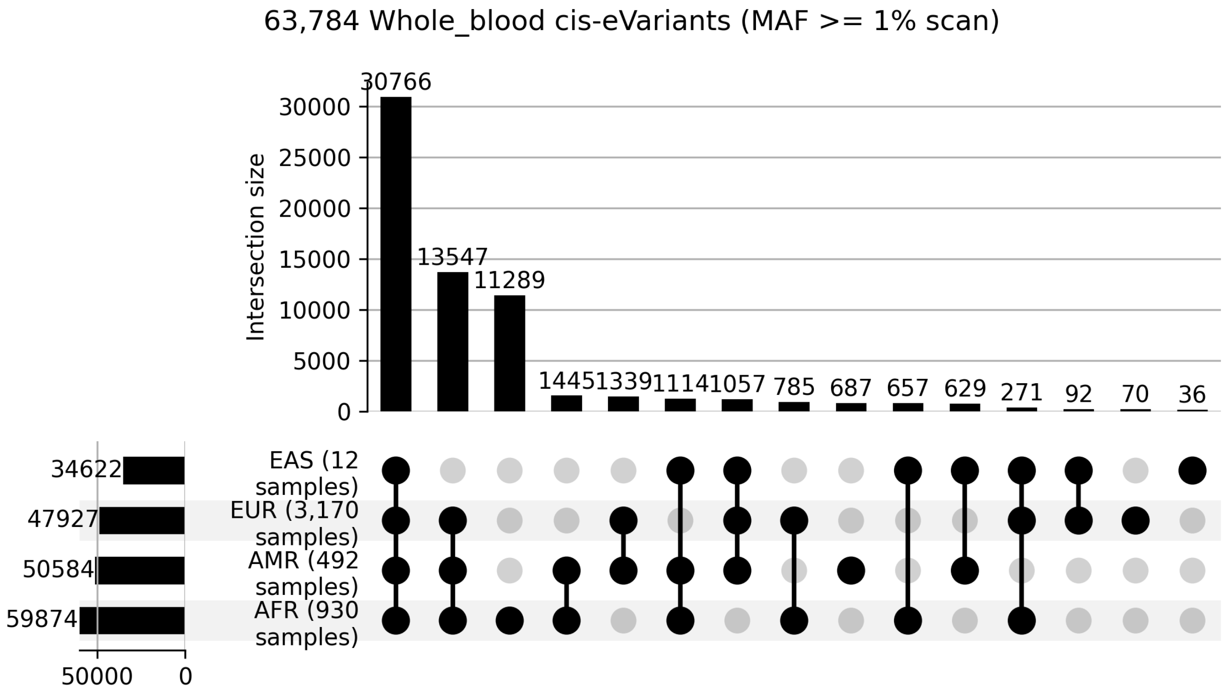


**Figure S29.** cis-eQTL variant presense in each ancestry. A sample was assigned to an ancestry if a sample's global ancestry is at least 75% from that ancestry, or 50% in the case of AMR (see methods; samples not meeting these thresholds were left unassigned and omitted). Only ancestries with at least 10 samples are plotted. Each credible set is represented by it's top PIP variant. A variant was considered present in the ancestry if it's MAF was $\geq 0.1 * x$, where $x$ is the max MAF across the displayed ancestries.

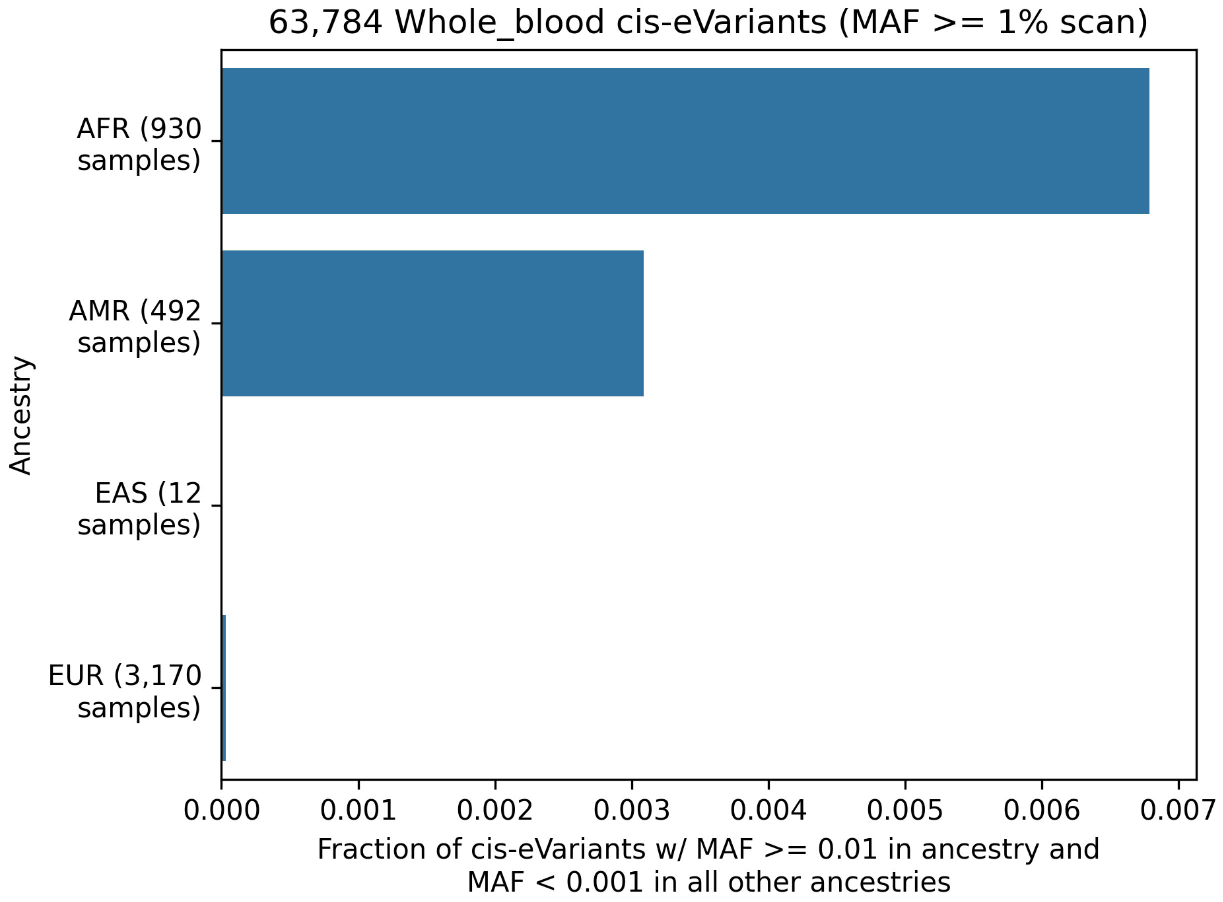


**Figure S30.** Fraction of whole blood cis-eVariants that are common in a given ancestry (MAF $\geq 0.01$) and rare in the others (MAF $\leq 0.001$). Sample ancestry was assigned as in Fig. **S29.**

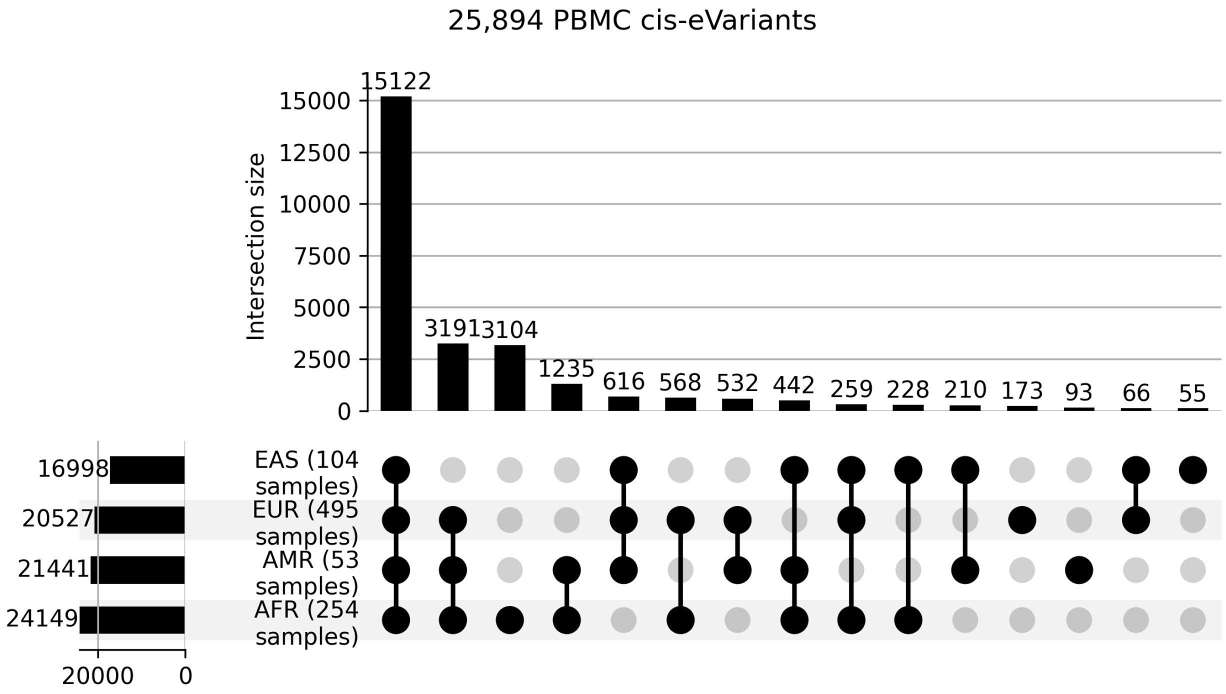


**Figure S31.** PBMC cis-eVariant presense in each ancestry. Sample ancestry was assigned as in Fig. **S29.**

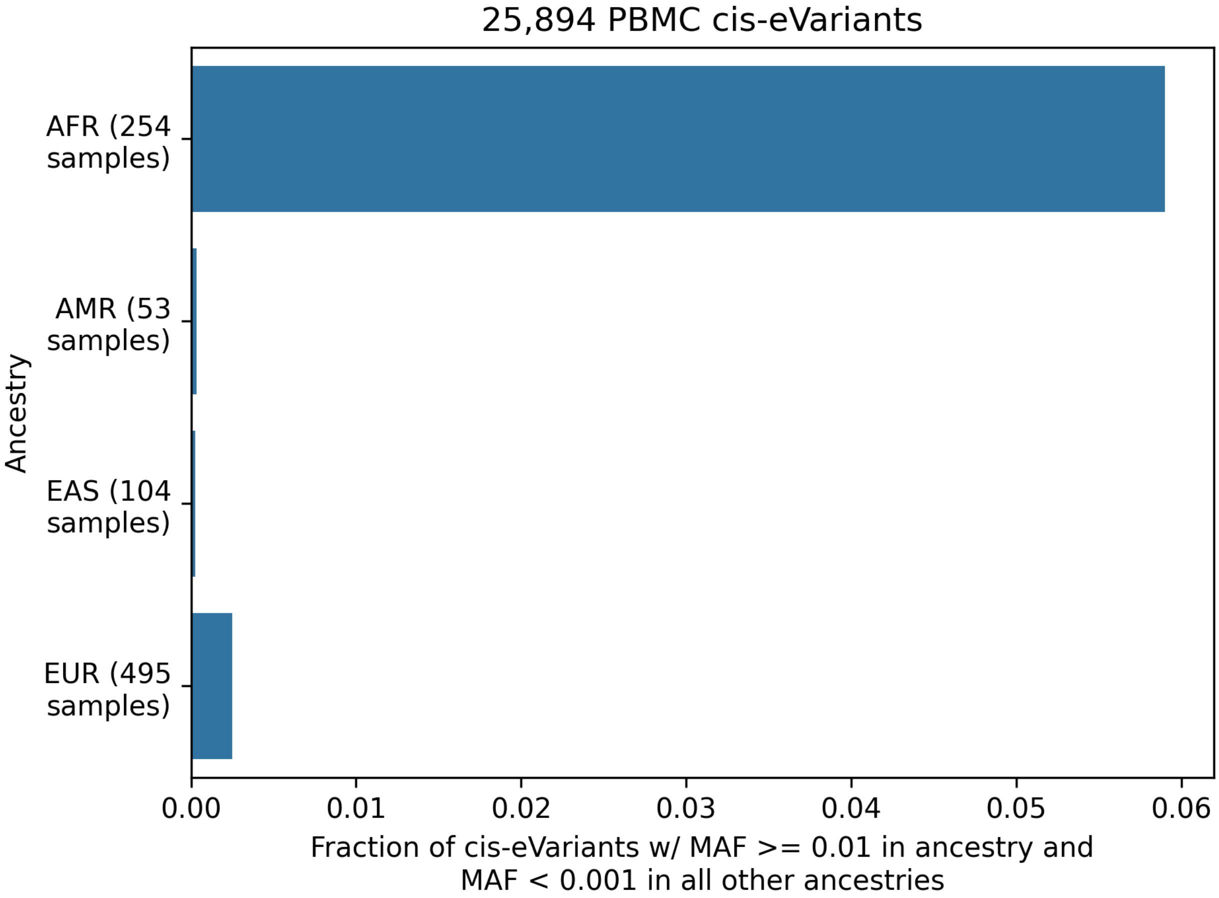


**Figure S32.** Fraction of PBMC cis-eVariants that are common in a given ancestry (MAF $\geq 0.01$) and rare in the others (MAF $\leq 0.001$). Sample ancestry was assigned as in Fig. **S29.**

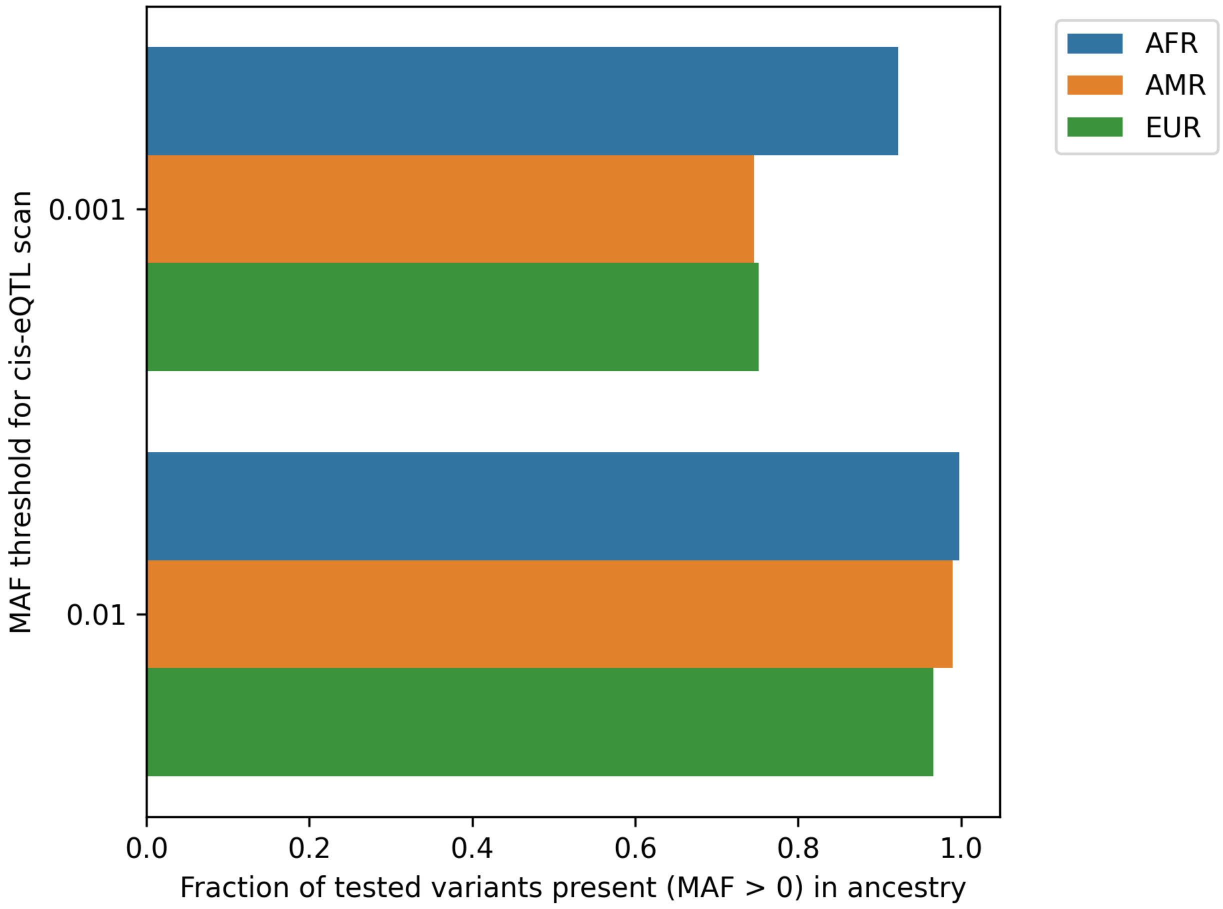


**Figure S33.** Fraction of variants tested in whole blood MAF $\geq 0.01$ and MAF $\geq 0.001$ cis-eQTL scans that are present in each ancestry. Sample ancestry was assigned as in Fig. **S29.**

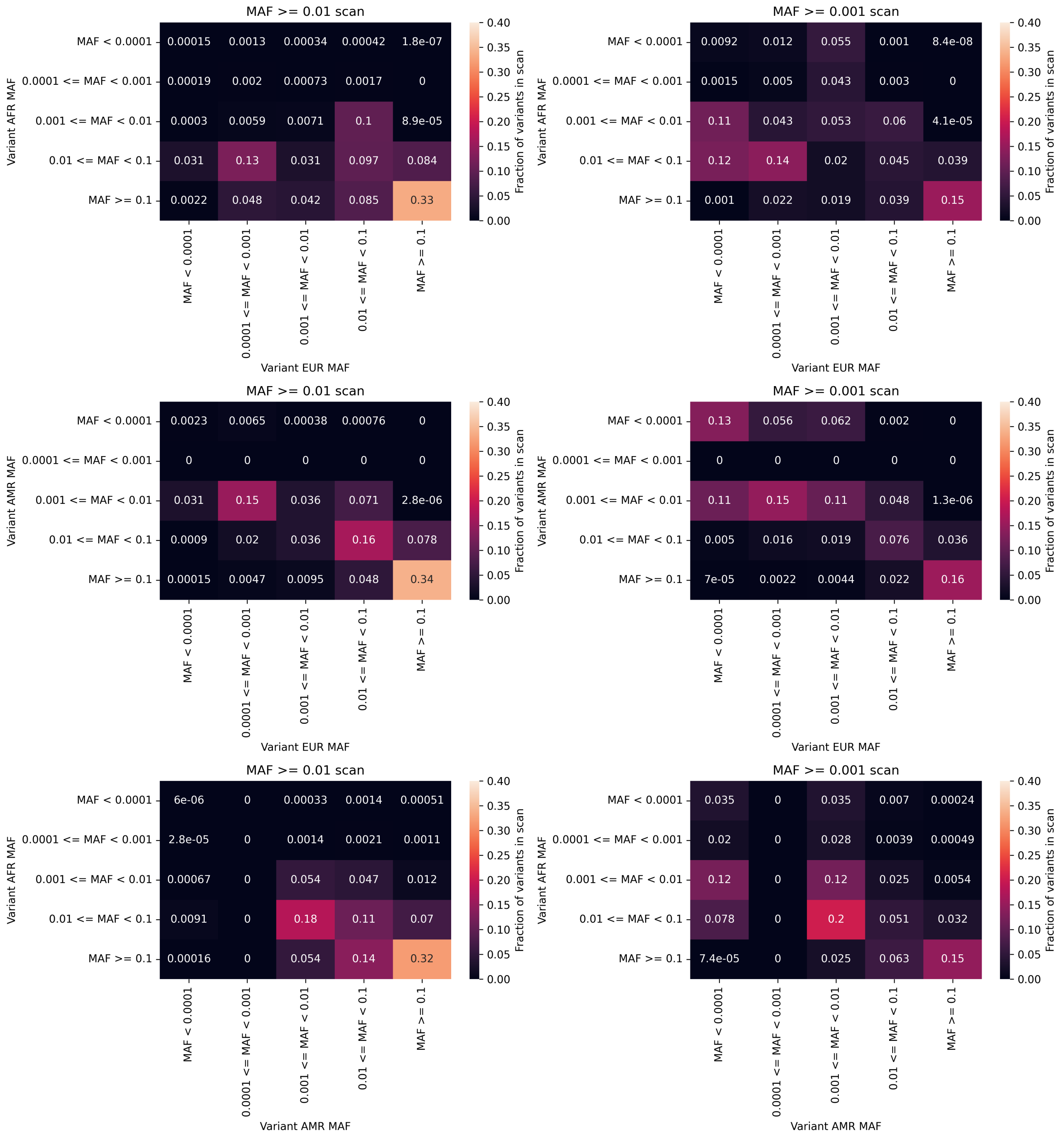


**Figure S34.** Binned ancestry-specific MAF for variants tested in whole blood MAF $\geq 0.01$ and MAF $\geq 0.001$ cis-eQTL scans. Sample ancestry was assigned as in Fig. **S29.**

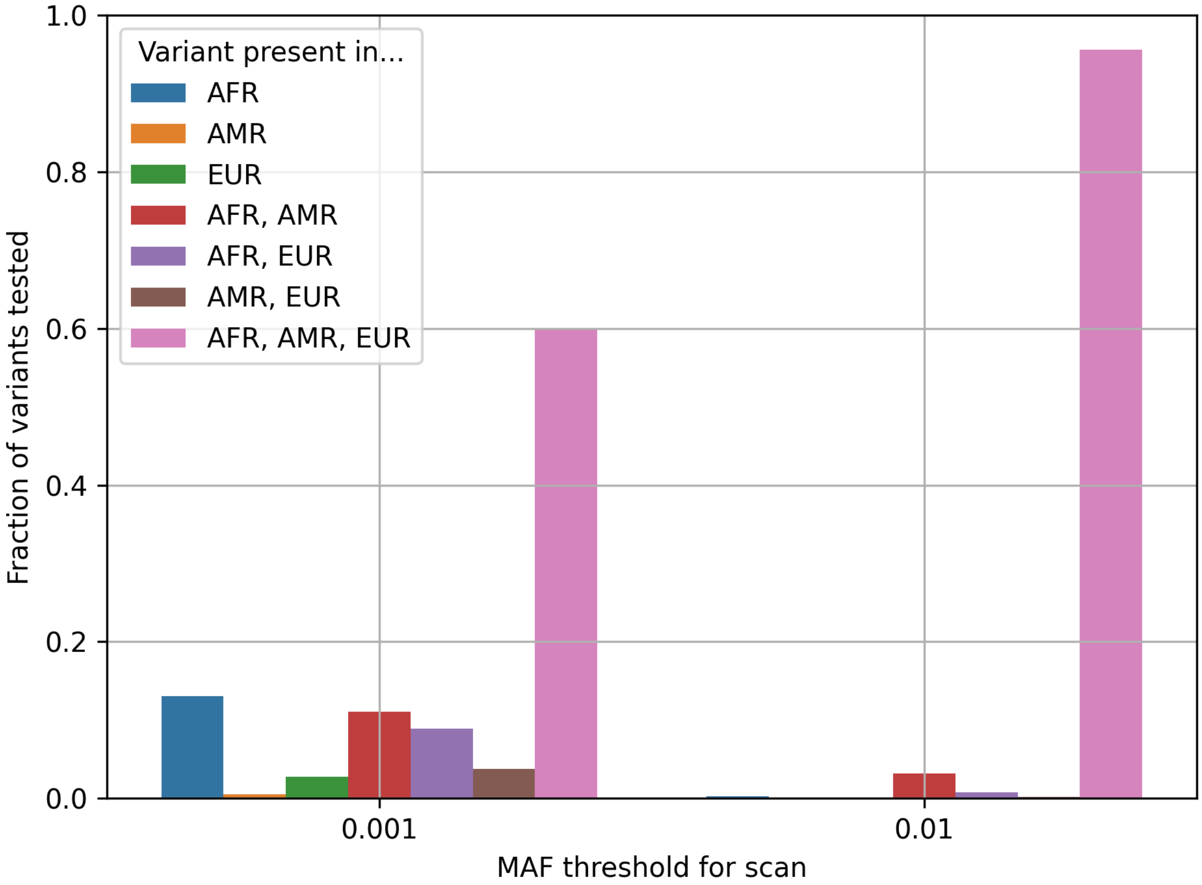


**Figure S35.** Fraction of tested variants present in each ancestry/set of ancestries in whole blood MAF $\geq$ 0.01 and MAF $\geq$ 0.001 cis-eQTL scans. Sample ancestry was assigned as in Fig. **S29.** Variants with MAF <0.001 in all of EUR, AFR, and AMR ancestries were omitted.

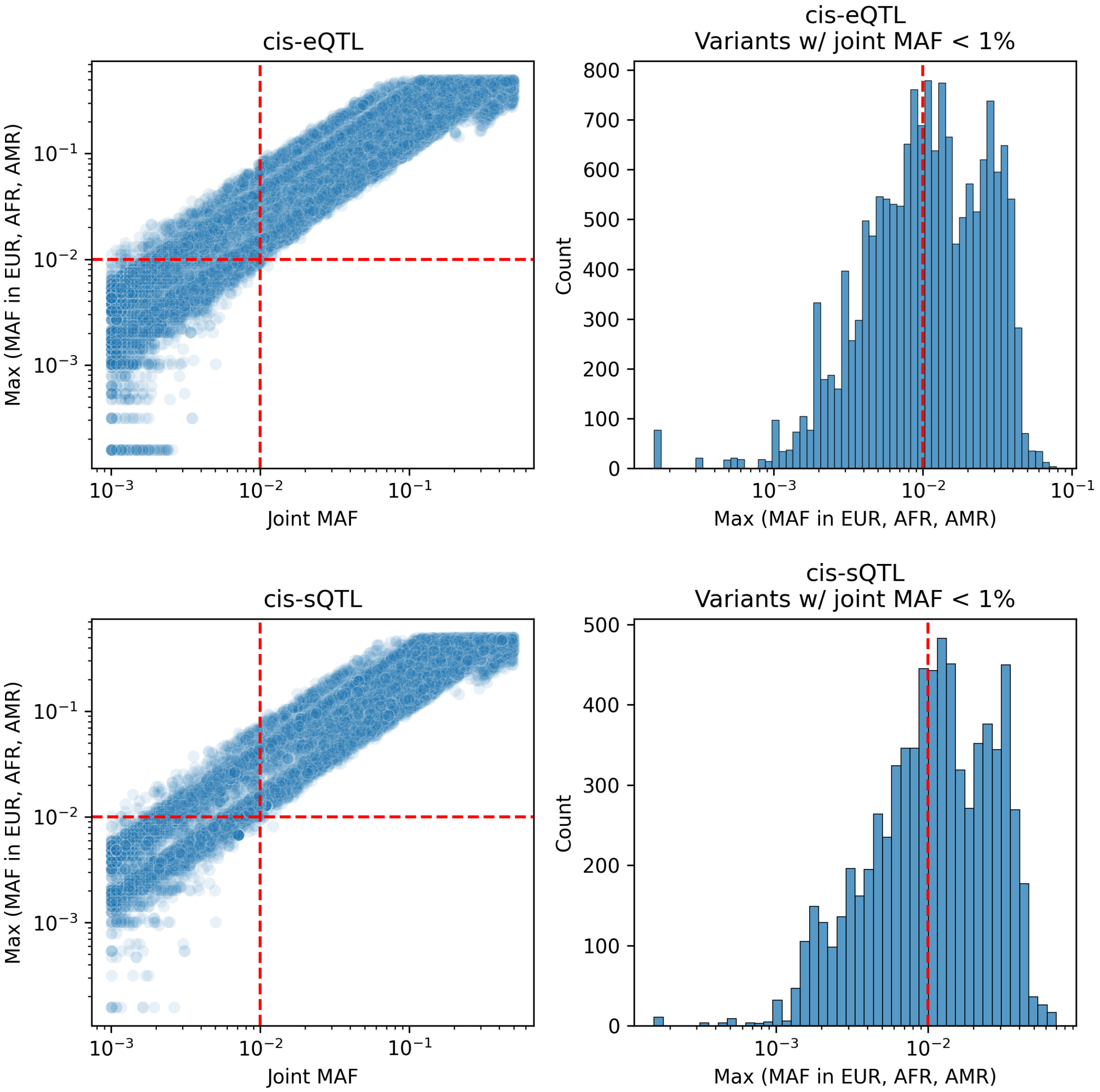


**Figure S36.** Left panels: joint MAF (MAF across all scan samples) for each whole blood cis-e/sQTL in the MAF $\geq 0.001$ scans (top PIP variant per credible set), vs the maximum MAF across EUR, AFR, and AMR ancestry samples. Right panels: Maximum MAF across EUR, AFR, and AMR ancestry samples for those cis-e/sQTL with MAF $< 0.01$. Sample ancestry was assigned as in Fig. **S29.** 135 variants with max MAF $= 0$ across EUR, AFR, and AMR ancestries were omitted.

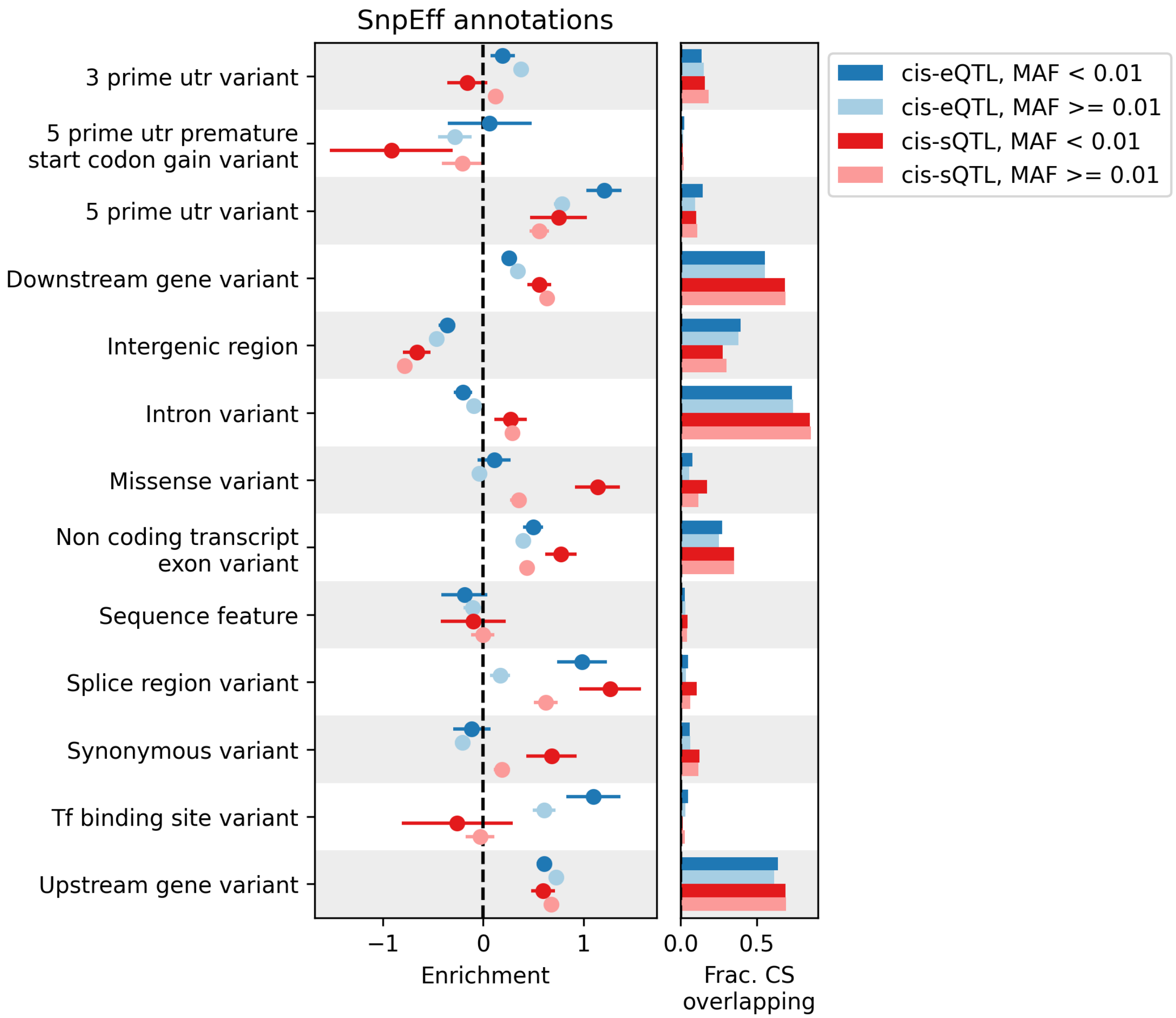


**Figure S37.** Enrichment of rare (MAF < 0.01 in EUR, AFR, and AMR individually) and common (MAF $\geq 0.01$ in EUR, AFR, and AMR individually) cis-e/sQTLs, in SnpEff annotations. Enrichment calculated relative to control credible sets matched on chromosome, MAF, LD, and number of genes tested against, using logistic regression (see Methods; enrichment = logistic regression coefficient; error bars represent 95% confidence intervals).

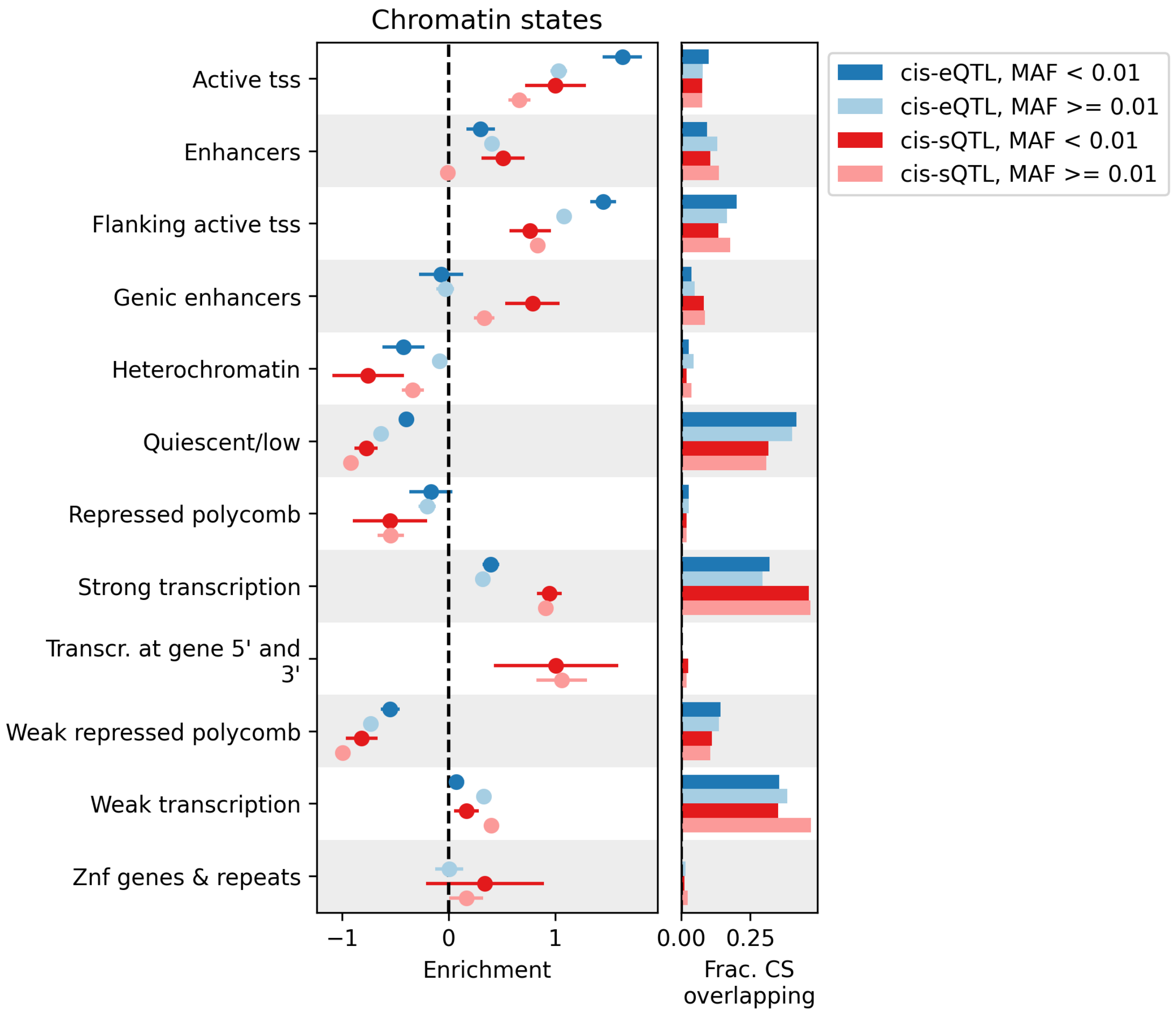


**Figure S38.** Enrichment of rare (MAF < 0.01 in EUR, AFR, and AMR individually) and common (MAF $\geq$ 0.01 in EUR, AFR, and AMR individually) cis-e/sQTLs, in Roadmap Epigenomics chromatin states. Error bars represent 95% confidence intervals.

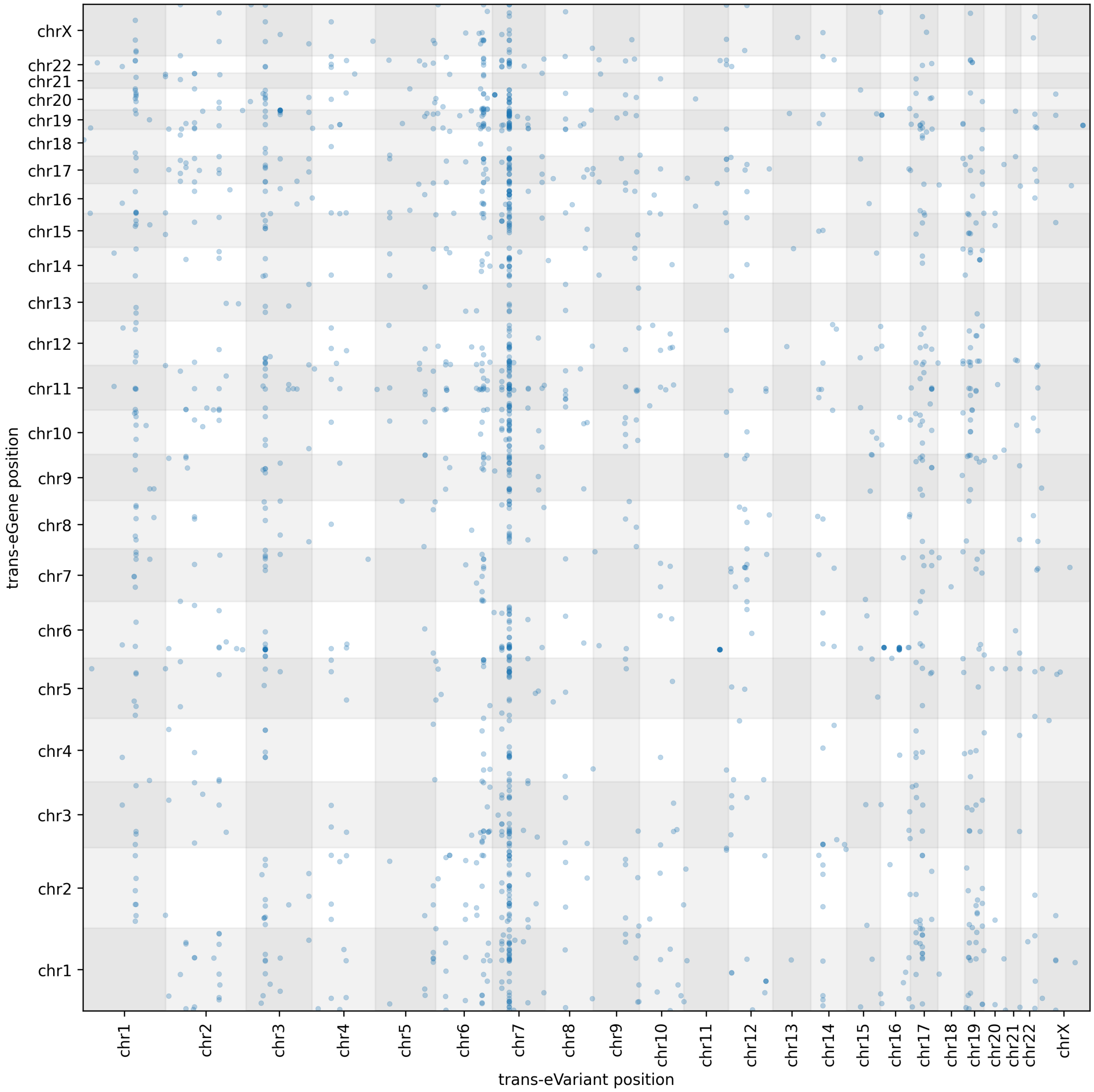


**Figure S39.** Whole blood primary trans-eQTL gene and variant positions.

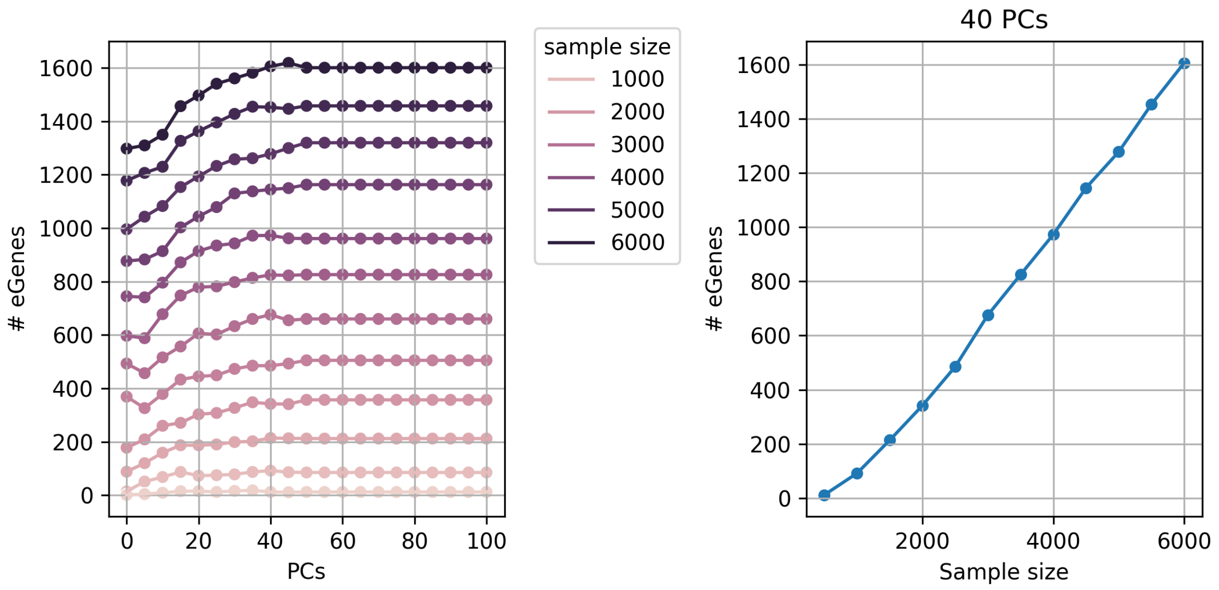


**Figure S40.** Trans-eQTL saturation analysis (trans-eGene discovery) using nested subsets of whole blood samples.

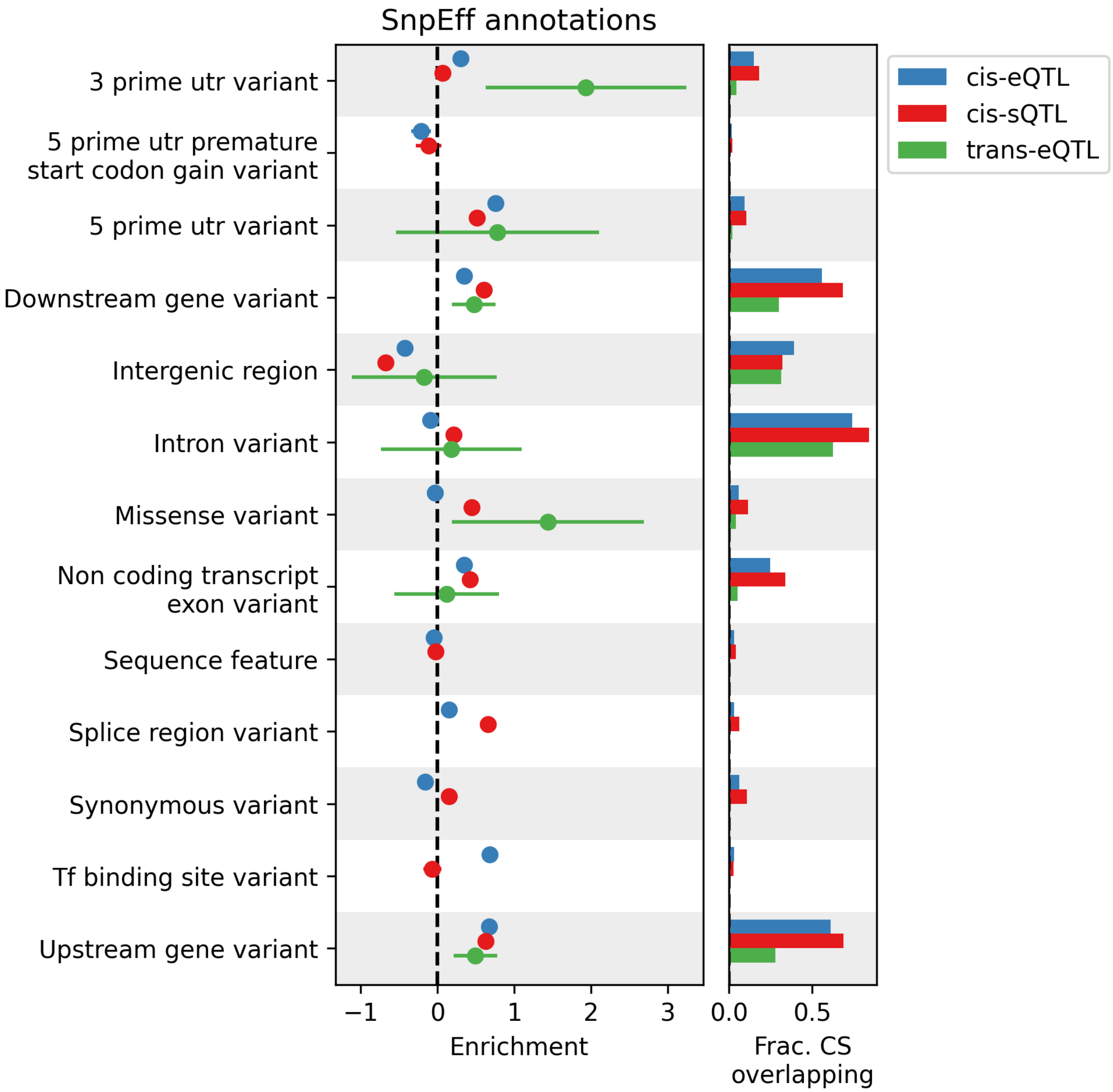


**Figure S41.** Enrichment of whole blood cis-e/s credible sets and trans primary hits, in SnpEff annotations. Error bars represent 95% confidence intervals.

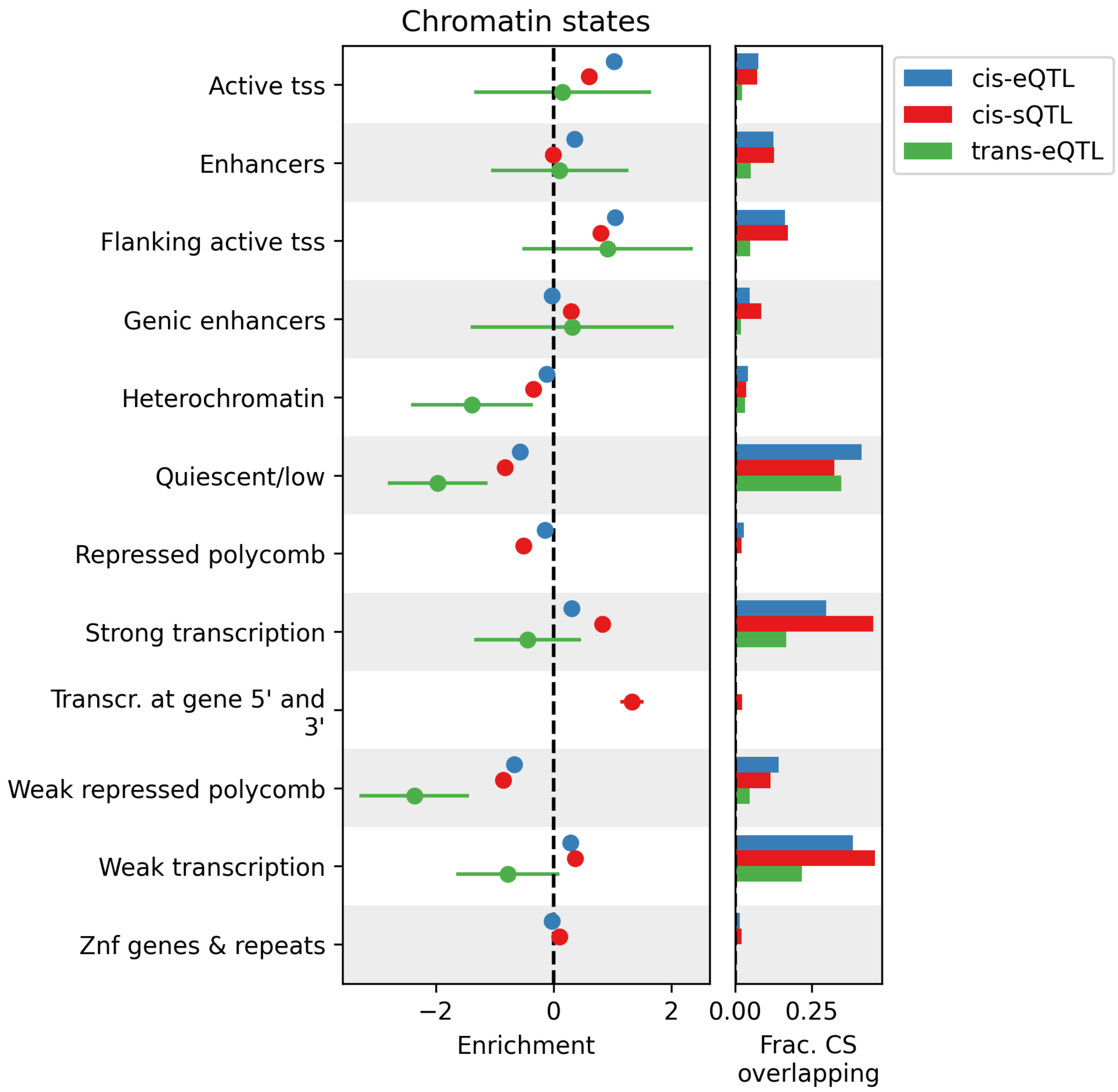


**Figure S42.** Enrichment of whole blood cis-e/s credible sets and trans primary hits, in Roadmap Epigenomics chromatin states. Error bars represent 95% confidence intervals. Trans-sQTL is excluded due to failure of the model to converge.

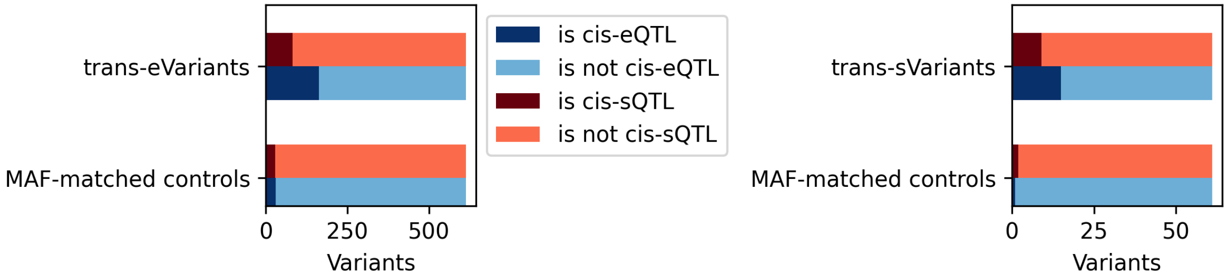


**Figure S43.** Enrichment of whole blood trans-e/sQTL primary hits in cis-e/sQTL credible sets.

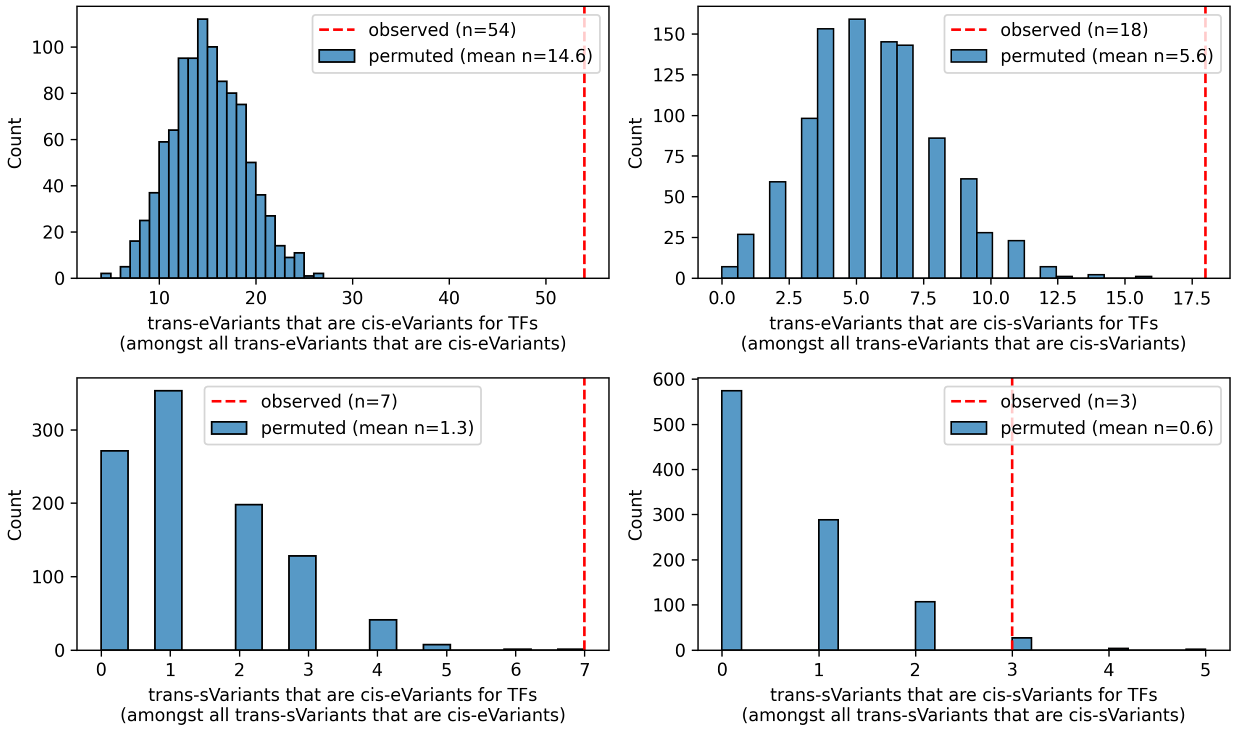


**Figure S44.** Whole blood cis-eGenes with cis-eQTL overlapping trans-e/sQTL are enriched for TF genes.

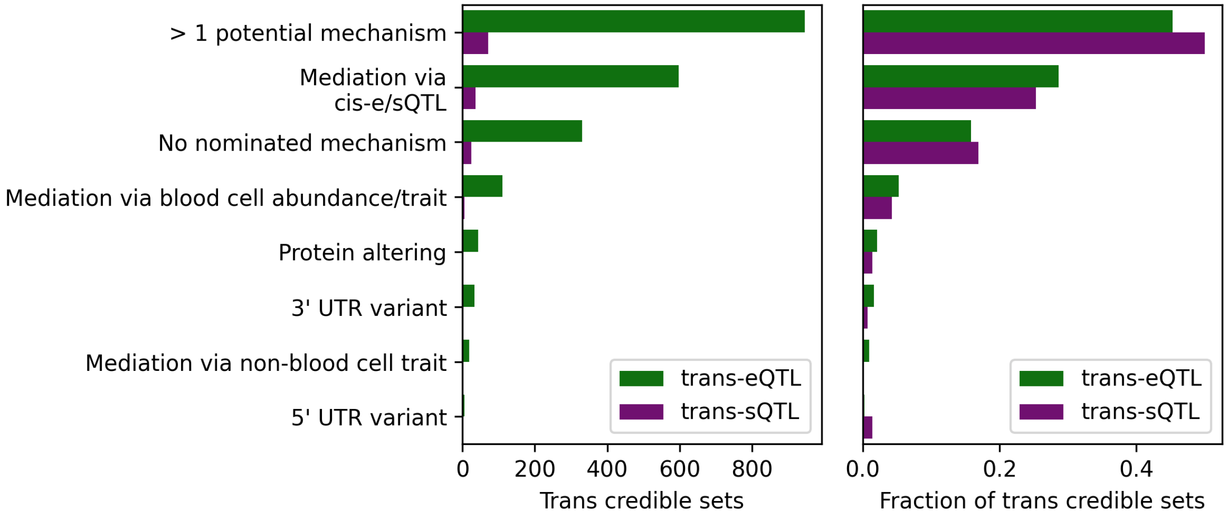


**Figure S45.** Nominated mechanisms of effect for whole blood trans-e/sQTL signals.

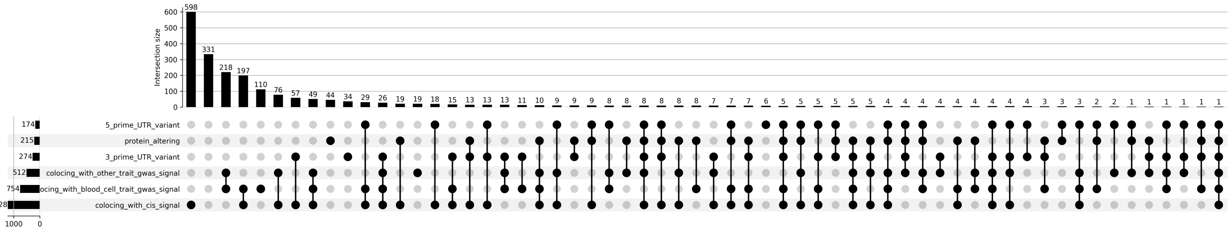


**Figure S46.** Nominated mechanisms of effect for whole blood trans-eQTL signals.

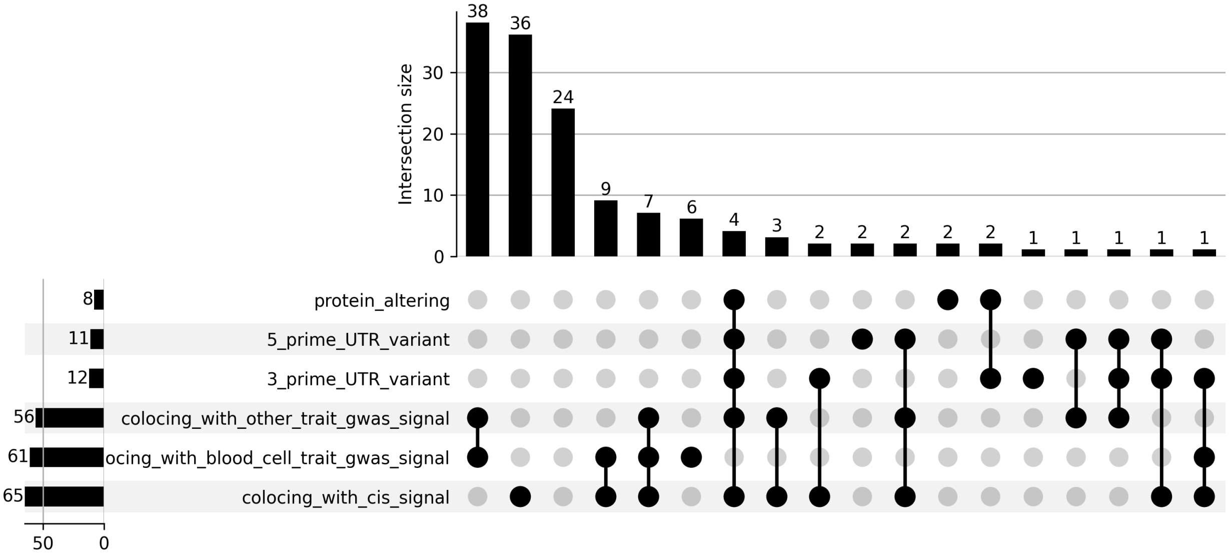


**Figure S47.** Nominated mechanisms of effect for whole blood trans-sQTL signals.

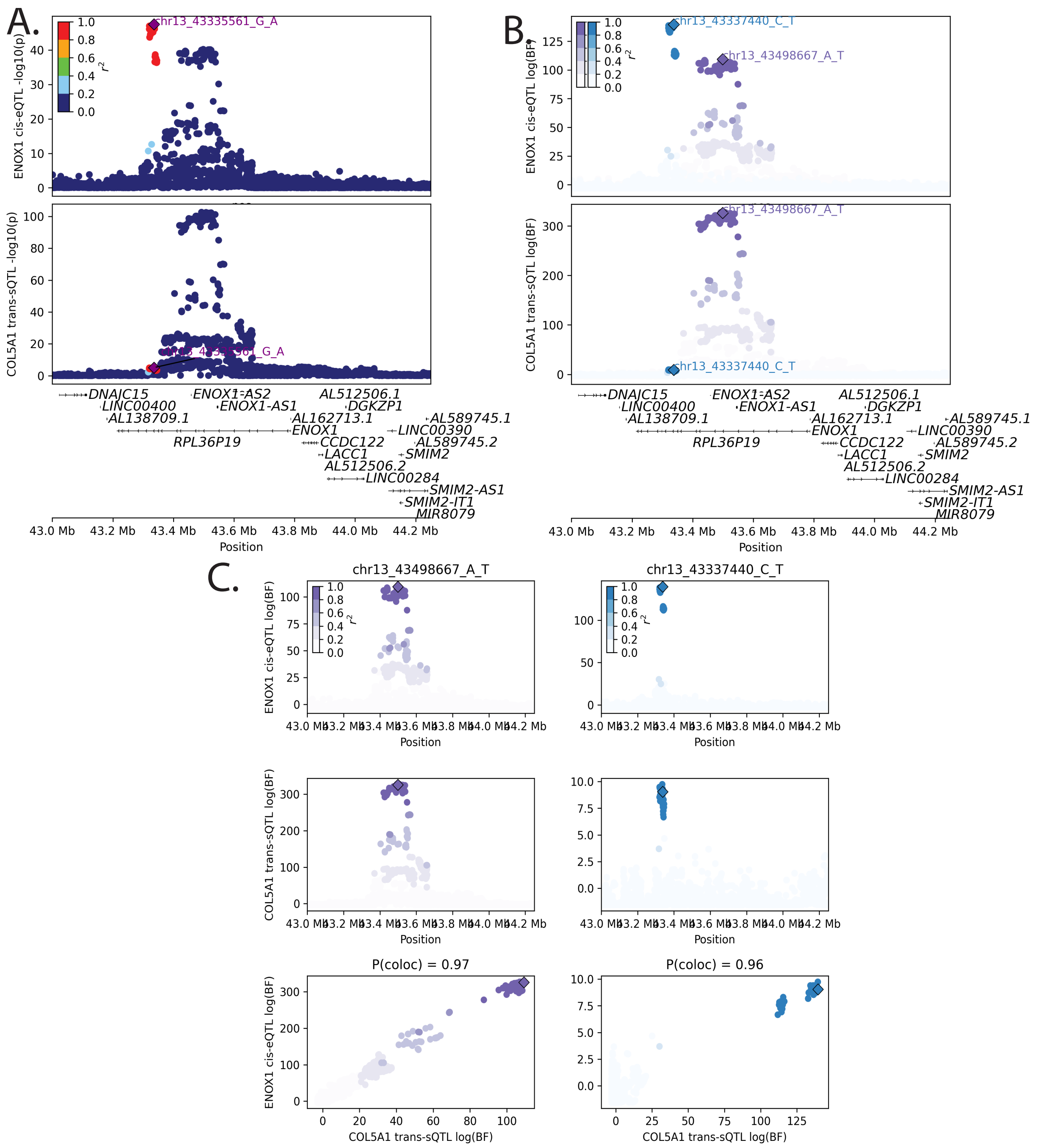


**Figure S48.** Colocalizations between two *ENOX1* cis-eQTL signals and two *COL5A1* trans-sQTL signals. (A) nominal p-values, (B) and (C) log10(Bayes factors) for each of the colocalizing signals

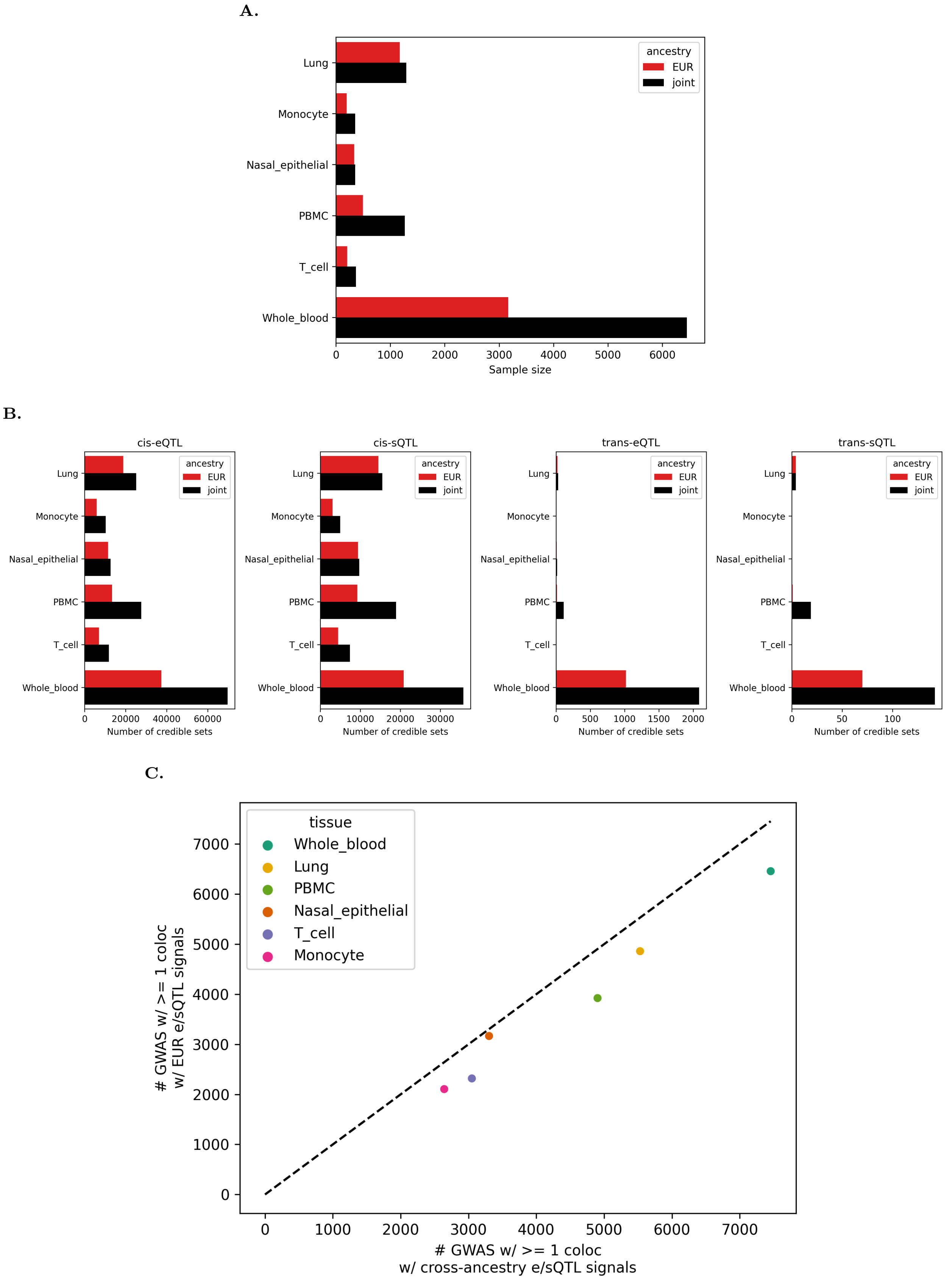


**Figure S49.** A. Sample size for cross-ancestry ('joint') e/sQTL scans or EUR-specific e/sQTL scans. B. Number of independent signals (SuSiE credible sets) discovered in EUR-specific vs cross-ancestry scans. C. Number of GWAS signals colocalizing with at least one e/sQTL signal in each tissue in EUR-specific vs cross-ancestry scans

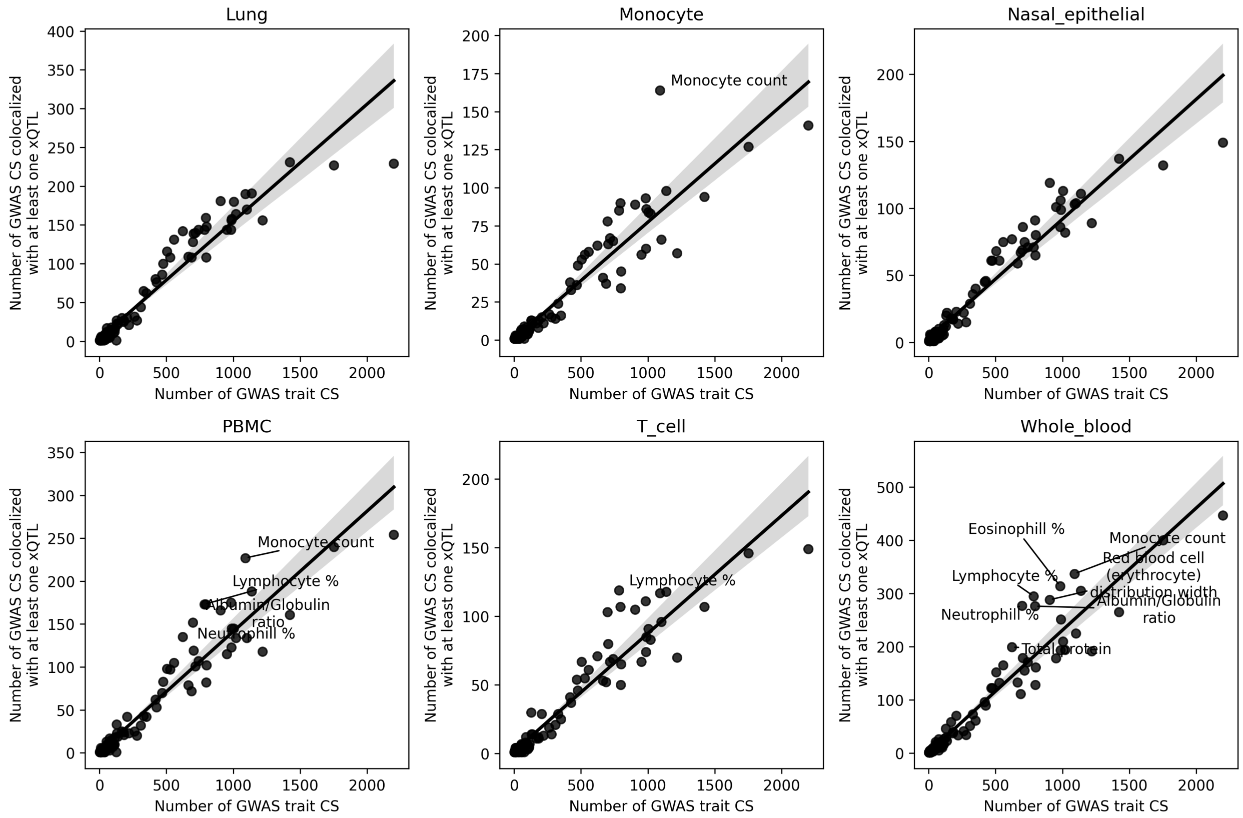


**Figure S50.** Number of credible sets per GWAS trait vs number of GWAS credible sets colocalizing with at least one e/sQTL signal. The largest deviations from expectation are labeled.

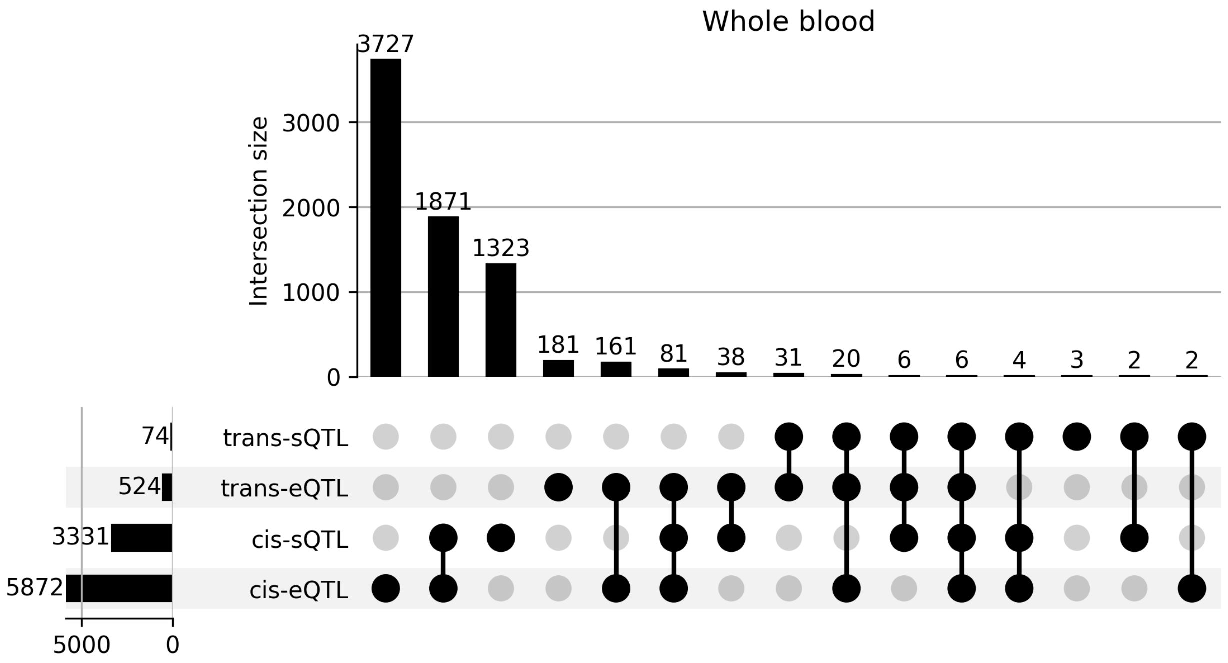


**Figure S51.** Whole blood xQTL modalities colocalizing with each GWAS signal.

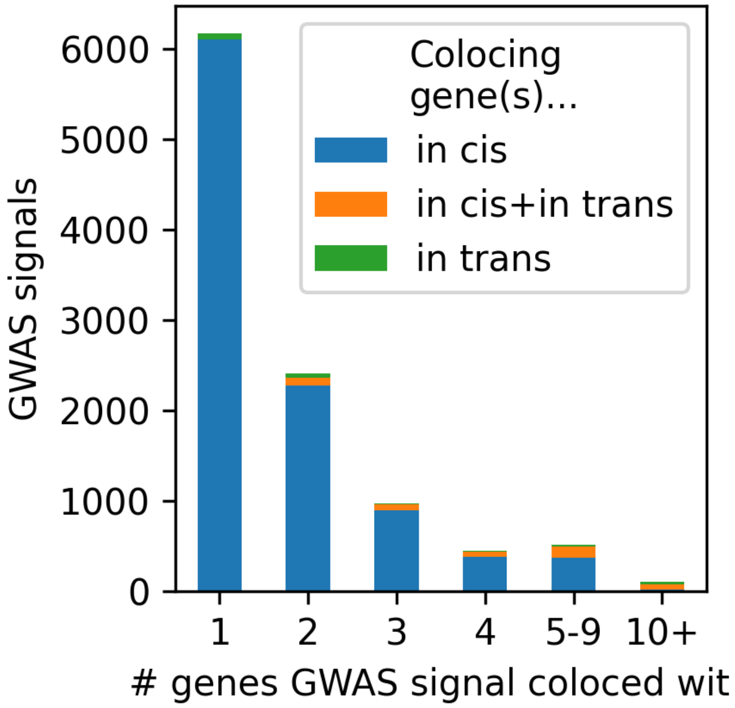


**Figure S52.** Number of genes represented amongst the e/sQTL signals each GWAS signal colocalizes with.

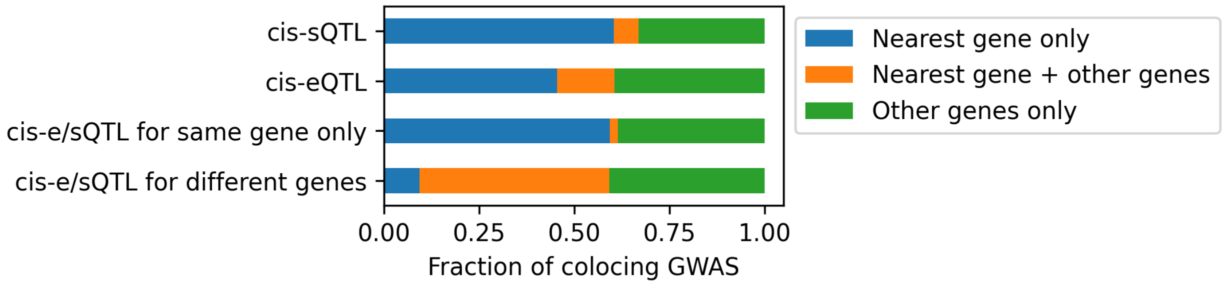


**Figure S53.** For GWAS signals colocalizing with cis-e/sQTL signals, how often does that colocalization involve the nearest gene. cis-eQTL = cis-eQTL signals that are not also cis-sQTL signals (based on colocalization of cis-e/sQTL signals); cis-sQTL = cis-sQTL signals that are not also cis-eQTL signals; cis-e/sQTL for same gene only = signals that are both cis-e and cis-sQTL signals for the same gene (and are not cis signals for any other genes); cis-e/sQTL for different genes = cis-e/sQTL signals that are cis-eQTL signals for at least one gene and cis-sQTL signals for at least one other gene.

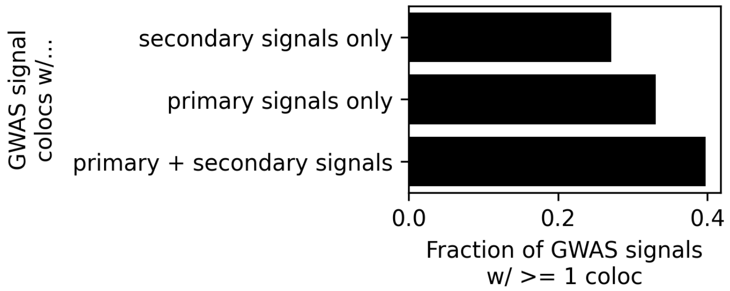


**Figure S54.** Fraction of GWAS signals with at least one colocalization colocalizing with only primary e/sQTL signals, only secondary/tertiary/etc. e/sQTL signals, or both, across all tissues.

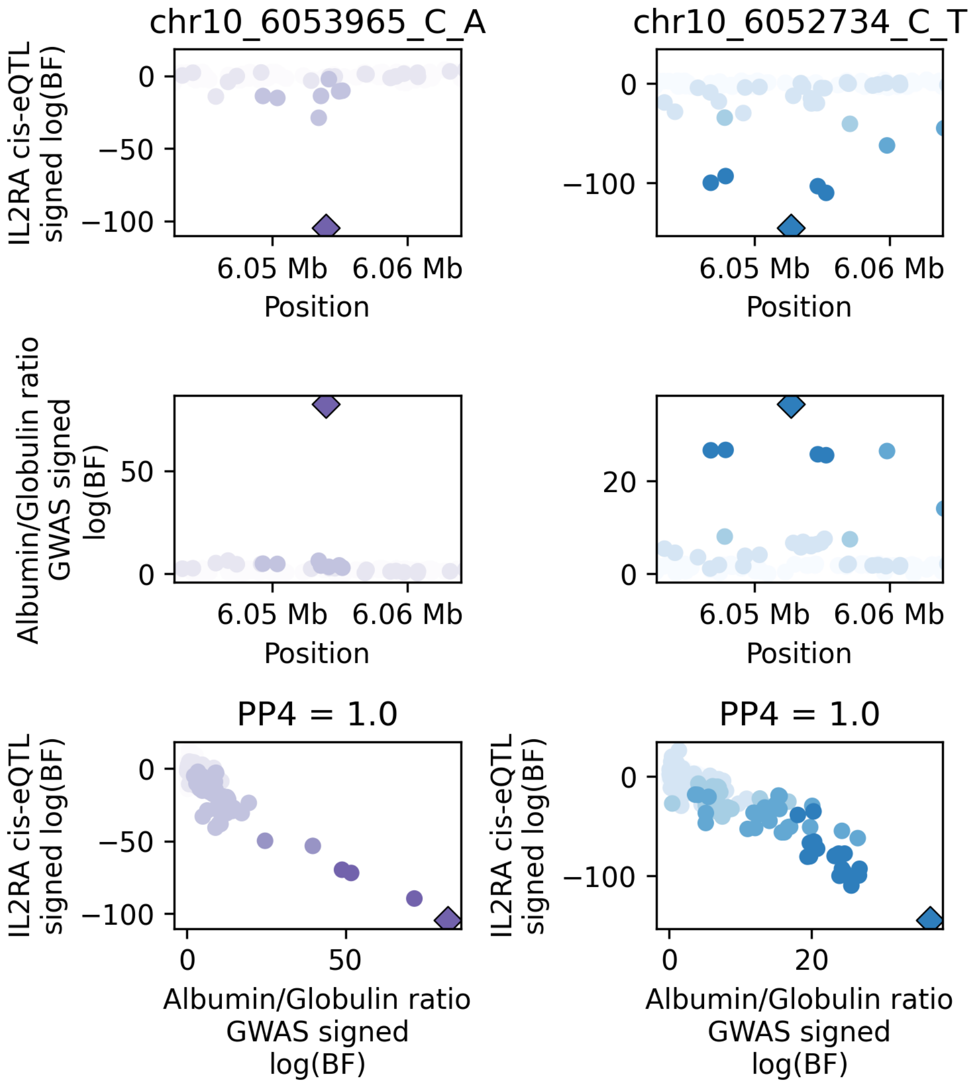


**Figure S55.** Colocalizations between two *IL2RA* cis-eQTL signals and two albumin/globulin ratio GWAS signals (see Fig 4).

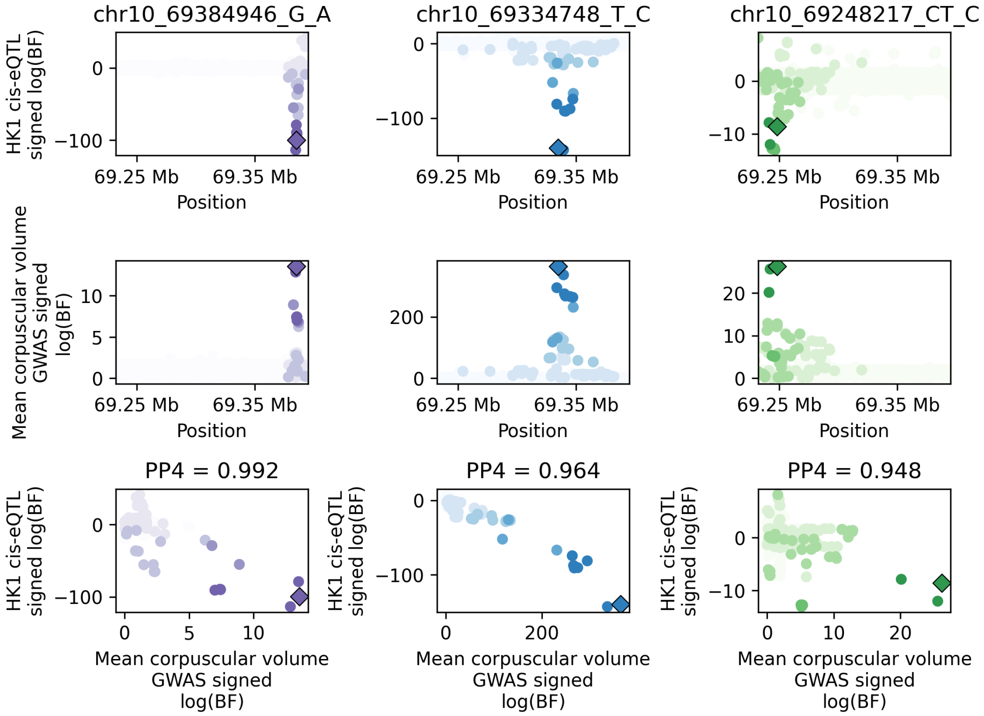


**Figure S56.** Colocalizations between three *HK1* cis-eQTL signals and three mean corpuscular volume GWAS signals (see Fig 4).

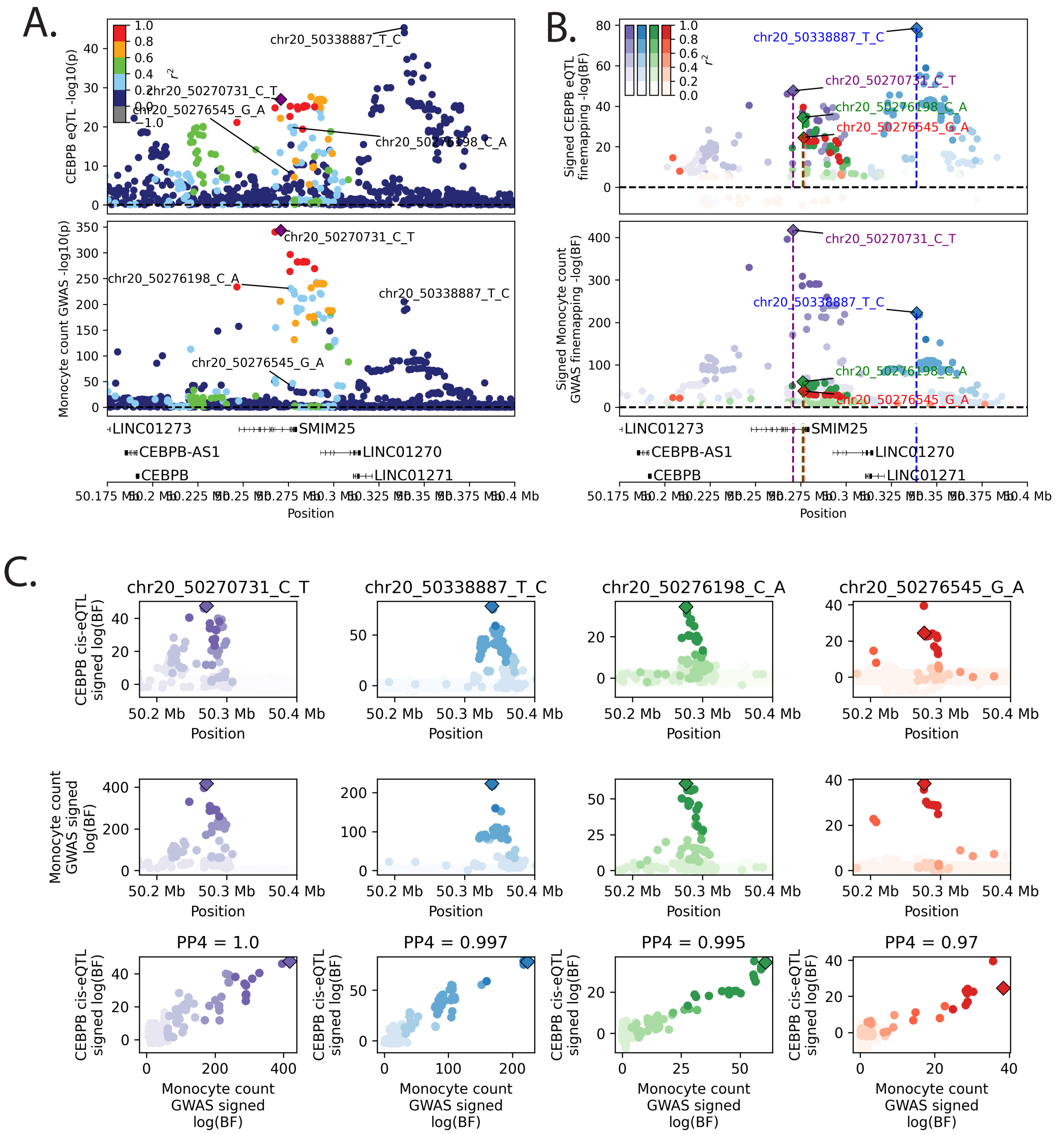


**Figure S57.** Colocalizations between four *CEBPB* cis-eQTL signals and four monocyte count GWAS signals.

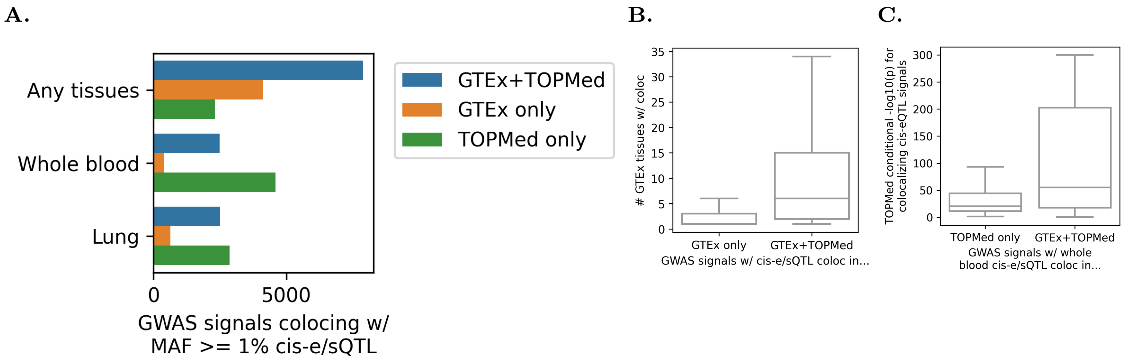


**Figure S58.** (A) Number of GWAS signals showing at least one cis-e/sQTL colocalization in TOPMed/GTEx whole blood, lung, or in any tissue. (B) GWAS signals colocalizing with GTEx but not TOPMed cis-e/sQTL tended to colocalize in a smaller number of GTEx tissues than those colocalizing with both GTEx and TOPMed cis-eQTL signals (C) GWAS signals colocalizing with TOPMed but not GTEx whole blood cis-e/sQTL signals tended to colocalize with weaker cis-eQTL signals than those colocalizing with both TOPMed and GTEx whole blood cis-e/sQTL signals

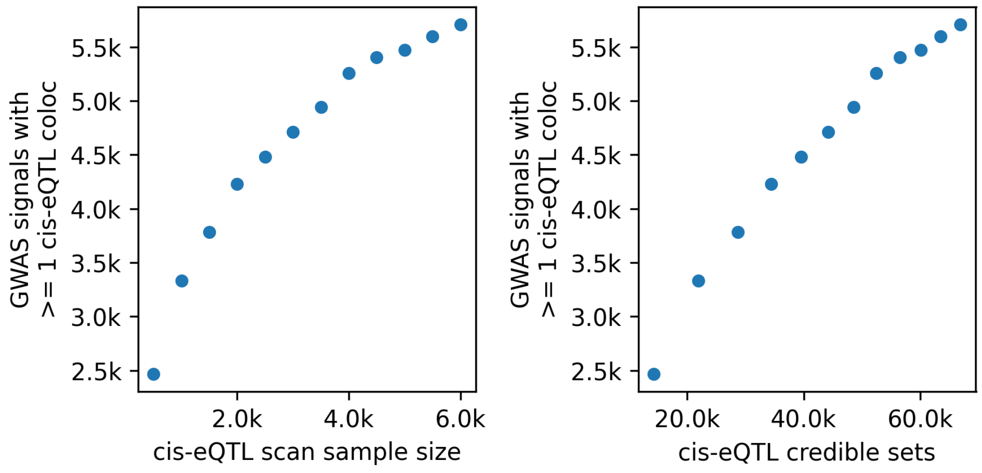


**Figure S59.** Number of GWAS signals colocalizing with whole blood cis-eQTL signals at different cis-eQTL sample sizes.

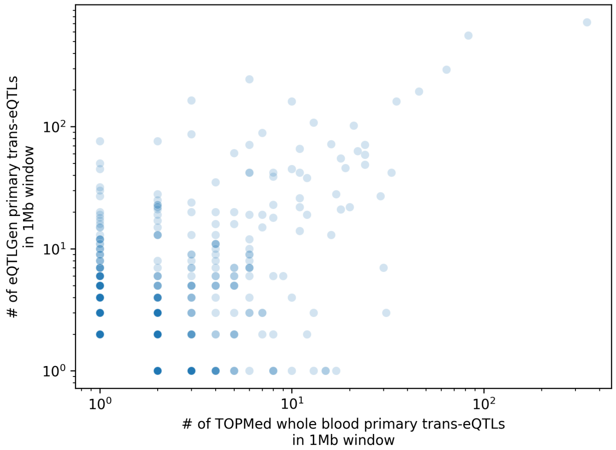


**Figure S60.** Primary whole blood trans-eQTLs per 1Mb window in eQTLGen and TOPMed.

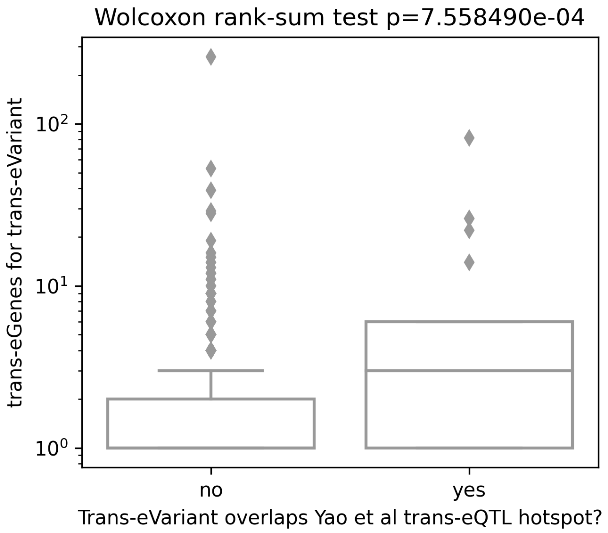


**Figure S61.** Trans-eVariants overlapping Yao et al 2017 trans-eQTL hotspots are associated with larger numbers of trans-eGenes than trans-eVariants outside of hotspots.

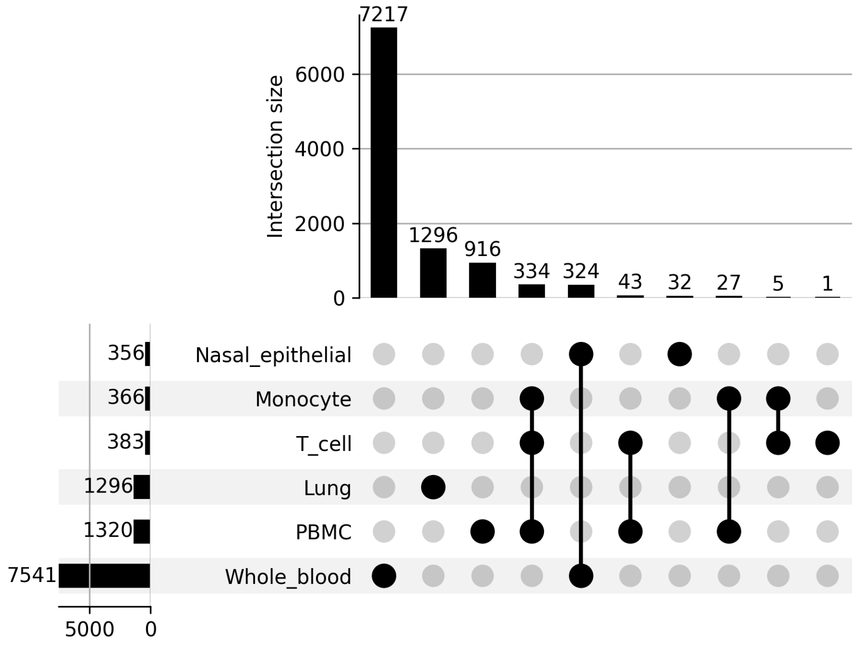


**Figure S62.** Cross-tissue donor representation.

**A.**

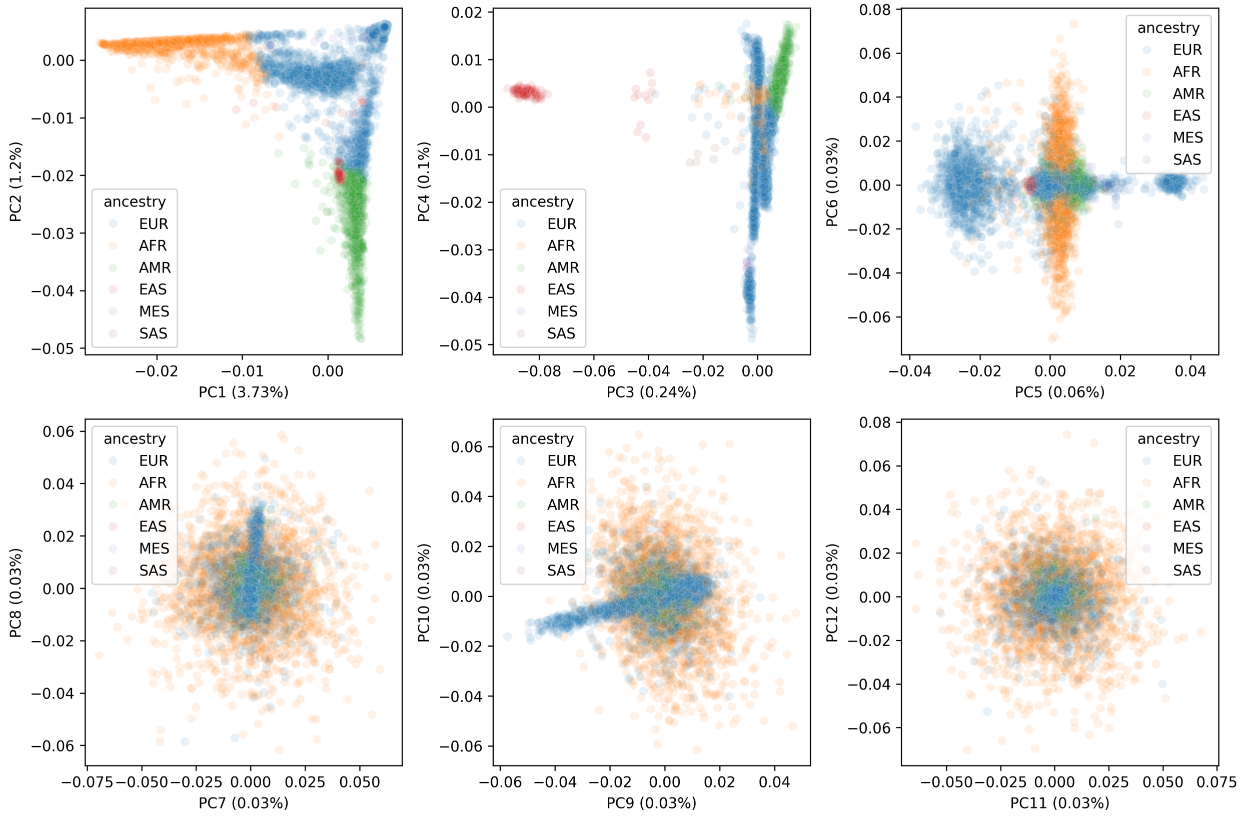


**B.**

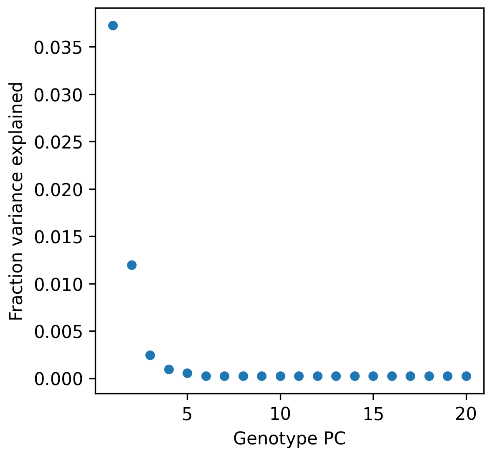


**Figure S63.** (A) Genotype PCA, colored by genetically-inferred ancestry (each sample assigned to the ancestry with the highest estimated percentage of global ancestry). (B) Screeplot for genotype PCA.

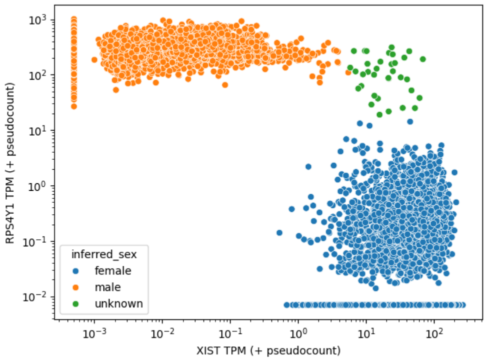


**Figure S64.** Sex inferred by gene expression.

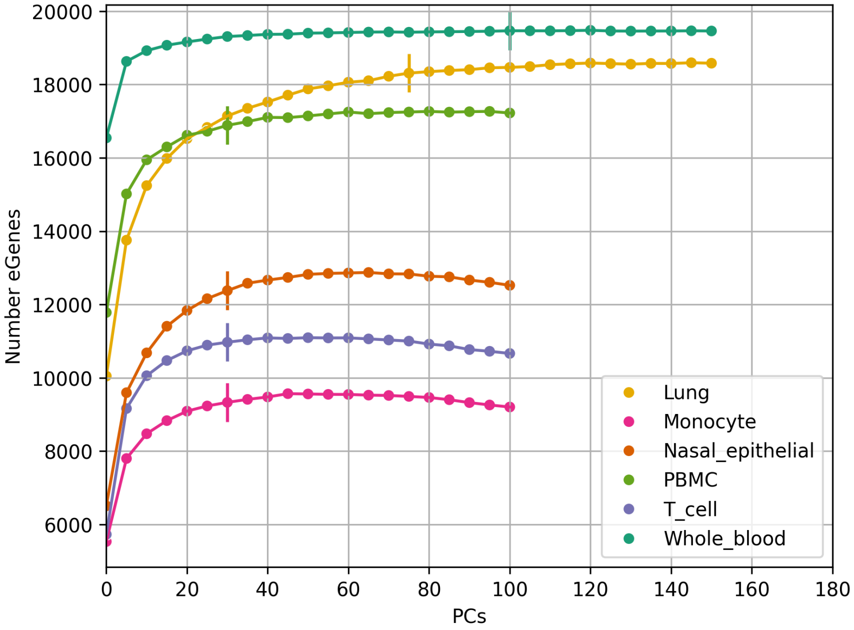


**Figure S65.** cis-eGene discovery as a function of gene expression PCs included as covariates.

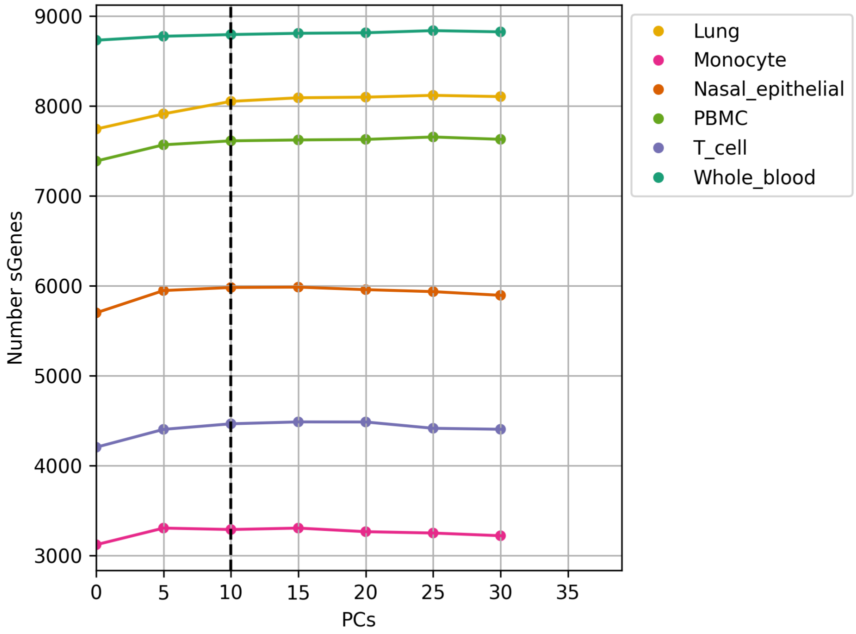


**Figure S66.** cis-sGene discovery as a function of splicing phenotype PCs included as covariates.

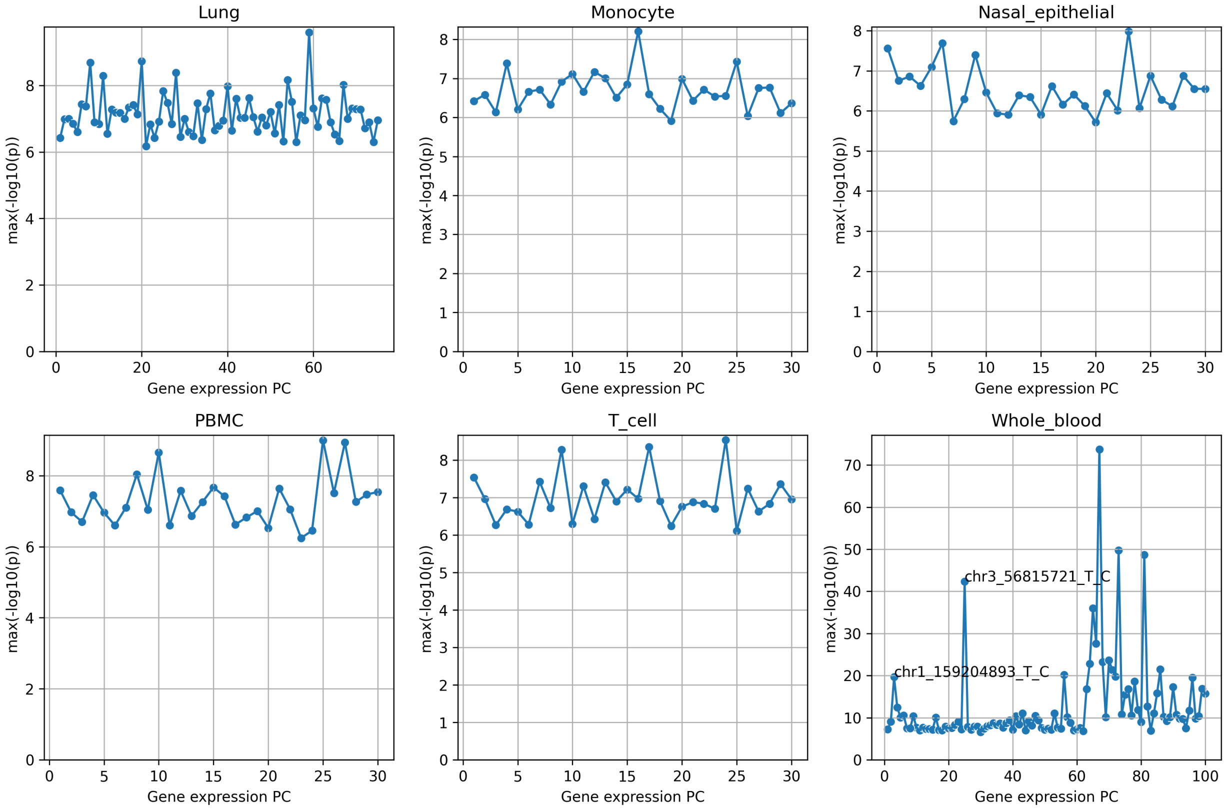


**Figure S67.** Strongest genetic associations observed with gene expression PCs.

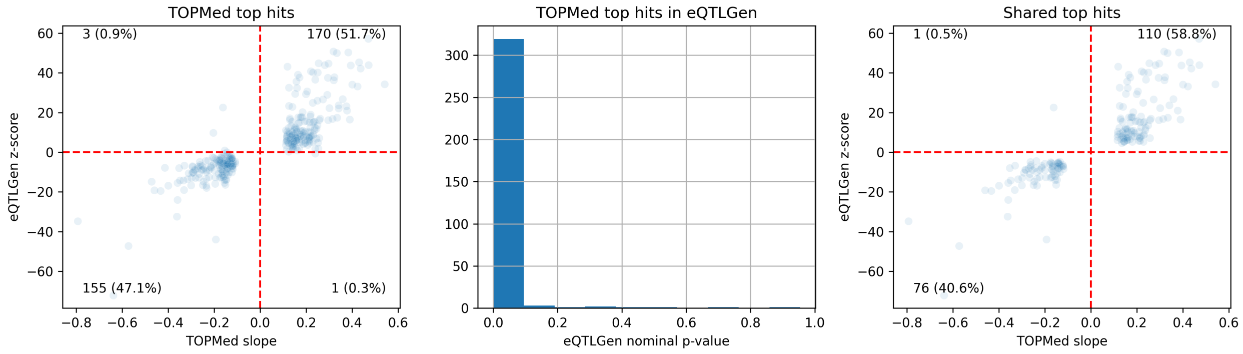


**Figure S68.** TOPMed primary whole blood trans-eQTLs in eQTLGen. Shared top hits are cases where the same variant is the same significant, top association for the same gene in both studies

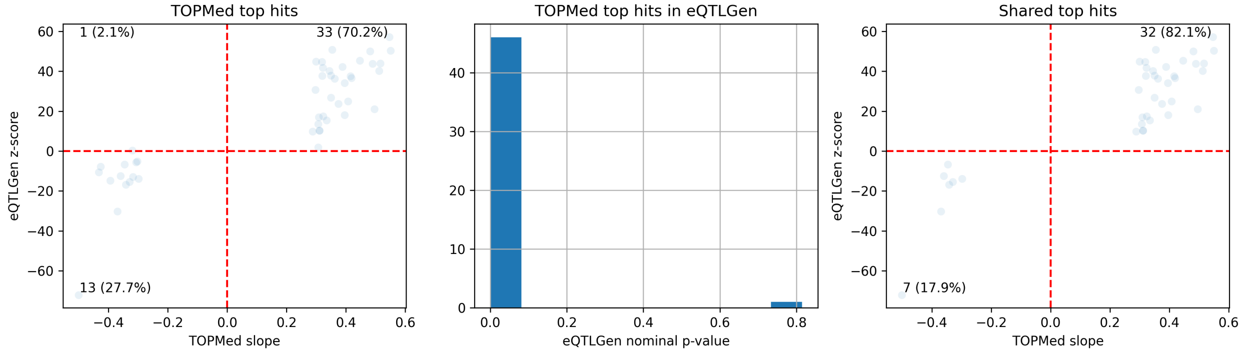


**Figure S69.** TOPMed primary PBMC blood trans-eQTLs in eQTLGen. Shared top hits are cases where the same variant is the same significant, top association for the same gene in both studies
