## Supplementary methods for "Cross-cohort analysis of expression and splicing quantitative trait loci in TOPMed"

### Participating TOPMed cohorts

#### Women's Health Initiative (WHI)

WHI is a long-term, prospective, multi-center cohort study that investigates post-menopausal women's health (The Women's Health Initiative Study Group, 1998). WHI was funded by the National Institutes of Health and the National Heart, Lung, and Blood Institute to study strategies to prevent heart disease, breast cancer, colon cancer, and osteoporotic fractures in women 50-79 years of age. WHI involves 161,808 women recruited between 1993 and 1998 at 40 centers across the US. The study consists of two parts: the WHI Clinical Trial which was a randomized clinical trial of hormone therapy, dietary modification, and calcium/Vitamin D supplementation, and the WHI Observational Study, which focused on many of the inequities in women's health research and provided practical information about the incidence, risk factors, and interventions related to heart disease, cancer, and osteoporotic fractures.

The Institutional Review Board from the Fred Hutchinson Cancer Center approved the studies. All participants provided written informed consent.

#### Subpopulations and Intermediate Outcome Measures In COPD Study (SPIROMICS)

The Subpopulations and Intermediate Outcome Measures in COPD Study (SPIROMICS) is a multi-center prospective cohort study in which extensive phenotype and molecular data has been collected for the primary purposes of identifying COPD subpopulations as well as identifying and validating intermediate outcome measures for use in future clinical trials. Funded by the NHLBI, SPIROMICS has longitudinally collected phenotypic, biomarker, genetic, genomic, and clinical data from approximately 2,981 participants at 12 study centers across the US. These participants are distributed over four enrollment strata: never-smokers, smokers without COPD, mild/moderate COPD, and severe COPD. Eligible participants were between ages 40 and 80 years old; never smoker was defined as less than 1 pack-year smoking history, and current or former smoker were defined as more than 20 pack-years smoking history. Key exclusion criteria included non-COPD obstructive lung disease, BMI > 40 kg/m2 at baseline, and diagnosis of unstable cardiovascular disease. Blood samples sent for RNA sequencing were obtained at clinical visits at baseline and one year later. The study design has been described in detail elsewhere (Couper et al., 2014).

The Institutional Review Boards for the participating sites (University of North Carolina at Chapel Hill, Columbia University, Temple University, Johns Hopkins University, Wake Forest University, University of Michigan, University of Illinois at Chicago, University of Iowa, University of Utah, National Jewish Health, University of California at San Francisco, and University of California at Los Angeles) approved the studies. All participants provided written informed consent.

#### Multi-Ethnic Study of Atherosclerosis (MESA)

The Multi-Ethnic Study of Atherosclerosis (MESA) is a study of the characteristics of subclinical cardiovascular disease and the risk factors that predict progression to clinically overt cardiovascular disease or progression of the subclinical disease (Bild et al., 2002). MESA consisted of a diverse, population-based sample of an initial 6,814 asymptomatic men and women aged 45-84. 38 percent of the recruited participants were white, 28 percent African American, 22 percent Hispanic, and 12 percent Asian, predominantly of Chinese descent. Participants were recruited from six field centers across the United States: Wake Forest University, Columbia University, Johns Hopkins University, University of Minnesota, Northwestern University and University of California - Los Angeles. Participants are being followed for identification and characterization of cardiovascular disease events, including acute myocardial infarction and other forms of coronary heart disease (CHD), stroke, and congestive heart failure; for cardiovascular disease interventions; and for mortality. The first examination took place over two years, from July 2000 - July 2002. It was followed by five examination periods that were 17-20 months in length, including the recently completed Exam 6 (2016-2018). MESA Exam 7 is approved and now being planned. Participants have been contacted every 9 to 12 months throughout the study to assess clinical morbidity and mortality. Informed consent was obtained for extensive data sharing (dbGaP) and genetic/omic studies, including candidate genes (NHLBI CARe), genome-wide scans (NHLBI SHARe), exome sequencing (NHLBI ESP) and, most recently, the NHLBI TOPMed program.

#### Framingham Heart Study (FHS)

Established in 1948, FHS is one of the oldest observational cohort studies for studying cardiovascular diseases and their risk factors. FHS has recruited over 15,000 largely caucasian participants, spanning over three generations. The gene expression component of the study was largely taken from the whole blood of the Offspring cohort (Pubmed 1208363) and the Third Generation Cohort (Pubmed 17372189) participants. All protocols for participant examinations and collection of genetic materials for this study were approved by the Boston Medical Center Institutional Review Board. All FHS participants gave written, informed consent.

#### Lung Tissue Research Consortium (LTRC)

The LTRC is an NHLBI-funded program that collected excess surgical lung tissue and blood samples on participants undergoing clinically indicated thoracic surgery, including lung transplantation, lung volume reduction surgery, nodule resection, and biopsy, enrolled between 2005 and 2019 (reference: NCT02988388 and PMC2862780?). Lung tissue samples were flash frozen and separated from possible cancer regions when present. Phenotypes include lung function, standardized questionnaires, and CT scans. The majority of samples are from three primary groups: COPD; interstitial lung disease, mostly idiopathic pulmonary fibrosis; and control subjects. A smaller number of subjects with other lung diseases, such as sarcoidosis, were also included. All LTRC subjects underwent whole genome sequencing through TOPMed.

#### COPDGene Study (COPDGene)

COPDGene is an observational study of 10,198 self-identified non-Hispanic White and African-American participants who were current and former smokers with and without COPD, as well as 454 additional non-smoking controls. Phenotypes at the baseline study visit (2007-2012) included questionnaires, pre- and post-bronchodilator spirometry, high-resolution chest CT scan (inspiratory and expiratory), and samples for genetic analysis, including whole genome sequencing. At the Phase 2 follow-up visit five years after the initial visit, 6,284 subjects completed similar phenotype assessments and provided blood sample collection for RNA. A subset of these subjects also provided nasal epithelial cells. Phase 3 (ten-year follow-up) visits are currently underway.

#### Gene-Environments and Admixture in Latino Asthmatics (GALA II) and Study of African Americans, Asthma, Genes, & Environments (SAGE)

Gene-Environments and Admixture in Latino Asthmatics (GALA II) study and Study of African Americans, Asthma, Genes, & Environments (SAGE) were parallel, case-control studies of asthma in children, designed with similar study protocols and questionnaires. Details of the study design have been described elsewhere (Borrell et al., 2013; Nishimura et al., 2013; Oh et al., 2012; Thakur et al., 2013). Briefly, SAGE recruited participants through a combination of clinic- and community-based recruitment centers in the San Francisco Bay Area. GALA II recruited participants from 5 urban study centers across the mainland United States (San Francisco Bay Area, New York City, Chicago, Houston) and Puerto Rico. Participants were eligible if they were 8-21 years of age with physician-diagnosed asthma and no history of other lung or chronic illnesses. Self-identification of race/ethnicity was required for enrollment. Parents and grandparents of participants self-identified as Latino (GALA II) or African American (SAGE). Study exclusion criteria included the following: 1) any smoking within one year of the recruitment date; 2) 10 or more pack-years of smoking; 3) pregnancy in the third trimester; 4) history of lung diseases other than asthma (for cases) or chronic illness (for cases and controls).

The local institutional review board from the University of California San Francisco Human Research Protection Program approved the studies (IRB# 10-02877 for SAGE and 10-00889 for GALA II). All subjects and their legal guardians provided written informed consent. Participants 17 or younger also provided age-appropriate assent.

### Replication of GTEx trans-e/sQTLs in TOPMed

Of the three lung trans-eQTLs discovered in GTEx, two were exactly replicated in TOPMed lung (same gene and lead variant with same direction of effect), while one did not replicate (the GTEx trans-eGene was not a trans-eGene in TOPMed lung; the GTEx eVariant did not pass QC in TOPMed genotype freeze 9b) (Table S15). Of thirteen whole blood trans-eQTLs discovered in GTEx, 7 had nominal p-value < 0.05 in TOPMed. Of the thirteen trans-eGenes, 8 trans-eGenes were trans-eGenes in TOPMed; of these, 2 had the same lead variant, 3 had different lead variants but the lead variants were in close proximity (<10kb) or in high LD ($R^2$ >= 0.8), and 3 had lead variants on separate chromosomes.

Two GTEx lung/whole blood trans-sQTL were in high LD ($R^2$ > 0.8) with a TOPMed trans-sQTL from the same tissue; the remaining GTEx whole blood trans-sQTL was not in high LD (and the variant did not pass QC in TOPMed freeze 9b), but it was at the same locus as a TOPMed trans-sQTL (the GTEx and TOPMed trans-sQTL variants were ~1.1kb away) (Table S16)

$R^2$ values in the above-cited supplementary tables were extracted from LDPair (EUR populations). Primary trans-e/sVariants were not clumped prior to these comparisons.

### Replication of eQTLGen and DIRECT trans-eQTLs in TOPMed

$\pi_1$ statistics and concordance in direction-of-effect (see Discussion) were calculated using the genes tested in TOPMed whole blood, after lifting to hg38. Variants not present in TOPMed were excluded.

### TOPMed trans-eQTLs in eQTLGen and DIRECT

eQTLGen performed trans-eQTL scans using 10,317 previously-published trait-associated SNPs. The TOPMed trans-eGene - trans-eVariant pairs for only 329 whole blood trans-eQTL and 47 PBMC trans-eQTL were considered in eQTLGen; for these TOPMed gene - variant pairs, the gene was an eQTLGen trans-eGene and the TOPMed variant matched the top eQTLGen variant for 187 (56.8%) whole blood and 39 (83.0%) PBMC gene - variant pairs (Fig S68-69).

Only 464 TOPMed trans-eGenes were also trans-eGenes in DIRECT. For those 464 trans-eGenes, the TOPMed primary trans-eQTL was in high LD ($R^2$ >= 0.8) with one of the DIRECT trans-eQTLs in 36.6% of cases (170 / 464).

### TOPMed Banner Authorship

Namiko Abe[1], Laura Almasy[2], Seth Ament[3], Pramod Anugu[4], Paul Auer[5,5], Dimitrios Avramopoulos[6], Adithya Balasubramanian[7], R. Graham Barr[8,8], Lucas Barwick[9], Terri Beaty[6,6], Diane Becker[6], Lewis Becker[6], Amber Beitelshees[3], Takis Benos[10], Marcos Bezerra[11], Joshua Bis[12,12], Jennifer Brody[12,12], Ulrich Broeckel[13], Jai Broome[12], Karen Bunting[1], Erin Buth[12], Vincent Carey[14], Cara Carty[15], Richard Casaburi[16], Mark Chaffin[17], Christy Chang[3], Yi-Cheng Chang[18], Sameer Chavan[19], Bo-Juen Chen[1], Wei-Min Chen[20], Seung Hoan Choi[17], Lee-Ming Chuang[18,18], Ren-Hua Chung[21], Matthew Conomos[12], Elaine Cornell[22], Carolyn Crandall[16], James Crapo[23], Jeffrey Curtis[24], Coleen Damcott[3], Sean David[25], Lisa de las Fuentes[26], Paul de Vries[27], Ranjan Deka[28], Dawn DeMeo[14,14,14], Scott Devine[3], Huyen Dinh[7], Harsha Doddapaneni[29], Qing Duan[30,30], Ravi Duggirala[31], Charles Eaton[32], Lynette Ekunwe[4], Adel El Boueiz[33,33], Leslie Emery[12], Charles Farber[20], Jesse Farek[7], Nora Franceschini[30,30], Chris Frazar[12], Mao Fu[3], Stephanie M. Fullerton[12], Lucinda Fulton[34], Shanshan Gao[19], Yan Gao[4], Margery Gass[35], Heather Geiger[36],

Auyon Ghosh[14,14], Chris Gignoux[37], David Glahn[38], Stephanie Gogarten[12], Da-Wei Gong[3], Harald Goring[39], Daniel Grine[19], C. Charles Gu[34], Yue Guan[3], Michael Hall[40], Yi Han[7], Daniel Harris[41], Ben Heavner[12], David Herrington[42], Brian Hobbs[14,14], Elliott Hong[3], Karin Hoth[43], Chao (Agnes) Hsiung[21], Jianhong Hu[7], Yi-Jen Hung[44], Haley Huston[45], Chii Min Hwu[46], Rebecca Jackson[47], Deepti Jain[12], Jill Johnsen[48,48], Rich Johnston[49], Kimberly Jones[6], Michael Kessler[3], Alyna Khan[12], Ziad Khan[7], Wonji Kim[33,33], John Kimoff[50], Greg Kinney[51], Holly Kramer[52], Christoph Lange[53,53], Ethan Lange[19], Cathy Laurie[12,12], Cecelia Laurie[12], Meryl LeBoff[14], Sandra Lee[7], Wen-Jane Lee[46], David Levine[12], Joshua Lewis[3], Yun Li[30,30,30], Xihong Lin[53], Simin Liu[32,32], Yu Liu[37], Barry Make[6], Alisa Manning[54,54], JoAnn Manson[14], Lisa Martin[55], Melissa Marton[36], Susan Mathai[19], Susanne May[12], Patrick McArdle[3], Merry-Lynn McDonald[56], Sean McFarland[33], Daniel McGoldrick[12,12], Caitlin McHugh[12], Hao Mei[4], James Meigs[57], Vipin Menon[7], Nancy Min[4], Matt Moll[14], Zeineen Momin[7], May Montasser[58], Josyf C Mychaleckyj[20], Rakhi Naik[6], Take Naseri[59], Pradeep Natarajan[17], Sarah C. Nelson[12], Bonnie Neltner[19], Caitlin Nessner[7], Osuji Nkechinyere[7], Jeff O'Connell[60], Tim O'Connor[3], Heather Ochs-Balcom[61], Geoffrey Okwuonu[7], James Pankow[62,62], Cora Parker[63], Gina Peloso[64], Juan Manuel Peralta[31], Marco Perez[37], James Perry[3], Ulrike Peters[35], Lawrence S Phillips[49], Toni Pollin[3], Julia Powers Becker[19], Meher Preethi Boorgula[19], Bruce Psaty[12], Dandi Qiao[14,14], Nicholas Rafaels[65], Mahitha Rajendran[7], Laura Rasmussen-Torvik[66], Aakrosh Ratan[20], Robert Reed[3], Elizabeth Regan[23], Muagututi‘a Sefuiva Reupena[67], Rebecca Robillard[68], Carolina Roselli[17], Ingo Ruczinski[6,6], Alexi Runnels[36], Pamela Russell[19], Kathleen Ryan[3], Ester Cerdeira Sabino[69], Shabnam Salimi[70], Sejal Salvi[7], Steven Salzberg[6], Kevin Sandow[71], Jireh Santibanez[7], Karen Schwander[34], Frank Sciurba[10,10], Frédéric Sériès[72], Amol Shetty[3], Aniket Shetty[19], Brian Silver[73], Robert Skomro[74], Tanja Smith[1], Sylvia Smoller[75], Beverly Snively[42], Adrienne M. Stilp[12], Garrett Storm[51], Elizabeth Streeten[3], Jessica Lasky Su[14], Yun Ju Sung[34], Jody Sylvia[14], Adam Szpiro[12], Margaret Taub[6,6], Simeon Taylor[3], Timothy A. Thornton[12], Machiko Threlkeld[12], Lesley Tinker[35], David Tirschwell[12], Hemant Tiwari[56,56], Catherine Tong[12], Michael Tsai[62], Dhananjay Vaidya[6], Tarik Walker[19], Robert Wallace[43], Avram Walts[19], Fei Fei Wang[12], Heming Wang[76], Karol Watson[16], Jennifer Watt[7], Lu-Chen Weng[57], Jennifer Wessel[77,77], Kayleen Williams[12], Carla Wilson[14], James Wilson[78,78], Lara Winterkorn[36], Quenna Wong[12], Baojun Wu[79], Huichun Xu[3], Lisa Yanek[6], Ivana Yang[19], Seyedeh Maryam Zekavat[17], Snow Xueyan Zhao[23], Wei Zhao[24], Xiaofeng Zhu[80]

1 - New York Genome Center, New York, New York, 10013, United States of America; 2 - Children's Hospital of Philadelphia, University of Pennsylvania, Philadelphia, Pennsylvania, 19104, United States of America; 3 - University of Maryland, Baltimore, Maryland, 21201, United States of America; 4 - University of Mississippi, Jackson, Mississippi, 38677, United States of America; 5 - Medical College of Wisconsin, Milwaukee, Wisconsin, 53211, United States of America; 6 - Johns Hopkins University, Baltimore, Maryland, 21218, United States of America; 7 - Baylor College of Medicine Human Genome Sequencing Center, Houston, Texas, 77030, United States of America; 8 - Columbia University, New York, New York, 10032, United States of America; 9 - The Emmes Corporation, Rockville, Maryland, 20850, United States of America; 10 - University of Pittsburgh, Pittsburgh, Pennsylvania, 15260, United States of America; 11 - Fundação de Hematologia e Hemoterapia de Pernambuco - Hemope, Recife, 52011-000, Brazil; 12 - University of Washington, Seattle, Washington, 98195, United States of America; 13 - Medical College of Wisconsin, Milwaukee, Wisconsin, 53226, United States of America; 14 -

Brigham & Women's Hospital, Boston, Massachusetts, 02115, United States of America; 15 - Washington State University, Pullman, Washington, 99164, United States of America; 16 - University of California, Los Angeles, Los Angeles, California, 90095, United States of America; 17 - Broad Institute, Cambridge, Massachusetts, 02142, United States of America; 18 - National Taiwan University, Taipei, 10617, Taiwan (Province of China); 19 - University of Colorado at Denver, Denver, Colorado, 80204, United States of America; 20 - University of Virginia, Charlottesville, Virginia, 22903, United States of America; 21 - National Health Research Institute Taiwan, Miaoli County, 350, Taiwan (Province of China); 22 - University of Vermont, Burlington, Vermont, 05405, United States of America; 23 - National Jewish Health, Denver, Colorado, 80206, United States of America; 24 - University of Michigan, Ann Arbor, Michigan, 48109, United States of America; 25 - University of Chicago, Chicago, Illinois, 60637, United States of America; 26 - Washington University in St Louis, St. Louis, Missouri, 63110, United States of America; 27 - University of Texas Health at Houston, Houston, Texas, 77030, United States of America; 28 - University of Cincinnati, Cincinnati, Ohio, 45220, United States of America; 29 - Baylor College of Medicine Human Genome Sequencing Center, Houston, Texas, 77030; 30 - University of North Carolina, Chapel Hill, North Carolina, 27599, United States of America; 31 - University of Texas Rio Grande Valley School of Medicine, Edinburg, Texas, 78539, United States of America; 32 - Brown University, Providence, Rhode Island, 02912, United States of America; 33 - Harvard University, Cambridge, Massachusetts, 02138, United States of America; 34 - Washington University in St Louis, St Louis, Missouri, 63130, United States of America; 35 - Fred Hutchinson Cancer Research Center, Seattle, Washington, 98109, United States of America; 36 - New York Genome Center, New York City, New York, 10013, United States of America; 37 - Stanford University, Stanford, California, 94305, United States of America; 38 - Boston Children's Hospital, Harvard Medical School, Boston, Massachusetts, 02115, United States of America; 39 - University of Texas Rio Grande Valley School of Medicine, San Antonio, Texas, 78229, United States of America; 40 - University of Mississippi, Jackson, Mississippi, 39216, United States of America; 41 - University of Maryland, Philadelphia, Pennsylvania, 19104, United States of America; 42 - Wake Forest Baptist Health, Winston-Salem, North Carolina, 27157, United States of America; 43 - University of Iowa, Iowa City, Iowa, 52242, United States of America; 44 - Tri-Service General Hospital National Defense Medical Center, Taiwan (Province of China); 45 - Blood Works Northwest, Seattle, Washington, 98104, United States of America; 46 - Taichung Veterans General Hospital Taiwan, Taichung City, 407, Taiwan (Province of China); 47 - Oklahoma State University Medical Center, Columbus, Ohio, 43210, United States of America; 48 - University of Washington, Seattle, Washington, 98109, United States of America; 49 - Emory University, Atlanta, Georgia, 30322, United States of America; 50 - McGill University, Montréal, QC H3A 0G4, Canada; 51 - University of Colorado at Denver, Aurora, Colorado, 80045, United States of America; 52 - Loyola University, Maywood, Illinois, 60153, United States of America; 53 - Harvard School of Public Health, Boston, Massachusetts, 02115, United States of America; 54 - Broad Institute, Harvard University, Massachusetts General Hospital; 55 - George Washington University, Washington, District of Columbia, 20037, United States of America; 56 - University of Alabama, Birmingham, Alabama, 35487, United States of America; 57 - Massachusetts General Hospital, Boston, Massachusetts, 02114, United States of America; 58 - National Heart, Lung, and Blood Institute, Bethesda, Maryland, 20817, United States of America; 59 - Ministry of Health,

Government of Samoa, Apia, Samoa; 60 - University of Maryland, Balitmore, Maryland, 21201, United States of America; 61 - University at Buffalo, Buffalo, New York, 14260, United States of America; 62 - University of Minnesota, Minneapolis, Minnesota, 55455, United States of America; 63 - RTI International, Research Triangle Park, North Carolina, 27709-2194, United States of America; 64 - Boston University, Boston, Massachusetts, 02118, United States of America; 65 - University of Colorado at Denver, Denver, Colorado, 80045, United States of America; 66 - Northwestern University, Chicago, Illinois, 60208, United States of America; 67 - Lutia I Puava Ae Mapu I Fagalele, Apia, Samoa; 68 - University of Ottawa, Ottawa, ON K1Z 7K4, Canada; 69 - Universidade de Sao Paulo, Sao Paulo, 01310000, Brazil; 70 - University of Maryland, Seattle, Washington, 98195, United States of America; 71 - Lundquist Institute, Torrance, California, 90502, United States of America; 72 - Université Laval, Quebec City, G1V 0A6, Canada; 73 - UMass Memorial Medical Center, Worcester, Massachusetts, 01655, United States of America; 74 - University of Saskatchewan, Saskatoon, SK S7N 5C9, Canada; 75 - Albert Einstein College of Medicine, New York, New York, 10461, United States of America; 76 - Brigham & Women's Hospital, Mass General Brigham, Boston, Massachusetts, 02115, United States of America; 77 - Indiana University, Indianapolis, Indiana, 46202, United States of America; 78 - Beth Israel Deaconess Medical Center, Cambridge, Massachusetts, 02139, United States of America; 79 - Henry Ford Health System, Detroit, Michigan, 48202, United States of America; 80 - Case Western Reserve University, Cleveland, Ohio, 44106, United States of America

https://doi.org/10.1016/s0197-2456(97)00078-0
